# Association between COVID-19 Outcomes and Mask Mandates, Adherence, and Attitudes

**DOI:** 10.1101/2021.01.19.21250132

**Authors:** Dhaval Adjodah, Karthik Dinakar, Matteo Chinazzi, Samuel P. Fraiberger, Alex Pentland, Samantha Bates, Kyle Staller, Alessandro Vespignani, Deepak L. Bhatt

## Abstract

We extend previous studies on the impact of masks on COVID-19 outcomes by investigating an unprecedented breadth and depth of health outcomes, geographical resolutions, types of mask mandates, early versus later waves and controlling for other government interventions, mobility testing rate and weather. We show that mask mandates are associated with a statistically significant decrease in new cases (-3.55 per 100K), deaths (-0.13 per 100K), and the proportion of hospital admissions (-2.38 percentage points) up to 40 days after the introduction of mask mandates both at the state and county level. These effects are large, corresponding to 14% of the highest recorded number of cases, 13% of deaths, and 7% of admission proportion. We also find that mask mandates are linked to a 23.4 percentage point increase in mask adherence in four diverse states. Lastly, using a large novel survey dataset of almost half a million people in 68 countries, we introduce the novel results that community mask adherence and community attitudes towards masks are associated with a reduction in COVID-19 cases and deaths. Our results have policy implications for reinforcing the need to maintain and encourage mask-wearing by the public, especially in light of some states starting to remove their mask mandates.

## 1 Introduction

As of December 2020, SARS-CoV-2, the virus responsible for COVID-19 has infected at least 66 million people worldwide and caused more than 1.5 million deaths [1]. Numerous studies have analyzed the role played by masks during the COVID-19 pandemic [2–6]: masks have been associated with a reduction in the infection rate among health care workers in a large hospital network [7], mask mandates have helped reduce the number of cases in the United States and in Germany [8–10], and simulations have shown that wearing a mask can protect against droplet infection by preventing the spread of viral particles [11–16]. Despite this evidence, there has been strong resistance against mask-wearing [17–20], and the states of Texas and Mississippi recently lifted their mask mandates even though they have some of the lowest vaccination rates in the country.

To strengthen the case that masks have a positive effect on the pandemic, we extend previous studies by investigating an unprecedented breadth and depth of outcomes, geographical resolutions, types of mask mandates, control variables, early versus later waves and controlling for other government interventions (such as stay-at-home orders). For example, it has been shown that state-level mask mandates have a positive effect on cases [8] using the same methodology as ours, the event study framework. Building on this work, we investigate the effect of mandates not only on cases, but also on hospitalizations and deaths, adding new controls for individual mobility, temperature and precipitation, COVID-19 test rates, and also investigating the effect of county-level mask mandates. Similarly, mask mandates have been found to be beneficial in a study of cases in Germany [10], but without looking at hospitalization and deaths, and not controlling for critically important temporally evolving variables such as mobility and testing rate (additionally, we expand our investigation to 68 countries). The work closest to ours investigates causal impact of mask policies [21] but does not look at hospitalizations, county-level mandates and outcomes, different types of mandates, earlier vs. later affected states, etc. However, we base our approach of how to control for voluntary individual behavioral responses to information on their causal graphical model.

Overall, our work makes the following novel contributions relative to recent studies: we look at a much larger number of counties (covering 77% of the US population) and all 50 states and the District of Columbia controlling for a number of temporally evolving and geography-specific variables compared to [9]; we investigate longer time horizon treatment effects and expand beyond hospital populations compared to [7]; we include states beyond New York and Washington compared to [11]; we include outcomes beyond hospitalization, look at all states and investigate county-level effects compared to [22]; we investigate the temporal evolution of mask mandates before and after their introduction compared to [6]; and we analyze much larger populations and include the temporal evolution and geographical heterogeneity of treatment effects compared to [23].

In addition to strengthening evidence of the effect of mask mandates on COVID-19 outcomes, this work introduces two novel contributions to the research of masks: we investigate the effect of mask mandates on mask adherence (the percentage of people who wear masks) and the effect of mask adherence on COVID-19 cases and deaths. Secondly, using a novel dataset of survey responses in 68 countries around the world across months-long waves of surveys on 479 thousand people in 51 languages, we investigate the effect of community mask adherence (how many people in a community wear masks) and community mask attitudes (how many people in a community believe that masks are an important for COVID-19) on country-level cases and deaths. This is especially important as similar community and adherence drivers behind mask avoidance [24] might impact the on-going deployment of vaccines.

This paper is structured as follows: in the next section, we describe our results: the main result is that mask mandates are associated with a significant improvement in COVID-19 outcomes (corresponding to 14% of the highest recorded number of daily new cases, 13% of deaths, and 7% of hospital admissions). As described in the Materials and Methods section, our analysis covers all 50 states (including the District of Columbia) and 857 counties (representing 77% of the US population) from Feb 2, 2020 to September 27, 2020, and we include a large number of robustness tests.

In support of our main result, we also observe that mask mandates are associated with a 23.4 percentage point increase in mask adherence in four geographically, culturally and politically diverse states – Hawaii, Iowa, North Dakota and New Hampshire – and that mask adherence is itself associated with a significant decrease in COVID-19 cases and deaths in all 50 states and D.C. Finally, using a large novel dataset [25], we observe that community mask adherence and attitudes towards masks are associated with fewer cases and deaths in 68 countries, controlling for population density, human development and mobility. We end by discussing the limitations and implications of this work.

Taken together, these results strongly suggest the positive effect of mask mandates, mask adherence and mask attitudes on COVID-19 cases, hospitalizations and deaths.

## 2 Results

### 2.1 Mask Mandates and COVID-19 Outcomes

As show in Fig 1, mask mandates were introduced at day zero, shown by the red vertical line. Because we are using an event study model, the y-axis represents the treatment effect associated with mask mandates on the COVID-19 outcome for each day before or after the introduction of a mask mandate relative to the day before the introduction of a mask mandate.

**Figure 1:**
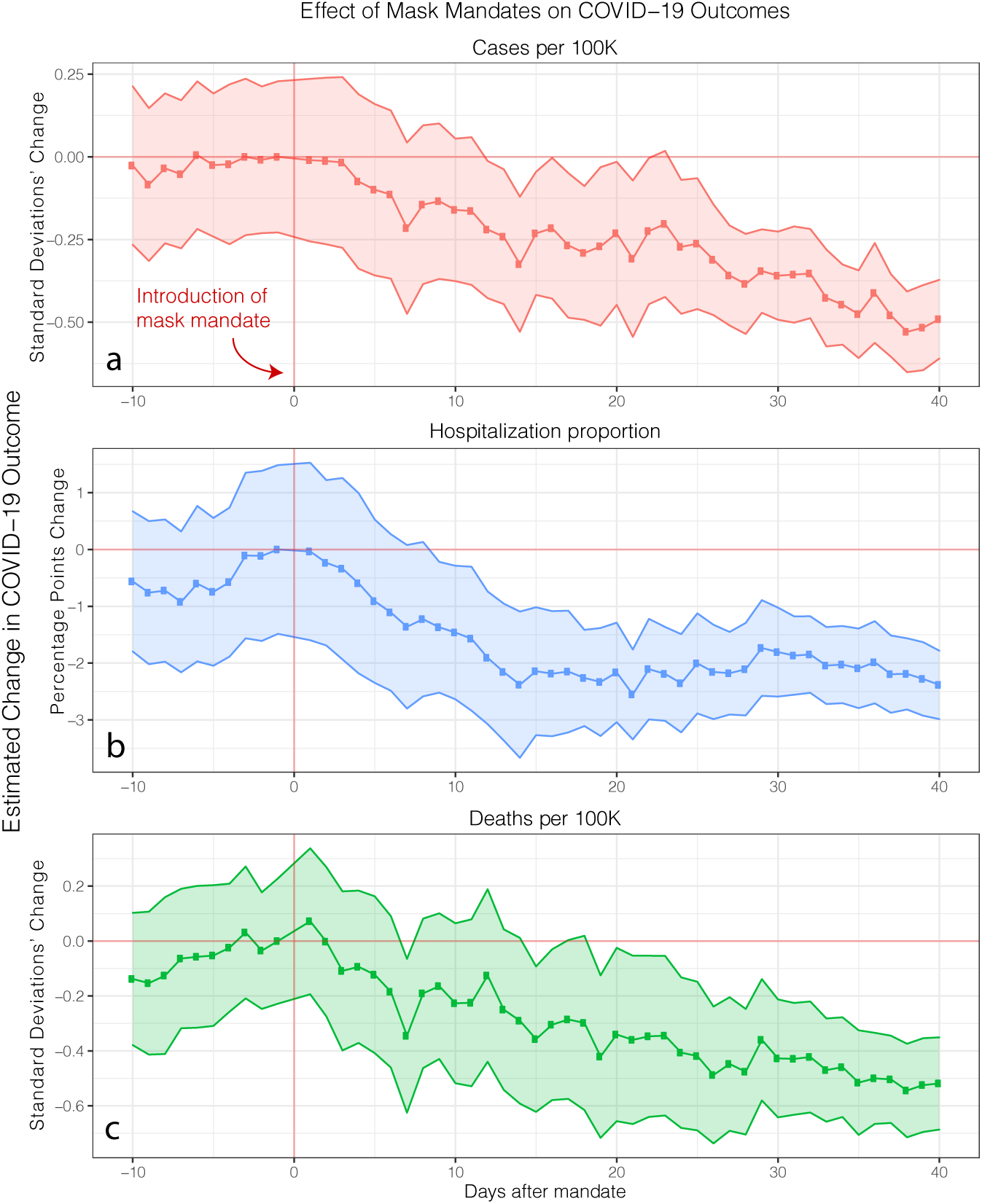
Effect of state-level mask mandates on COVID-19 outcomes. Using an event study design, we estimate the treatment effect (y-axis) of mask mandates on Z-scored population-normalized COVID-19 daily new confirmed cases, daily new hospitalization admissions proportion, and deaths across all 50 states and D.C. over the time period between February 1 and September 27, 2020. Shaded area represents 95% confidence interval, with standard errors clustered at the state level. Red vertical line represents the start of the mask mandate, and the red horizontal line represents the line of zero treatment effect. We observe the treatment effect to start near zero when mask mandates are introduced (red vertical line) and to gradually increase (i.e. the treatment effect becomes more negative) reaching, 40 days after the introduction of mask mandates, -0.49 standard deviations (95% confidence interval is [-0.61,-0.37], adjusted *R*^2^ is 0.632 and *p <* 0.001) for new cases **(a)**, -2.38 percentage points ([-2.99,-1.78], adjusted *R*^2^ is 0.698 and *p <* 0.001) for the proportion of daily hospitalization admissions **(b)** due to COVID-19, and 0.52 standard deviations (95% confidence interval is [-0.69,-0.35], adjusted *R*^2^ is 0.537 and *p <* 0.001) for deaths **(c)**. These effects are large, corresponding to 14% of the highest recorded number of cases, 13% of deaths, and 7% of admission proportion.

For all three outcomes, we observe the treatment effect (y-axis) to be negative which means that mask mandates are associated with a decrease in cases, hospitalizations and deaths. Specifically, we observe this treatment effect to start near zero when mask mandates are introduced and to gradually increase (i.e. the treatment effect becomes more negative) reaching, 40 days after the introduction of mask mandates, -0.49 standard deviations (95% confidence interval is [-0.61,-0.37], adjusted *R*^2^ is 0.632 and *p <* 0.001) for new cases, -2.38 percentage points ([-2.99,-1.78], adjusted *R*^2^ is 0.698 and *p <* 0.001) for the proportion of daily hospitalization admissions due to COVID-19, and 0.52 standard deviations (95% confidence interval is [-0.69,-0.35], adjusted *R*^2^ is 0.537 and *p <* 0.001) for deaths.

In real terms, these treatment effects are large, corresponding to an associated decrease of 3.55 cases per 100,000 people (or 14% of the highest recorded number of new cases per 100,000 people during our observation period), 0.13 deaths per 100,000 people (or 13% of the highest recorded number of new deaths), and a 7% decrease compared to the highest recorded proportion (34%) of COVID-19 related hospitalization admissions during our observation period. As detailed in the specification of the model in the Methods section 4.2.2, we control for many confounders such as other NPIs (stay-at-home and non-essential business closures), mobility, weather, new test rates and various aspects of behavior change.

#### 2.1.1 Mask mandate scope: business employees vs. the public

We conduct a number of checks to test the robustness of our results. First, we investigate changing how mask mandates are defined. As noted earlier, so far, we have defined the start of a mask mandate as the earlier date between mandates for all employees or mandates for all members of the public.

Here, we consider two alternative specifications in which we only consider mask mandates requiring everyone to wear a mask outside, and another specification where we only consider mandates requiring only business employees to wear masks. The former specification is more stringent than our original specification (which used the earlier of the two types of mandates) as these public mandates generally came later in the year than those focusing on business employees only, and more states still do not have the public mandates. As shown in Fig 2, the trajectory and magnitude of the treatment effect associated with mask mandates under either types of mandates is consistent with our earlier estimates (shown in Fig 1) which suggests that our treatment effects are not sensitive to our definition of mask mandates.

**Figure 2:**
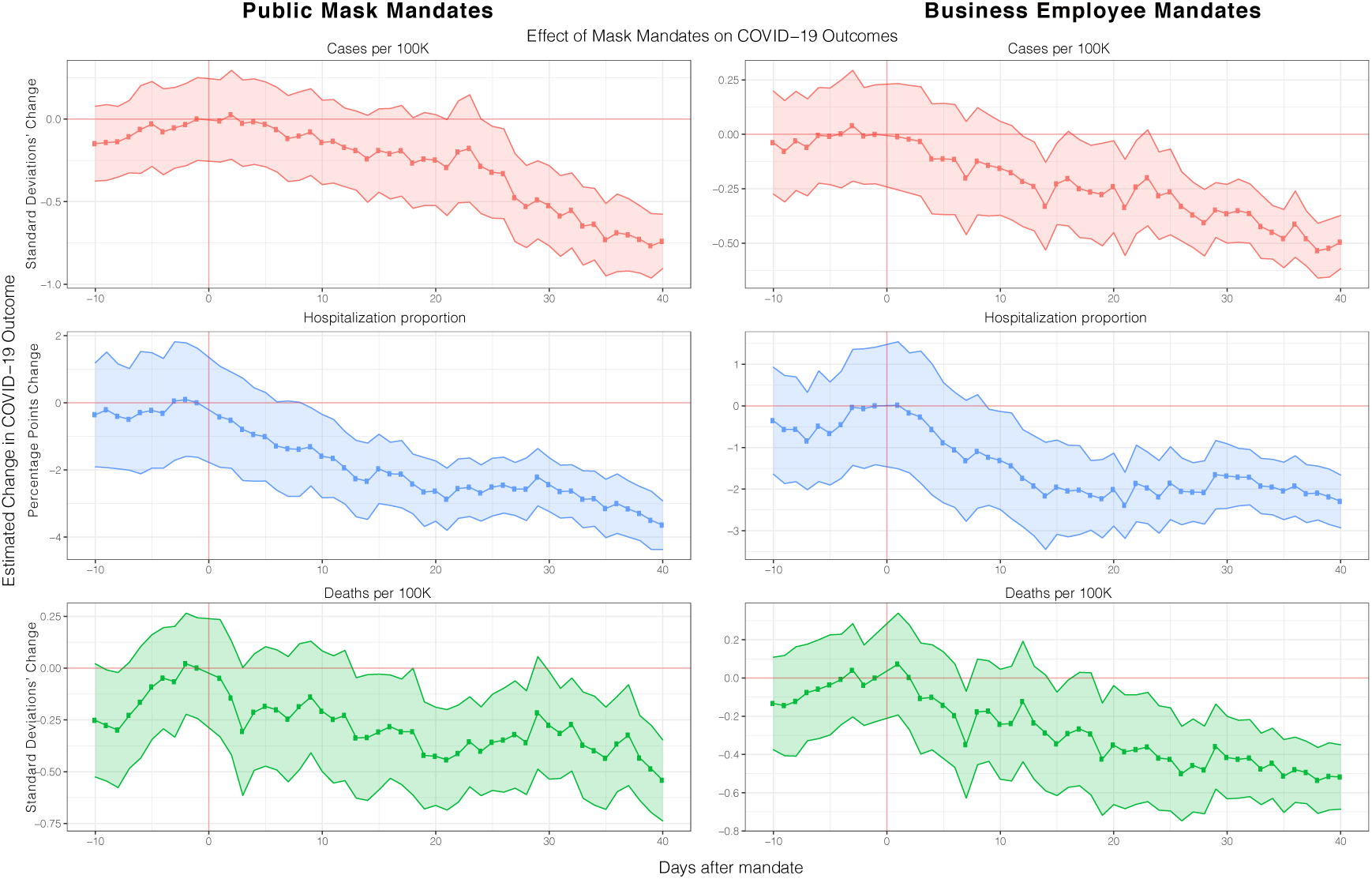
Effect of different types of mask mandates. Robustness check of main event study estimation where, here, we only consider mask mandates that require business employees (right) vs. the public (left) to wear masks. The trajectory and magnitude of the treatment effect is consistent with our previous main result, and do not differ significantly between each other, which demonstrates that our main result is not sensitive to our definition of mask mandate start dates. Full regression results are shown in tables S5 and S6.

#### 2.1.2 Early wave vs. later wave states

So far, we have always looked at all 50 states and the District of Columbia all together. Here we investigate the heterogeneity of treatment effect between earlier and later-wave states as it is possible that the effect of mask mandates on states that were part of the COVID-19 pandemic’s early wave might be different from states that were affected later. For example, mask shortages were present during the early stages of the pandemic [26–28] which might affect the proportion of people who wear masks (mask adherence) following a mask mandate for early states compared to later states.

Therefore, we look separately at the first 15 states to have the highest number of cases per 100K during the month of April (namely, NY, NJ, MA, CT, MI, DC, RI, IL, WA, PA, GA, VT, MD, FL, LA) compared to the rest of the states (i.e. later wave states). As shown in Fig 3, the effect of masks mandates was more gradual and noisier in earlier states whereas they had a faster effect on later states. This is consistent with the literature of behavior change whereby asking people to change their habits (here, mask-wearing) is much harder when the behavior is still very novel [29, 30]. Because the trajectory and magnitude of the treatment effect overall is consistent with our previous main result, this suggests that our specification is not sensitive to subsets of earlier wave, later wave or all states.

**Figure 3:**
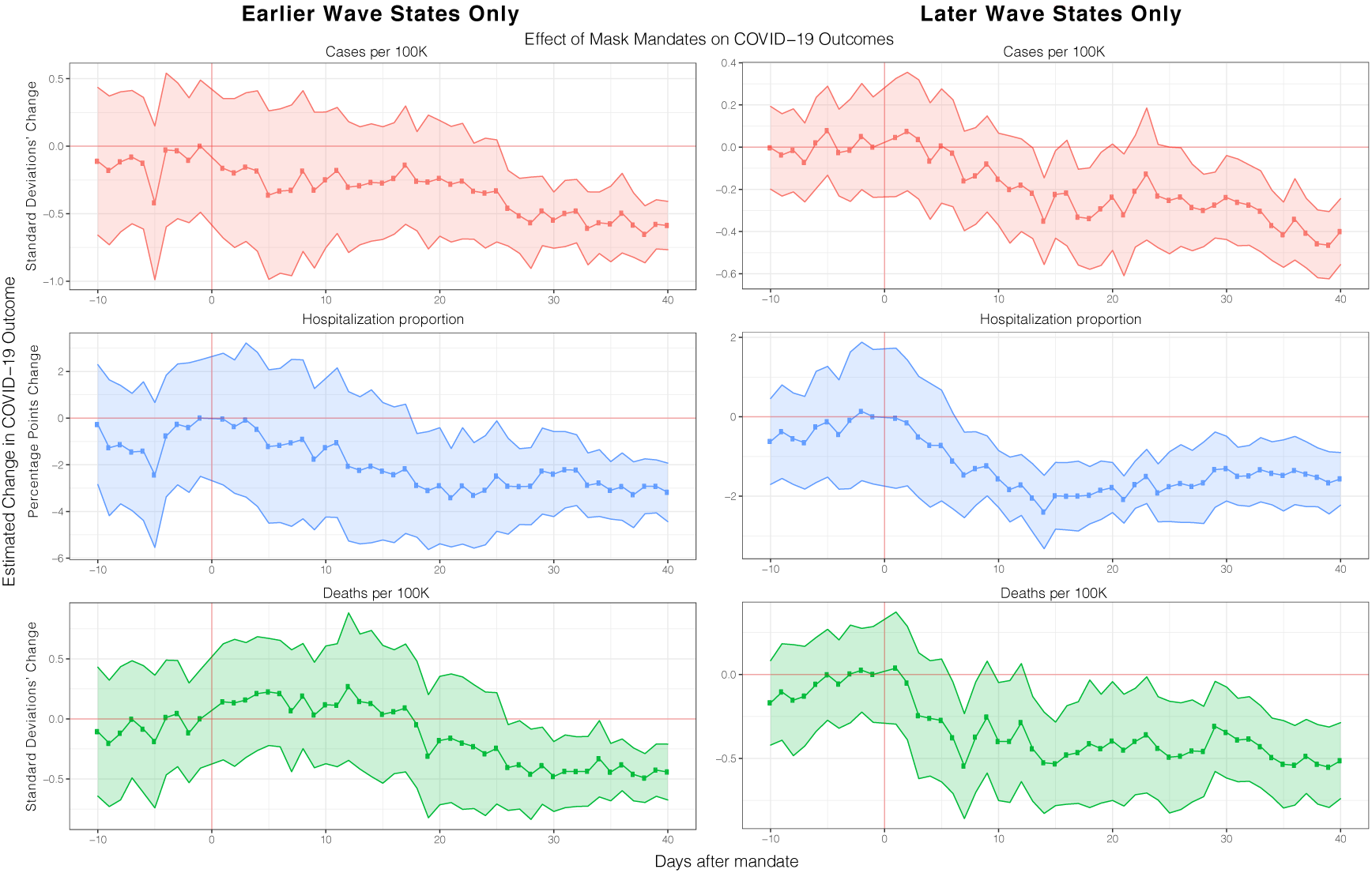
Effect of mask mandates on earlier-hit or later-hit states. Robustness check of main event study results (section 2.1 Fig 1) where we compare the effect of mask mandates on cases, deaths and hospitalization for earlier wave states only (i.e. states who had early spikes of COVID-19 peaks in the pandemic, namely, NY, NJ, MA, CT, MI, DC, RI, IL, WA, PA, GA, VT, MD, FL and LA) vs later wave states. Masks mandates took more time to have an effect on earlier states whereas they had a faster effect on later states. Full regression results are shown in tables S9.

#### 2.1.3 Testing rate exogeneity

We also investigate if the decreases in COVID-19 outcomes we observe might be because the number of COVID-19 tests each day done might be decreasing following the introduction of mask mandates leading us to observe, erroneously, a lower rate of COVID-19 outcomes. Although we already control for the testing rate in our event study specification, here, we also investigate if testing rates were themselves exogenously decreasing during the period of observation. We do so in two ways: first, we look at the per-state and mean (over all states) testing rate (daily new tests per 100K) every day. As shown in supplementary Fig S2 (left), it does not decrease but instead increases over our period of investigation. As a further test, we conduct an event study estimation to investigate a confounding treatment effect associated with mask mandates on testing rate. As shown in Fig S2 (right), there is no associated treatment effect of mask mandates on testing rate. Therefore, it is unlikely that our estimates of decreasing COVID-19 outcomes after the introduction of a mask mandate are due to decreased detection of COVID-19 outcomes.

#### 2.1.4 Controlling for confirmed cases

We can estimate our treatment effect of mandates on the number of deaths and the proportion of hospitalizations when controlling for the number of cases. This specification allows us to investigate whether when we keep the number of cases constant (by controlling for cases), we observe that hospitalizations and deaths still decrease following the introduction of mandates. As shown in supplementary Fig S6, controlling for the number of cases has little effect on the trajectory of the effect of mask mandates on deaths and hospitalization which suggest that the treatment effect of mask mandates is strong even when controlling for cases.

#### 2.1.5 Political affiliation, demographics and trends through time and unit fixed effects

Because we control for time and unit (state or county) fixed effects, we are implicitly controlling for a range of factors that did not change geographically and temporally. For example, including unit fixed effects allows us to control for political affiliation and including the 2018 election vote proportion by political party for each state (or county) correctly did not alter our estimation. The same is true for demographic factors such as age distributions and economic inequality. Similarly, including time fixed effects allows the model to disentangle the particular trajectory in outcome numbers that each state (or county) had over time.

#### 2.1.6 Behavioral change endogeneity and pre-trends

A potential issue with the event study framework is that policy adoption might be endogenous which here might mean that people might be changing their behavior before the policy was introduced, or that they changed their exposure to the virus for other endogenous reasons unrelated to but coinciding with the timing of the introduction of mask mandates. For example, people might be going out less around the time that the mandate was introduced due to fear of infection, had started wearing masks before the mandate was introduced, or that the mandate did not increase mask adherence (how much people wear masks).

We first focus on the issue of pre-trends, which is the fact that people might have anticipatorily started wearing masks even before the introduction of mask mandates. For all three outcomes, the pre-treatment effect is statistically indistinguishable from zero (because 95% confidence interval still contains the zero-effect horizontal zerp line). We do observe a slight upward trend but further investigation using the standard methods for pre-trend robustness checks [31] shows that this pre-trend is still minimal: as show in supplementary Fig S5, omitting time and unit fixed effects and using a longer pre-treatment window of 20 days (we cannot use values earlier than -20 as it leads to some state no longer having enough data as many states implemented mandates very soon after COVID-19 outcome data was available) shows that the anticipatory behavior is indistinguishable from zero for cases and hospitalization, and small for deaths.

Secondly, because we control for six types of mobility (i.e. mobility at the residential, workplace, transit, parks, grocery and pharmacy and retail and recreation) and include the squared values of each mobility indicator as a proxy for social contacts [32, 33], our estimate of the effect of mask mandates has precluded (theoretically, they have been expected out) the effect of people’s endogenous mobility and contact behavior. Similarly, we include controls for the two broadest and far-reaching NPIs, stay-at-home and non-essential business closures, which again precludes the potential confounding effect of other NPIs. Another potential source of endogeneity is the fact that people might be wearing masks (or not wearing them) irrespective of the mask mandates, i.e. the mask mandates themselves did not cause behavior changes. We investigate this in the next section where we show that mask mandates lead to a 23.4 percentage point increase in mask adherence (how many people wear masks) following a mask mandate, and that mask adherence itself are directly associated with improved case and death outcomes in the US, and in 67 other countries.

As another control for behavioral changes, we leverage the causal graphical model of [21] which shows that declines COVID-19 outcomes are not only due to the introduction of policies but are also a result of private behavioral responses to observations of increased outcome numbers. A consequence of their very general causal model is that conditioning on the delayed outcome and the growth rate of the outcome with an approximately 2 week lag enables the confounding path due to behavior change on the outcome to be blocked (see [34] for a review of causal D-separation). As seen in supplementary Fig S1, not controlling for the delayed outcome and growth rate has little effect on the overall trajectory and magnitude of the treatment effect, suggesting that our main model estimation is robust to behavioral endogeneity.

#### 2.1.7 County-level heterogeneity

So far, our analysis has been at the state level. There is significant heterogeneity at the county and city level as to when these smaller units introduced mask mandates [9]. We therefore use a dataset of county-level mask mandates [35] on county-level outcomes and controls to investigate the effect of mask mandates at the county-level. We make sure to only use county-level mask mandates (as opposed to county and state-level mandates) for location which had both, and we do not include data that happened after a state-mandate was also introduced. As shown in supplementary Fig S4, we again observe that mask mandates are associated with a helpful effect on cases, hospitalizations and deaths. However, the treatment effect is noisier as is expected due to a number of noise signals when using data at the county level such as peer-effects of neighboring counties’ mandates due to the underlying interdependence between county mobility patterns [36], as well as the fact that people living in one county often have to travel to a different county to get medical care, resulting in inconsistent accounting of COVID-19 health outcomes at the county level [37].

Overall, our analysis shows that the introduction of mask mandates at the state and county level is associated with a statistically significant and large decrease of cases, hospitalizations, and deaths.

### 2.2 Mask Mandate and Mask Adherence

Here, as a supporting result, we investigate whether the introduction of mask mandates is associated with a significant increase in mask adherence i.e., do mask mandates lead to more people wearing masks? Although there are significant on-going efforts to estimate mask adherence at a population level such as by leveraging computer vision on social media data [38–41], data that tracks mask usage at a county or state level over time has is only now starting to be available: as of September 8, 2020, the CMU Delphi project [42] has started publicly releasing mask adherence data that measures the estimated percentage of people who wore a mask most or all of the time while in public in the past 5 days by asking the question “In the past 5 days, how often did you wear a mask when in public?” in an on-going Facebook online survey ran on millions of people in the US. We therefore use their weighted state-level estimates of mask adherence as our dependent variable. Their weighting strategy to obtain representative samples is described briefly in the Data section, and comprehensively described in their technical report [43].

During the period starting September 8 (when adherence data started being available), only 4 states have enacted new state level requirements: Hawaii and Iowa on November 16, North Dakota on November 14 and New Hampshire on November 20. Although some of these states had county-level mask mandates, had memos encouraging mandates at county or state-level, or had mask requirements in certain businesses, they did not have a state-wide requirement with implementation until the above dates. We run a similar event study estimation as earlier but this time using mask adherence as the outcome variable and for 13 days after mandate introduction (because this is the longest time horizon we have simultaneously for all four states).

As shown in Fig 4, there is a flat pre-treatment trend up to 8 days before the introduction of a mask mandate followed by an uptick in mask adherence after the introduction of mask mandates with a maximum increase in adherence of 23.4 [12.0,34.8] percentage points (adjusted *R*^2^ is 0.925 and *p <* 0.001) 13 days after mask mandate introduction. We observe a delay of about 4 days before we observe an upward trend in mask adherence which is consistent with behavior change literature [29, 30]. As a robustness test, we investigate if our estimation changes significantly if we remove testing rate, and cases and deaths as controls which we included as a confounders because, perhaps, the more case and deaths people see, the more they might be likely to wear masks. As seen in supplementary Fig S7, our estimation is robust to these controls. Unfortunately, we cannot replicate this result at the county level because no county mask mandate datasets currently exists with county-level mask mandates starting during the period that mask adherence data is available. Our results are consistent with previous work [44] although they do not control for time and unit fixed effects which prevents the disentanglement of people already increasing their mask wearing irrespective of the mask mandate.

**Figure 4:**
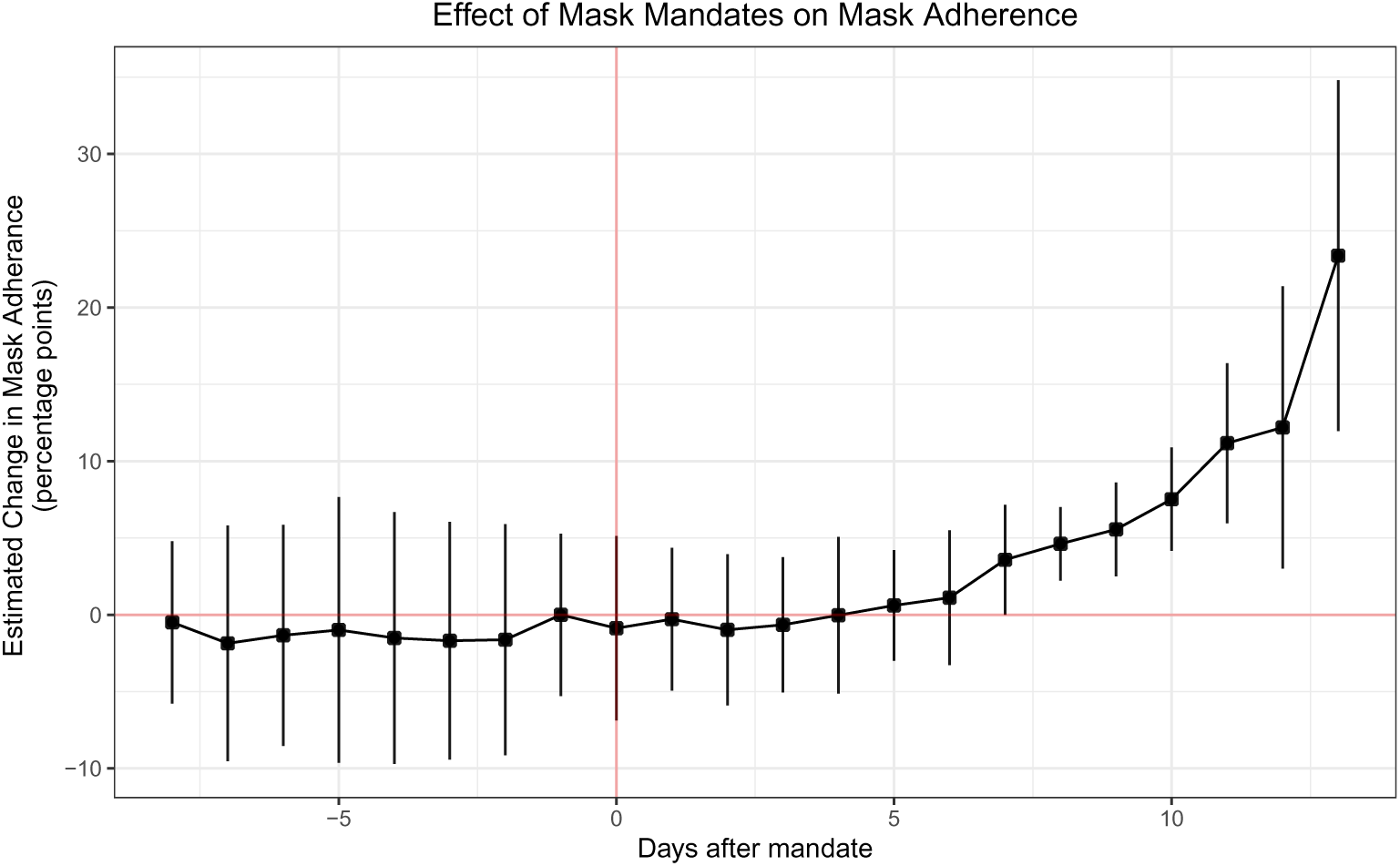
Effect of mask mandates on mask-adherence. Treatment effect of mask mandates on mask adherence in the only four states (Hawaii, Iowa, North Dakota and New Hampshire) which had late mask mandates (Nov 14, 16, and 20) during the period we have daily state-level mask adherence data. We find that there is a flat pre-treatment trend up to 13 days before the introduction of a mask mandate followed by a 23.4 [12.0,34.8] percentage points increase in mask adherence following introduction (adjusted *R*^2^ is 0.925 and *p <* 0.001). This result suggests the strong effect of mask mandates on mask adherence. We expect our results to be a fair estimate of treatment effect in the US due to the fact that these four states are very geographically, culturally and politically diverse.

Although we only estimate the effect of mandates on adherence in these 4 states due to the fact that they are the only ones that implemented mask mandates late enough that we have adherence data for them, we expect this result to support our main result that mask mandates are associated with a significant decrease in COVID-19 outcomes, especially due to the fact that these four states (Hawaii, Iowa, North Dakota and New Hampshire) are very geographically, culturally and politically diverse.

As an additional result, because we have a limited window of time where we have both COVID-19 outcomes (only cases and deaths, no hospitalization data as access ended in September) and mask adherence data, we can use it to investigate if mask adherence is more directly associated with a positive effect on COVID-19 outcomes. To do so, we implement a multi-linear regression of COVID-19 outcomes on compliance, controlled, as before, by a number of factors as specified in equation 4.2.3. Note that we use *all* states (including D.C.) here, not just the four states that had mandates after Sep. 8th. As shown in supplementary table S14, an increase of 1% in mask adherence leads to a decrease of -1.63 [-1.76, -1.50] new confirmed cases per 100K (adjusted *R*^2^ is 0.502 and *p <* 0.001), and -0.016 [-0.015, -0.018] new deaths per 100K (adjusted *R*^2^ is 0.343 and *p <* 0.001), controlling for test rates, weather and mobility. For reference, the rate of COVID-19 outcomes was 25.4 new daily cases per 100K and 0.95 daily new deaths per 100K, suggesting that even a 1% increase in masking adherence can have a significant positive effect.

### 2.3 International 68-country mask adherence and attitudes

We expand the scope of our analysis and look at the effect of mask adherence and attitudes towards mask wearing internationally. We do so by using the novel survey-based dataset ‘COVID-19 Beliefs, Behaviors & Norms Survey’ which asks more than 150 questions (see [25] for the complete survey instrument) about COVID-19 to more than half a million online Facebook survey respondents in 51 languages from 68 countries. Because the 68 countries we include in our analysis are from different parts of the world and were in various stages of the pandemic (first wave, second waves, before mandates, after mandates, etc.) during the 8 waves over 4 months of observation, we expect that our estimates are unbiased by such confounders.

We focus on the two questions most relevant to the effect of masks on COVID-19 outcomes: a question about mask adherence: “Out of 100 people in your community, how many do you think wear a face mask or covering when they go out in public?”; and a question about mask norms: “Out of 100 people in your community, how many do you think believe the following because of COVID-19: People should wear a face mask or covering when out in public?”. The first question is a self-reported sample estimate of the percentage of a respondent’s community that wears masks in public, and the second question provides a sample estimate of the percentage of the community that *believes* masks to be important to wear in public.

We use a weighted survey regression approach to regress each country’s new daily deaths per 100K and cases per 100K (both smoothed) against the self-reported mask adherence and mask attitude with countries as survey strata and individual anonymized survey responses as survey clusters. Samples are expected to be representative because we use a unique weight for each sample which correct for a variety of biases including demographics (age bracket and sex) of the respondents compared to the census data in each country and compared to Facebook’s online population (through post-stratification), and for response and non-response drivers (the estimated design effects was below four as detailed in [25]). The dataset’s weighting strategy to obtain representative samples is described more in the Data section, and comprehensively described in their technical report [25]. We also control for mobility, new test rate, population density and the country’s human development index. The regression specification is described in equation 4.2.4.

As shown in Fig 5, the associated effect coefficients suggest that a 1% increase in mask adherence is associated with a decrease of -0.45 [-0.70,-0.29] cases per 100K and a decrease of -0.042 [-0.046,-0.037] deaths per 100K. Similarly, we find that a 1% increase in attitude about the importance of wearing masks leads to a decrease of -0.53 [-0.64,-0.42] cases per 100K and a decrease of -0.035 [-0.038,-0.032] deaths per 100K. For reference, the current statistics in the world as of December 14, 2020 is 10 daily new cases per 100K and 0.17 daily deaths per 100K, which suggest that even a 1% increase in mask adherence and attitudes is associated with a strong positive impact on COVID-19 outcomes worldwide.

**Figure 5:**
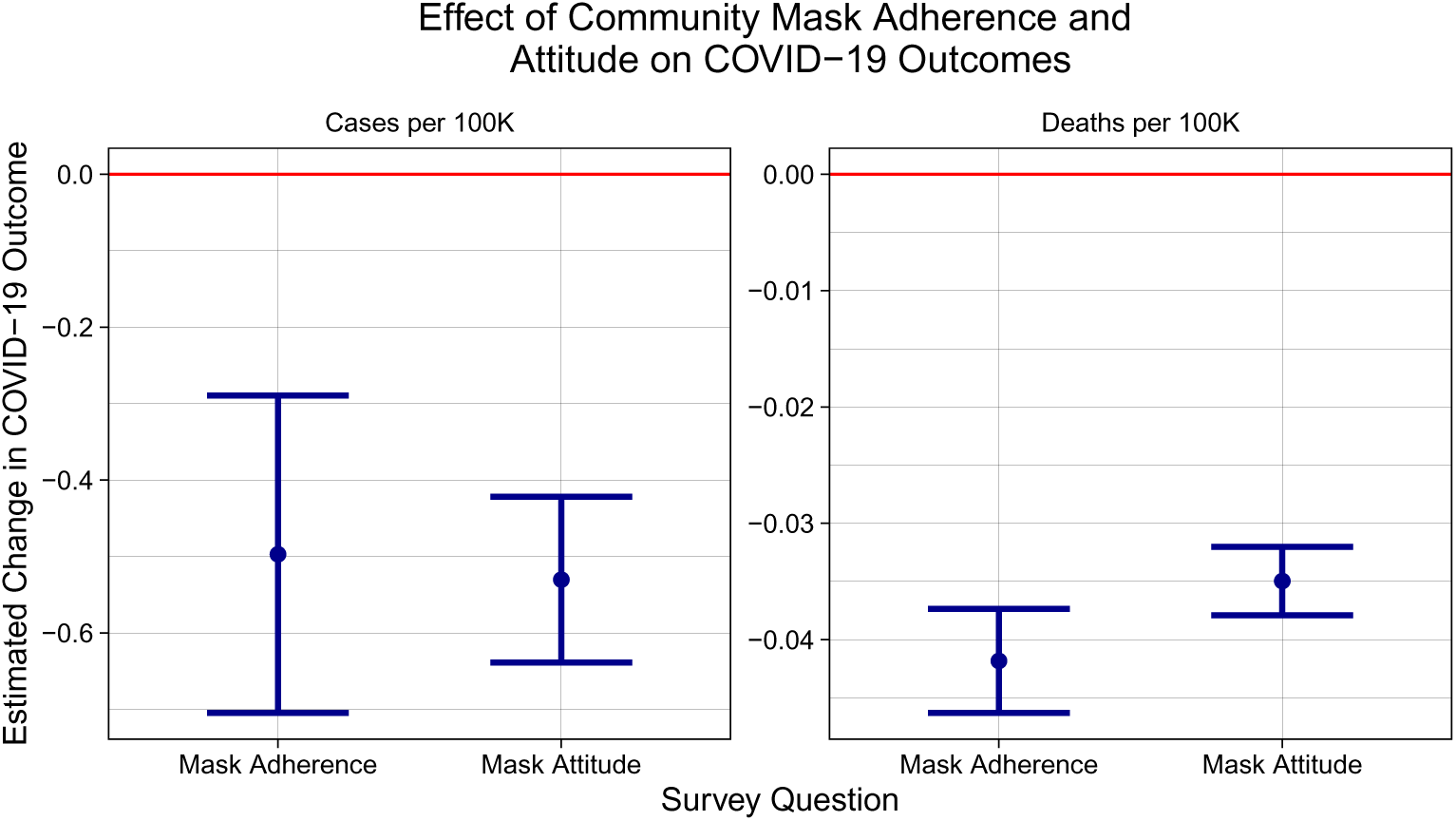
Impact of community mask adherence and attitudes. Using a novel survey-based dataset of more than half a million online Facebook survey respondents in 51 languages from 68 countries, we estimate that a 1% increase in community mask adherence is associated with a decrease of -0.45 [-0.70,-0.29] cases per 100K and a decrease of -0.042 [-0.046,-0.037] deaths per 100K. Similarly, we find that a 1% increase in community attitude about the importance of wearing masks leads to a decrease of -0.53 [-0.64,-0.42] cases per 100K and a decrease of -0.035 [-0.038,-0.032] deaths per 100K. For reference, the current COVID-19 outcomes in the world as of December 14, 2020 is 10 daily new cases per 100K and 0.17 daily deaths per 100K, which suggest that mask adherence and attitudes are associated with a strong positive impact on COVID-19 outcomes worldwide. Samples are expected to be representative because we use a unique weight for each sample which corrects for a variety of biases.

As a robustness check, we perform the same weighted survey regression but this time dis-aggregating by survey waves. As shown in supplementary Fig S11, the regression coefficients of mask adherence and attitudes on deaths and cases are negative or indistinguishable from zero, which supports our results that mask adherence and attitudes have a positive impact on COVID-19 cases and deaths across waves.

## 3 Discussion

We extend previous studies by investigating an unprecedented breadth and depth of outcomes, geographical resolutions, types of mask mandates, early versus later waves, and control variables such as mobility, weather, test rate, and other government interventions (such as stay-at-home orders). We find that, overwhelmingly, mask mandates are associated with a decrease in cases, hospitalizations and deaths. Additionally, we observe that mask mandates are associated with an increase in mask adherence in the 4 states where adherence data is present before and after the introduction of a mask mandate. Using a novel dataset of surveys from 68 countries across a variety of pandemic phases and pre/post mask mandate stages, we also observe that community mask adherence (how many people in a community wear masks) and community mask attitudes (how many people in a community believe that masks are an important for COVID-19) are associated with a decrease in cases and deaths.

Our results have policy implications for reinforcing the need to maintain and encourage mask-wearing by the public. These results are especially significant in light of some states starting to remove their mask man-dates and attempts by state governments to prevent local governments from implementing mask orders, even though they are associated with a minimal economic impact [45]. Generally, our results are relevant to the ongoing effort to determine how to best design, implement and sustain the adoption of non-pharmaceutical interventions (NPIs) to curb the spread of the COVID-19 pandemic [46–51].

Even though our results, taken together, strongly suggest the positive effect of mask mandates, mask adherence and mask attitudes on COVID-19 cases, hospitalizations and deaths, an important open question is the investigation of the optimal incentives and regulatory mechanisms to increase mask-adherence [18,52–56] especially in a climate where conspiracy theories are emerging to discourage mask-adherence [57]. More research is also needed to probe improvements in public health communication strategies to encourage mask-adherence, including harm-reduction frameworks [58] and policies tailored to help disadvantaged communities’ lived experiences [59]. Lessons can be taken from successful approaches in other public health crises such as HIV [60–63].

Our results suffer from the usual limitations of econometrics-based studies: although we control for many confounders such as other NPIs (stay-at-home and non-essential business closures), mobility, weather and test rate, it is possible that unexplained exogeneity and endogeneity persists. For example, it is possible that our treatment effects are underestimates due to the peer effects caused by neighboring states’ mask mandates affecting other states [36]. Once outcome data disaggregated by demographics (e.g. age, race, gender) is available at the state or county level, an interesting question for future work is to investigate the differential impact of mask mandates over these demographic groups. In our study of mask mandates on mask adherence, ongoing efforts using computer vision to estimate mask adherence from social media images and videos [38–41] should soon provide more detailed data on mask adherence, hopefully starting from the beginning of the pandemic. Another limitation is that although our survey-based sample estimates of mask adherence and mask attitudes in the 68-country dataset include weights to correct for a number of biases, it is possible that there is some bias remains although this is unlikely due to their small estimated instrument design effect [25].

We leave to future work the important design and estimation around these limitations but expect that our findings will still provide new insights into the long-term effect of mask mandates on mask adherence, COVID-19 cases, hospitalizations and deaths, and introduces the importance of community mask adherence and community mask attitudes.

## 4 Materials and Methods

We obtain data from a large variety of publicly available sources which we detail in the Data section below. We collected no data for this study. Our research received MIT IRB Exempt Status E-2578 and E-2859.

The Analysis section next describes how we produced our three main results. To ensure the reproducibility of our results, we have shared the code and data used to perform data processing, analysis and plotting on Github.

### 4.1 Data

#### 4.1.1 Mask mandate start and end dates and types

To identify the start and, if any, the end of state-level mask mandates, we used two publicly available state-level mask mandate datasets [64, 65] and followed their references to official state government memos to perform accuracy checks. We found the datasets to be highly accurate even though the specifics of how and when mandates were instituted varies strongly between states. We investigated three specifications of mandates to use as our treatment: mandates requiring only business employees to wear masks, mandates for the general public, or whichever came first between mandates for business employees and the public. For the state-level analysis, we select only mandates that are at the state-level that require mask use as opposed to county-level mask mandates or using mandates that merely encouraging people to wear masks. The table of mask mandate start and end dates is shown in supplementary table A.3.

In order to investigate county-level mandate effect heterogeneity within a state, we investigate the effect of county-level mask mandates. We use a publicly available dataset [35] which we similarly verify for accuracy.

#### 4.1.2 Control NPIs: Stay-at-home and non-essential business closures intervention

Mask mandates were often introduced around the same time that other non-pharmaceutical interventions (NPI) such as stay-at-home orders and non-essential business closures were implemented. In order to estimate the effect of mask mandates without the bias of other NPI’s, we use the same public datasets [64, 65] to obtain the start and end dates of stay-at-home and non-essential business closures (the two broadest and far-reaching NPIs), which we also verify for accuracy.

#### 4.1.3 COVID-19 Cases, Hospitalizations and Deaths Data

State and county-level COVID-19 outcome data was obtained from the CMU Delphi project [42]. Specifically, we use as outcomes the daily new confirmed cases per 100K population, new confirmed deaths per 100K population, percentage of new hospital admissions with COVID-associated diagnoses, and the percentage of outpatient doctor visits primarily about COVID-related symptoms.

It is important to note that we only observed the *proportion* of hospitalizations, which is the number of new hospitalizations due to COVID-19 relative to the number of total admissions. Unfortunately, this proportion is a possibly noisy proxy for the number of hospitalizations per 100K due to COVID-19 (that we would like to observe) as it is well documented that hospital capacity was deliberately increased to accommodate surges in the number of hospitalizations and that fewer people went to the hospital as regular admissions were discouraged and elective treatments postponed during the pandemic [66–69] which affects the denominator of the hospitalization proportion. However, even though possibly noisy, we investigate the effect of mandates on the proportion of hospitalization as it still provides a useful perspective of the effect of mask mandates on COVID-19 outcomes.

We use the smoothed outcomes measures using 7-day averages and with systematic day-of-week effects removed. Our outcome data cover all 50 states in addition to the District of Columbia and runs from Feb 2, 2020 to September 27, 2020 (we cannot obtain data past September for some outcomes such as hospitalization). 43 states and the District of Columbia had a required state-level mask mandates during this period of data. For the county-level dataset, we have 857 counties for which we have outcome and control variable data. This is because county-level datasets are currently not available due to their higher resolution. However, our 857 counties cover a large portion of the country - 77% of the US population as shown in supplementary Fig S3 - and are therefore representative.

We make sure not to use any outcome data that was after a state or county’s mask mandate was lifted to prevent end-of-treatment effects from biasing our treatment estimates, and we only use the first mask mandate start and end dates in cases where there were multiple waves of mandates over the year. Overall, this dataset contains 10,078 data points used for the state-level event study (described in the next section) and 77,930 data points for the county-level event study.

#### 4.1.4 Mask Adherence Data

State-level mask adherence data (how much people are wearing face masks) became available on September 8, 2020 from the Delphi project [42]. This data was collected by them via online surveys of more than 1 million U.S. residents and we use their weighted state-level estimates as our indicator of state-level mask adherence. Their survey weighing [43] accounts for various factors including adjusting estimates so that they are representative of the US population according to a state-by-age-gender stratification, adjusting for the US Facebook user population, and adjusting for the propensity of a Facebook user to take the survey. For the regression of the effect of mask mandates on adherence, we have 323 data points for the four states, and for the regression of adherence on COVID-19 cases and deaths, we have 3,841 data points for all states (including D.C.). County-level mask mandate data is only available until August 4, 2020 which is before mask adherence data starts (September 8, 2020), making the use and investigation of county level adherence data not possible.

#### 4.1.5 International Mask Norm and Adherence Data

International mask norm and adherence data was obtained from the COVID-19 Beliefs, Behaviors and Norms Survey [25] which collected more half a million online Facebook survey responses in 51 languages, from 68 countries. The survey was deployed in 8 waves starting on July 7, 2020 and is still on-going. Each survey response comes with a weight that allows for representativeness of estimates at the country and wave-level. Weights are calculated to account for a variety of biases including demographics (age bracket and gender) of the respondents compared to the census data in each country (through post-stratification), and response or non-response behaviors. The reported design effects due to both the non-response and post-stratification weighting was below four. In our analysis, we only use survey responses where the full survey was completed (including the demographic part) and where the mask-related questions we use in our analysis were completed. Overall, we have 479,917 data points covering 8 waves of survey deployment. The survey dataset (de-identified) is available to academic and non-profit researchers upon completion of the Facebook Data Use Agreement.

#### 4.1.6 Controls variables

We control for a large number of variables in our analyses: we use restaurant and recreation, grocery and pharmacy, parks, transit stations, workplace and residential percentage change from baseline (using the median day-of-the-week value from the first 5 weeks of January and February) mobility data from Google’s COVID-19 Community Mobility Reports [70]; new tests per 100K population from the COVID Tracking Project [71], and temperature and precipitation data from NOAA’s Weather and Climate Toolkit [72]. For international mask norm and adherence, we obtain COVID-19 country-level confirmed cases and confirmed deaths from the Our World in Data initiative [73].

Because we also include the squared values of each mobility indicator as a proxy for social contacts ( [32, 33]), we first re-scale the Google mobility index [70] as such: re-scaled mobility = (mobility + 100)/100. This is done to prevent the issue of Google’s the square of negative mobility indicator values to be the same as the positive values.

### 4.2 Analysis

#### 4.2.1 Event study framework

We build on recent work using event study designs to estimate treatment effects in the context of the COVID-19 pandemic: the effect of NPIs (e.g. business closures and stay-at-home orders) on the volume of online searches related to unemployment [74], on social distancing in the US [75], on the airline industry [76], on stock markets worldwide [77–80], and on COVID-19 cases at the county level [81].

An event study design allows us to estimate the treatment effect associated with mask mandates on COVID-19 outcomes on each day following the introduction of the mandate relative to the day prior to its introduction. Our geographical unit of analysis are states to minimize peer-effects of neighboring counties’ mandates due to the underlying interdependence between county mobility patterns [36], as well as the fact that people living in one county often have to travel to a different county to get medical care, resulting in inconsistent accounting of COVID-19 health outcomes at the county level [37]. By contrast, patients rarely cross state borders to get medical help as health insurance in the US is predominantly state-based. We include a robustness check by running our analysis at the county-level using a dataset of county-level only mask mandates [35] to investigate county-level heterogeneity.

The event study framework is well-suited for our purposes as it allows for the same state to be used both as treatment and control based on the timing of a mandate, which is critical because states introduced mask mandates at different dates over the course of the pandemic [74]. While the first mandates were implemented in April, the latest ones occurred in November, some states still having had no mandate as of this writing (see table A.3).

#### 4.2.2 Event study specification

Our specification is:

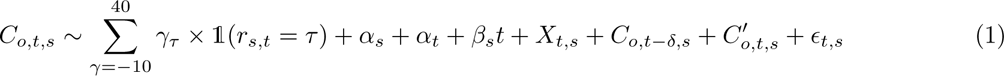

The outcome is *C_o,t,s_* (*o* ∈ {new deaths due to COVID-19, new confirmed cases of COVID-19, hospitalization proportion due to COVID-19}) for each day *t* in state (or county when investigating county-level mandates) *s*. We make sure to only use new cases, hospital admissions and deaths each day, as opposed to cumulative numbers. Because some states such as New York had a much higher COVID-19 incidence than other states, we used population-scaled (per hundred thousand people) outcomes, and we further Z-normalize the daily value of each cases and deaths using state-specific means and standard deviations (hospitalization is already a percentage and therefore does not require normalization). Due to these transformations, our treatment effect estimates are expressed in units of standard deviations. We also ensured that all our variables have been smoothed and de-trended (for day of the week variations). *∈_t,s_* represent the model residuals.

The goal is to estimate treatment effects *γ_τ_* where *τ* is the number of days before or after the start of a mask mandate. 1 is the delta function such that *γ_τ_* is non-zero only when the number of days *r_s,t_* relative to the start of mask implementation is *τ*. As is standard in an event study framework [74], we normalize *γ_τ_* _=_*_−_*_1_ = 0 which means that our estimated treatment effects are to be interpreted relative to the day before the introduction of a mask mandate. We set the range of days *r_s,t_* before or after the introduction of mask mandates to be from -10 to 40 because choosing values earlier than -10 leads to some state no longer having enough data (as they implemented mandates very soon after COVID-19 outcome data was available) and 40 to account for reporting delays and the pathogenesis characteristics of the virus as the interquartile range of the time between symptom onset and death is 8-26 days [82–89]. The clock time interval for our data is February 1 to September 27, 2020, the time during which data for all outcomes and controls was available.

*α_s_* and *α_t_* are state and time fixed effects, *β_s_t* is a state-specific trend coefficient multiplying the time trend variable to allow for the fact that different states had different outcome trajectories (see [90] for more details). We report standard errors clustered at the state-level.

*X_t,s_* are our controls for a number of confounding factors: (a) indicators of human mobility [70] (number of visits to recreational areas, grocery stores, pharmacies, parks, transit stations, workplace and residential areas) to control for variations in the spread of the virus due to variations in mobility and as a proxy for other NPIs such as stay-at-homes (as they impact mobility); (b) daily new test rate [71] to control for variations in the number of tests being administered; (c) the start and end dates of stay-at-home and non-essential business closures (the two broadest and far-reaching NPIs) to estimate the effect of mask mandates without the bias of other NPI’s; and (d) temperature and precipitation [72] to control for weather-induced behaviors.

We also include a time-delayed (*δ* = two weeks) outcome *C_o,t−δ,s_* and a growth rate (over the past two weeks) 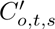 because conditioning on the delayed outcome and growth rate results in the confounding path due to behavior change to be blocked based on the general causal graphical model of [21]. A delay of two weeks was chosen based on both our sensitivity analysis and that the estimated behavioral lag in the estimation of the causal graphical model of [21]. Additionally, as will be shown in the results section, shorter or longer delays would be at odds with the pathogenesis characteristics of the virus [82–89]. As a robustness check, we explore the effect of including or not delays on the event study estimation in Fig S1.

While political affiliation has been shown to correlate the propensity to wear a mask [91], this effect should captured in the state fixed effect since we do not expect political affiliation to change significantly during our observational period.

We use the R package’s `lfe`’s regression function `felm` to run this specification as it uses the method of alternating projections to speed up regression on our large datasets [92].

As a robustness check, we repeat employ a similar event study specification to verify that the decrease in COVID-19 outcomes we observe is not due to a decrease in test rate (per 100K of population. We use the following specification:

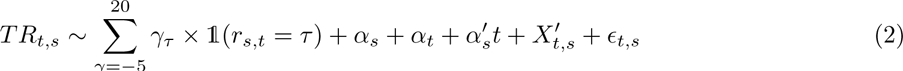

where *T R_t,s_* is the daily new test rate, and all other variables are similar to above, with 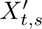 being our controls without test rate (since it is now the dependent variable).

#### 4.2.3 Mask Mandate and Mask Adherence

Using daily mask adherence data from September 8 to November 30, 2020, we estimate a similar event study specification to the above using mask adherence *A_s,t_* as the dependent variable. Additionally, we control for deaths and confirmed cases because we want to control for the fact that the more case and deaths people see, the more they might be likely to wear masks (although we do not observe much of a difference when omitting them). Again, we control for a number of factors: testing rate due to its potential effect on mask usage as more people might wear masks if they are more regularly tested, and mobility as we want to control for the fact that perhaps the more people leave their house, the more they might wear masks.

We implement a multi-linear regression of COVID-19 outcomes on compliance, controlled by a number of factors, as specified below:

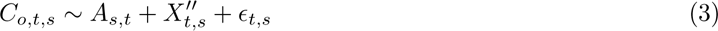

where *C_o,t,s_* are the daily *t* COVID-19 outcomes *o* ∈ {deaths, cases} and 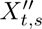 are the mobility, new test rate and weather controls.

#### 4.2.4 International Mask Adherence and Norms

Here, we use multi-linear regression to estimate the effect of mask norms or mask adherence on COVID-19 case and death outcomes for each wave *w*:

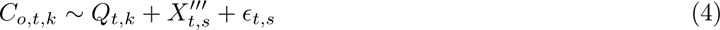

where *C_o,t,k_* are the daily *t* COVID-19 outcomes *o* ∈ {deaths, cases} for each country *k*, for question *q*, controlling for mobility and new test rate 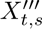. One separate model estimation is run for each wave *w*, each outcome *o* and each question *q ∈* {mask norms, mask adherence}.

More details about survey weights and the dataset can be found in the 4.1 section. We also verify that the weighted average mask adherence, weighted average mask attitude and total number of responses did not change significantly over time during the survey as shown in supplementary figures S9, S10 and S8.

We use a complex survey regression (using R’s survey package) with countries as survey strata and individual anonymized survey responses as clusters. We only select responses that finished the whole survey (including several attention check questions), and we remove responses with missing values for the questions under consideration, and also those with missing weights (*<*1% of responses).

The 68 countries in our dataset are: United Kingdom, Poland, Italy, Germany, Japan, Argentina, France, Turkey, Mexico, Colombia, United States, Pakistan, Romania, Indonesia, Philippines, Egypt, Malaysia, Bangladesh, Nigeria, Brazil, Thailand, Vietnam, India, Netherlands, Azerbaijan, Australia, South Africa, Canada, Estonia, Senegal, Afghanistan, Tanzania, Angola, Ecuador, Georgia, Mongolia, Peru, Algeria, Mozambique, Bolivia, Portugal, Iraq, Cameroon, Morocco, South Korea, Uruguay, Honduras, Nepal, Sri Lanka, United Arab Emirates, Spain, Cote d’Ivoire, Myanmar, Chile, Venezuela, Guatemala, Trinidad & Tobago, Sudan, Kenya, Jamaica, Ghana, Uganda, Ukraine, Taiwan, Singapore, Cambodia, and Kazakhstan.

## Data Availability

All data and code are available at https://github.com/d-val/Mask_Mandates_Adherence_Attitudes

## A Supplementary

### A.1 Characteristics of COVID-19 outcome and adherence data

**Table S1:** Characteristics of COVID-19 outcome variables. For the proportion of daily hospitalization admissions due to COVID-19, admissions are coded as COVID-19 related if the admission code U071, U072, B9729, or if primary ICD-10 code is R05, R060, R509, Z9911, R0902, R0603, R0609, R062, R069, R0602, R05, R0600, J9691, J9692, J9621, J9690, J9601, J9600, J189, J22, J1289, J129, J1281, B9721, B9732, B342, B349, A419, R531 or R6889 and there is a secondary ICD-10 code of Z20828, or if the primary ICD-10 code is Z20828.

|  | Outcome |  |  |  |
| --- | --- | --- | --- | --- |
|  | New confirmed COVID-19 cases | New confirmed deaths due to COVID-19 | % of new hospital admissions with COVID-associated diagnoses | % of people who wore a mask for most or all of the time while in public in the past 5 days |
| **Source** | Delphi COVIDcast Epidata API | Delphi COVIDcast Epidata API | Delphi COVIDcast Epidata API | Delphi COVIDcast Epidata API |
| **Frequency** | Daily | Daily | Daily | Daily |
| **Geographical Resolution** | State-level | State-level | State-level | State-level |
| **Geographical Coverage** | All States and D.C. | All States and D.C. | All States and D.C. | Hawaii, Iowa, North Dakota, New Hampshire |
| **Date Range** | Feb. 1 - Sep 27 2020 | Feb. 1 - Sep 27 2020 | Feb. 1 - Sep 27 2020 | Sep 8 - Nov 27, 2020 |
| **Smoothing** | 7-day average signal | 7-day average signal | Systematic day-of-week effects removed | Seven day pooling |
| **Adjusted** | Per 100K population | Per 100K population | Per total admissions that day | Weighted to correct for a variety of biases |
| **Normalization** | Z-scored state-specific means and standard deviations | Z-scored state-specific means and standard deviations | Not Needed (already normalized as a percentage) | Z-scored state-specific means and standard deviations |

### A.2 Characteristics of COVID-19 control data

**Table S2:** Characteristics of control variables

|  | Source | Freq. | Geograph.<br>Resolution | Date Range | Normalization |
| --- | --- | --- | --- | --- | --- |
| **Retail+recreation mobility** | Google Mobility Reports | Daily | State-level | Feb 1-Sep 27 2020 | Median day of the week baseline |
| **Grocery+pharmacy mobility** | Google Mobility Reports | Daily | State-level | Feb 1-Sep 27 2020 | Median day of the week baseline |
| **Parks mobility** | Google Mobility Reports | Daily | State-level | Feb 1-Sep 27 2020 | Median day of the week baseline |
| **Transit stations mobility** | Google Mobility Reports | Daily | State-level | Feb 1-Sep 27 2020 | Median day of the week baseline |
| **Workplaces mobility** | Google Mobility Reports | Daily | State-level | Feb 1-Sep 27 2020 | Median day of the week baseline |
| **Residential mobility** | Google Mobility Reports | Daily | State-level | Feb 1-Sep 27 2020 | Median day of the week baseline |
| **Temperature** | NOAA | Daily | State-level | Feb 1-Sep 27 2020 | Avg daily temperature |
| **Precipitation** | NOAA | Daily | State-level | Feb 1-Sep 27 2020 | Avg daily precipitation |
| **Population Density** | Oxford World in Data | Daily | State-level | Feb 1-Sep 27 2020 | Per square. km |
| **Human Develop. Index** | Oxford World in Data | Daily | Country-level | Feb 1-Sep 27 2020 | Relative to UNDP limits |
| **New Test Rate** | COVID Tracking Project | Daily | Country-level | Feb 1-Sep 27 2020 | Per 100K population |

### A.3 State-level Mask Mandates Dates

**Table S3:**
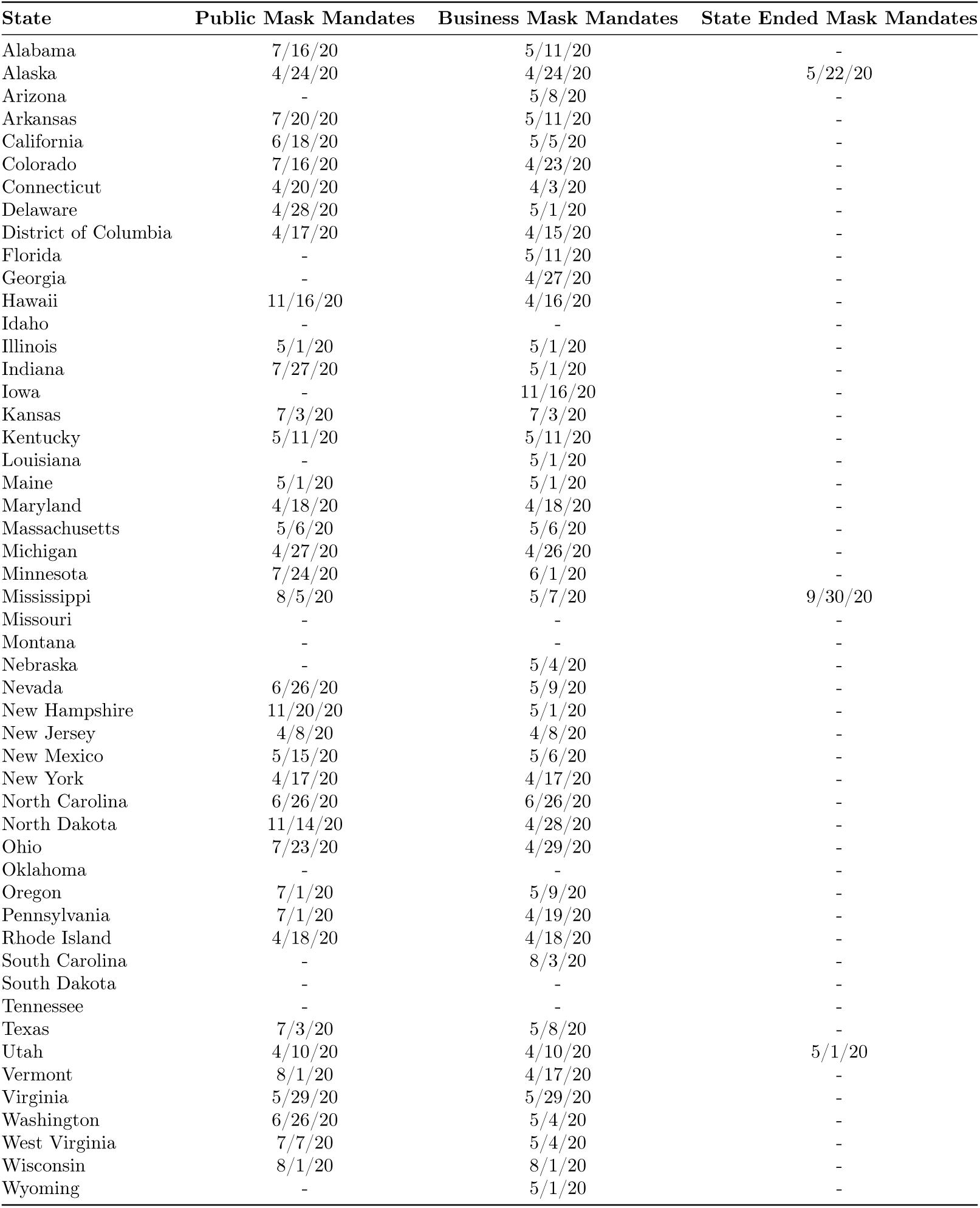
Mask mandate start and end dates as of Nov 30, 2020

| State | Public Mask Mandates | Business Mask Mandates | State Ended Mask Mandates |
| --- | --- | --- | --- |
| Alabama | 7/16/20 | 5/11/20 | - |
| Alaska | 4/24/20 | 4/24/20 | 5/22/20 |
| Arizona | - | 5/8/20 | - |
| Arkansas | 7/20/20 | 5/11/20 | - |
| California | 6/18/20 | 5/5/20 | - |
| Colorado | 7/16/20 | 4/23/20 | - |
| Connecticut | 4/20/20 | 4/3/20 | - |
| Delaware | 4/28/20 | 5/1/20 | - |
| District of Columbia | 4/17/20 | 4/15/20 | - |
| Florida | - | 5/11/20 | - |
| Georgia | - | 4/27/20 | - |
| Hawaii | 11/16/20 | 4/16/20 | - |
| Idaho | - | - | - |
| Illinois | 5/1/20 | 5/1/20 | - |
| Indiana | 7/27/20 | 5/1/20 | - |
| Iowa | - | 11/16/20 | - |
| Kansas | 7/3/20 | 7/3/20 | - |
| Kentucky | 5/11/20 | 5/11/20 | - |
| Louisiana | - | 5/1/20 | - |
| Maine | 5/1/20 | 5/1/20 | - |
| Maryland | 4/18/20 | 4/18/20 | - |
| Massachusetts | 5/6/20 | 5/6/20 | - |
| Michigan | 4/27/20 | 4/26/20 | - |
| Minnesota | 7/24/20 | 6/1/20 | - |
| Mississippi | 8/5/20 | 5/7/20 | 9/30/20 |
| Missouri | - | - | - |
| Montana | - | - | - |
| Nebraska | - | 5/4/20 | - |
| Nevada | 6/26/20 | 5/9/20 | - |
| New Hampshire | 11/20/20 | 5/1/20 | - |
| New Jersey | 4/8/20 | 4/8/20 | - |
| New Mexico | 5/15/20 | 5/6/20 | - |
| New York | 4/17/20 | 4/17/20 | - |
| North Carolina | 6/26/20 | 6/26/20 | - |
| North Dakota | 11/14/20 | 4/28/20 | - |
| Ohio | 7/23/20 | 4/29/20 | - |
| Oklahoma | - | - | - |
| Oregon | 7/1/20 | 5/9/20 | - |
| Pennsylvania | 7/1/20 | 4/19/20 | - |
| Rhode Island | 4/18/20 | 4/18/20 | - |
| South Carolina | - | 8/3/20 | - |
| South Dakota | - | - | - |
| Tennessee | - | - | - |
| Texas | 7/3/20 | 5/8/20 | - |
| Utah | 4/10/20 | 4/10/20 | 5/1/20 |
| Vermont | 8/1/20 | 4/17/20 | - |
| Virginia | 5/29/20 | 5/29/20 | - |
| Washington | 6/26/20 | 5/4/20 | - |
| West Virginia | 7/7/20 | 5/4/20 | - |
| Wisconsin | 8/1/20 | 8/1/20 | - |
| Wyoming | - | 5/1/20 | - |

### A.4 Comparing controlling for outcome delay and growth

**Figure S1:**
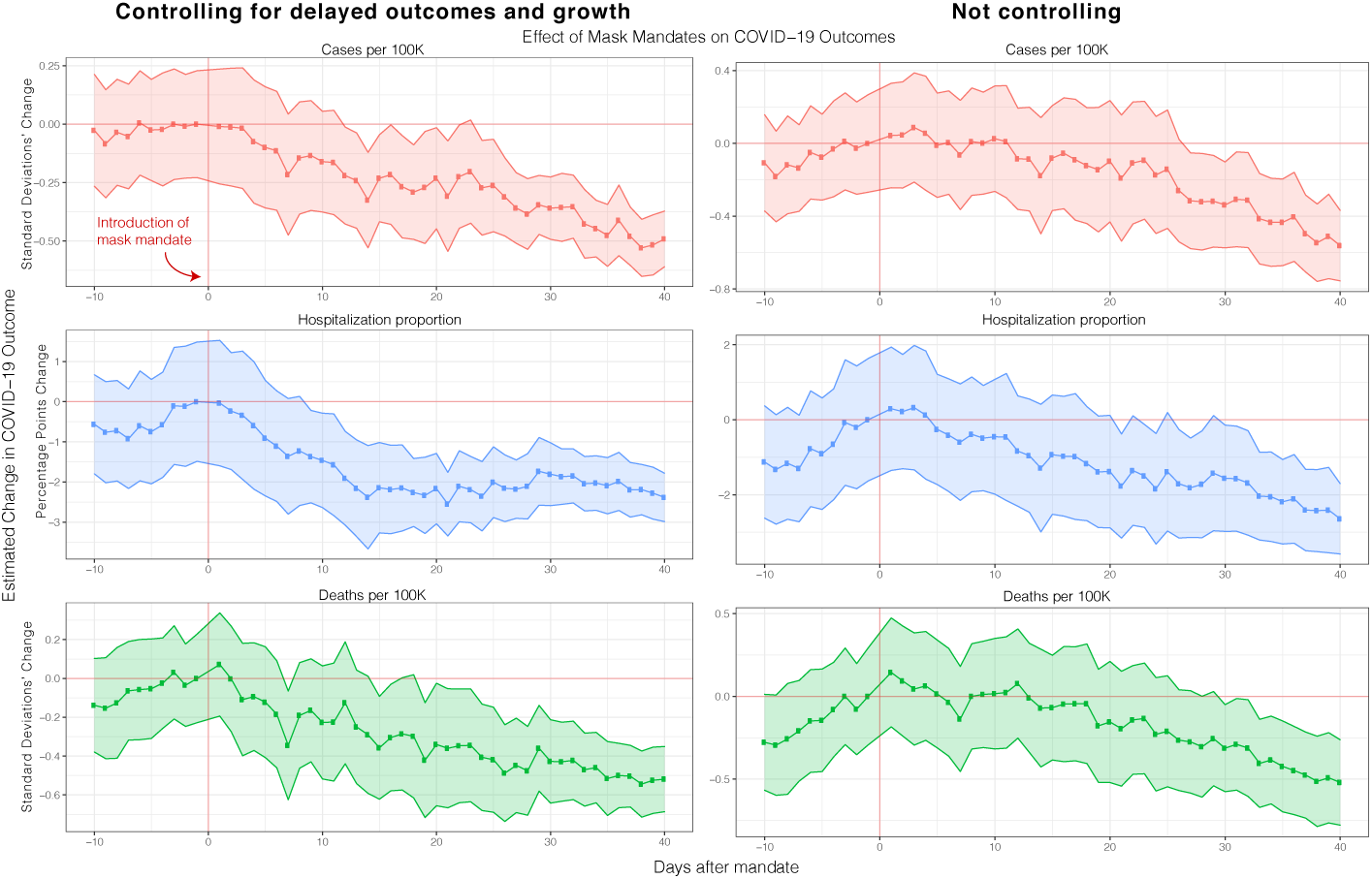
Robustness check of main event study results (section 2.1 fig. 1) where we compare controlling for past outcome values (with a delay of 14 days) and growth rates as a way to minimize confounding due to peoples private behavioral changes to COVID-19 [21]. As can be see in the figure, the overall trajectory and magnitude of the treatment effect is consistent with our previous main result. Full regression results are shown in table S8.

### A.5 Testing rate

**Figure S2:**
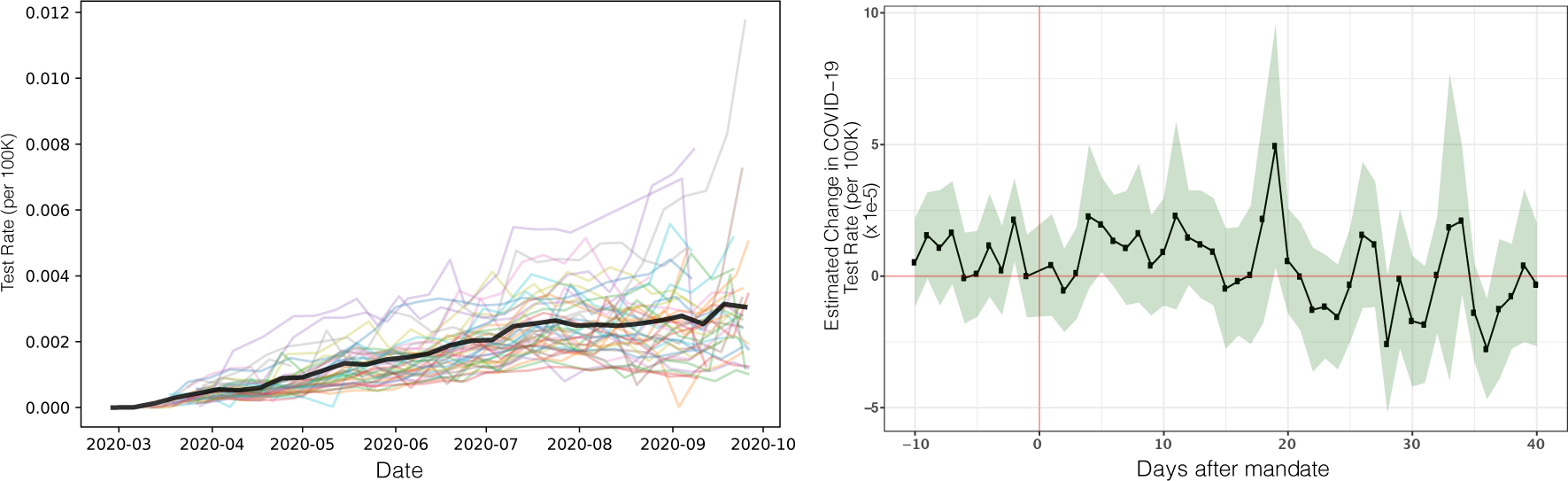
**Left:** Per state (each state a different color) and average (over all states, thicker black line) testing rate increased during our period of investigation. **Right:** Event study of testing rate over all states controlling for a number of factors showing that mask mandates are not associated with changes in testing rate during our period of investigation. This suggests that testing rate decreases are not behind the decrease in COVID-19 outcomes we observe after the introduction of mask mandates. Full regression results are shown in table S12.

### A.6 County-level heterogeneity

**Figure S3:**
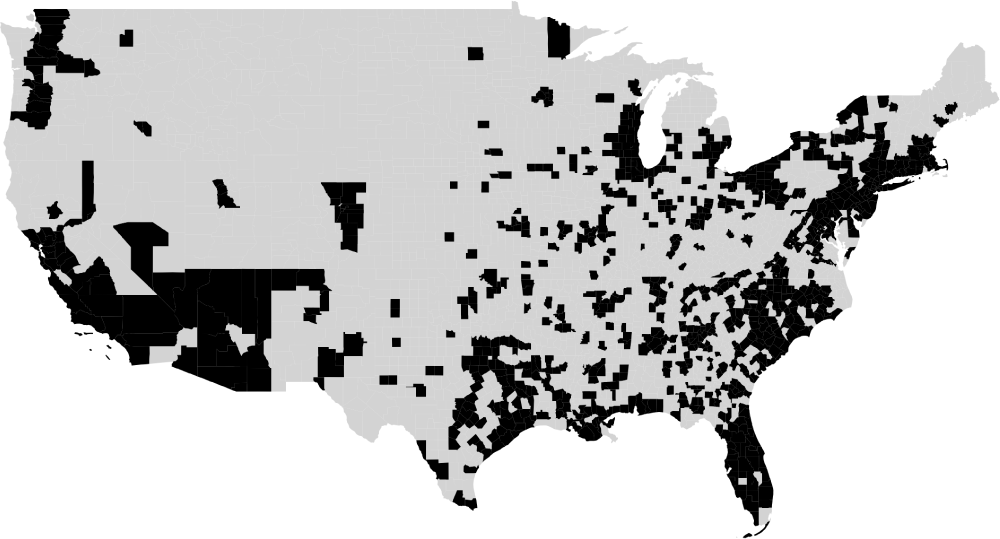
Map showing the 857 counties for which we have outcome and control variable data. Missing counties either had too few cases (and therefore cannot be reported for privacy reasons) or had data available later than the end of a mask mandate ended. The counties we have data for contain 77% of the US population (as per the 2018 US Census).

**Figure S4:**
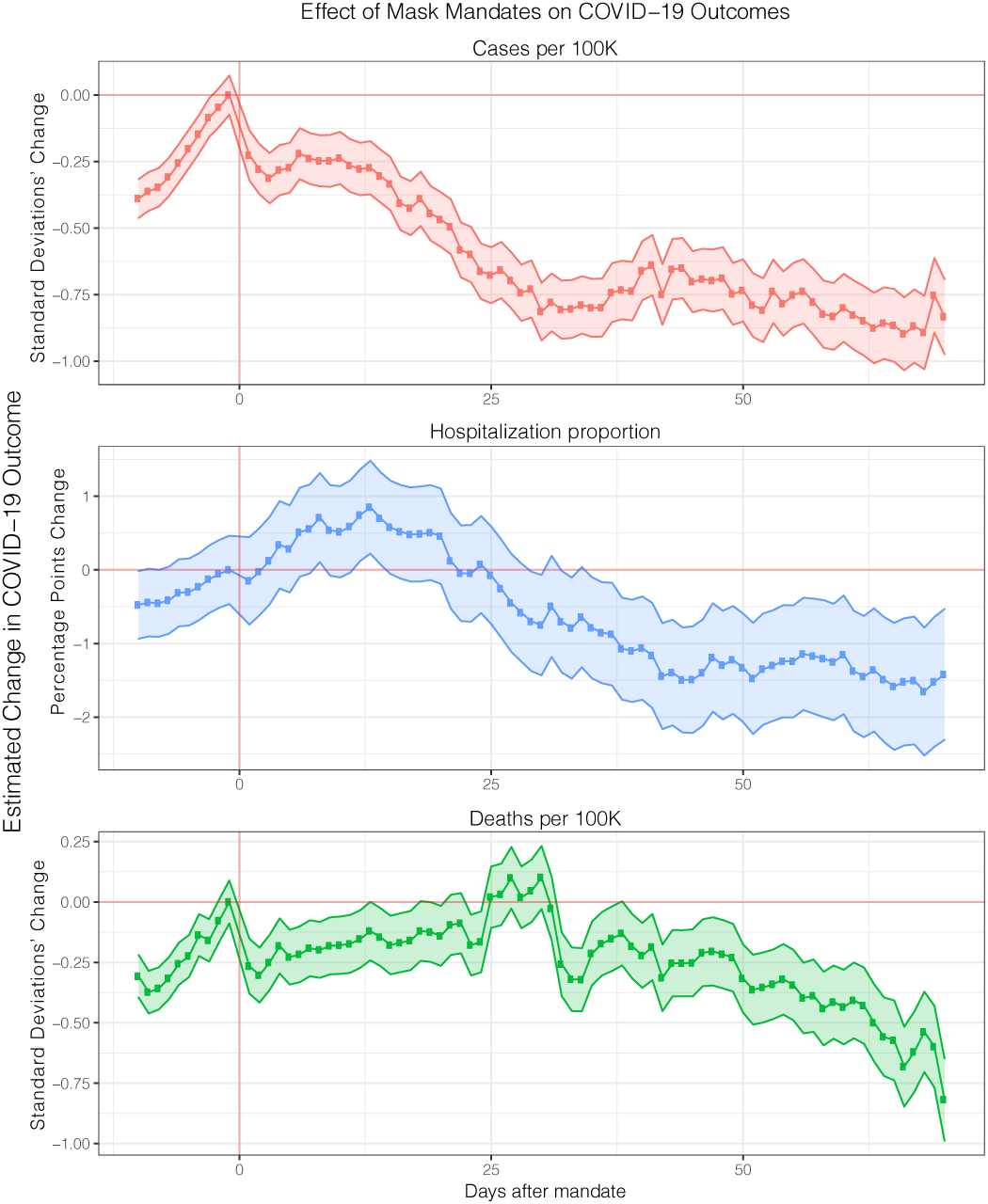
Robustness check of main event study results (section 2.1 Fig 1) where we focus on county level mandates only. Full regression results are shown in table S11.

### A.7 Pre-trends

**Figure S5:**
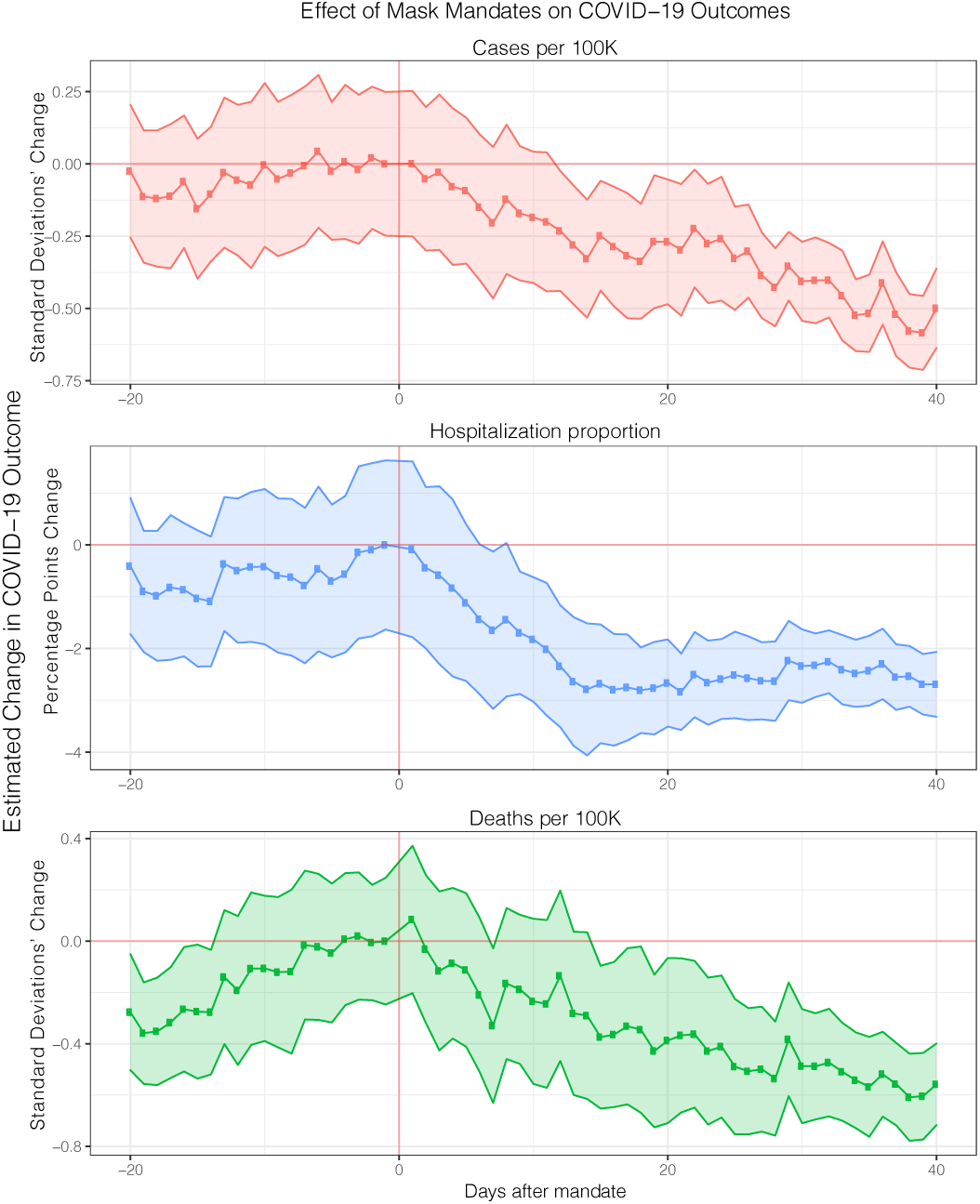
Robustness check of main event study results (section 2.1 Fig 1) where we investigate longer pre-treatment windows and no fixed effect.

### A.8 Controlling for confirmed cases

**Figure S6:**
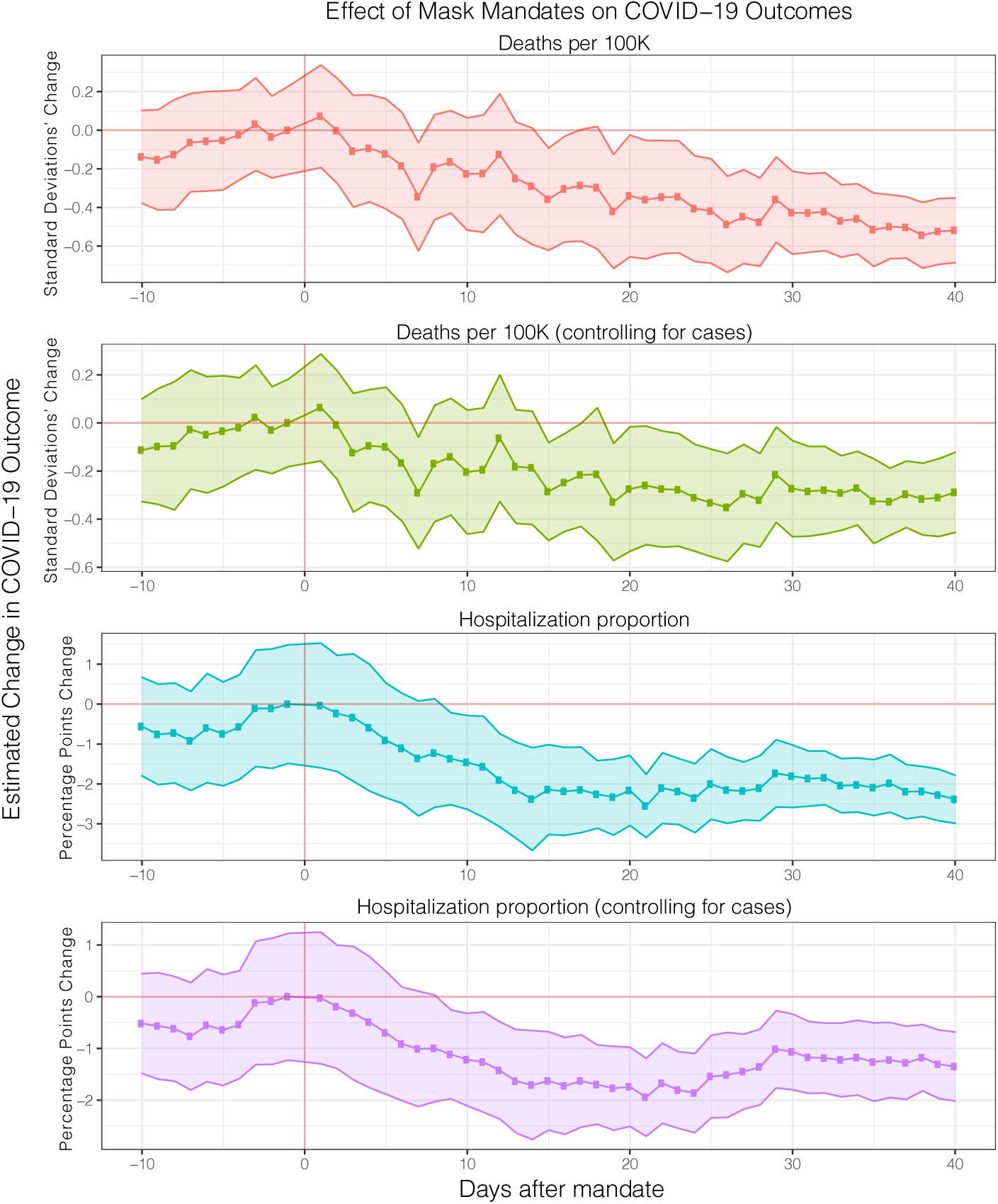
Robustness check of main event study results (section 2.1 Fig 1) where we compare the effect of mask mandates on deaths and hospitalization with and without controlling for cases. Because the trajectory and magnitude of the treatment effect is consistent with our previous main result under either specification (with and without controlling for cases), this demonstrates that our main result is not sensitive to whether we account for the cases underlying the more severe outcomes. Full regression results are shown in table S7.

### A.9 Mask Adherence following mask enforcement

**Figure S7:**
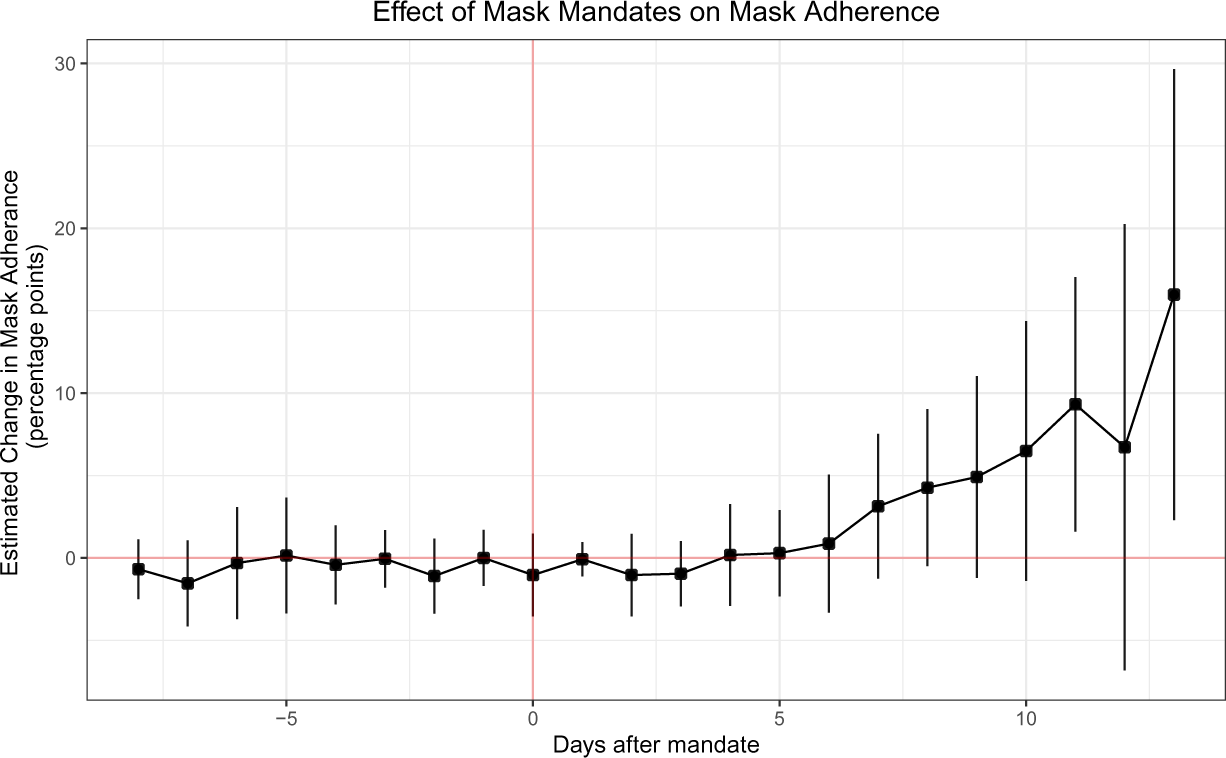
Event study of mask adherence when not controlling for cases, deaths and testing rate. As expected, the treatment effect becomes noisier but reamins consistent to when controlling for cases, deaths and testing rate. Full regression results are shown in table S13.

### A.10 International Community Mask Adherence and Attitudes

**Figure S8:**
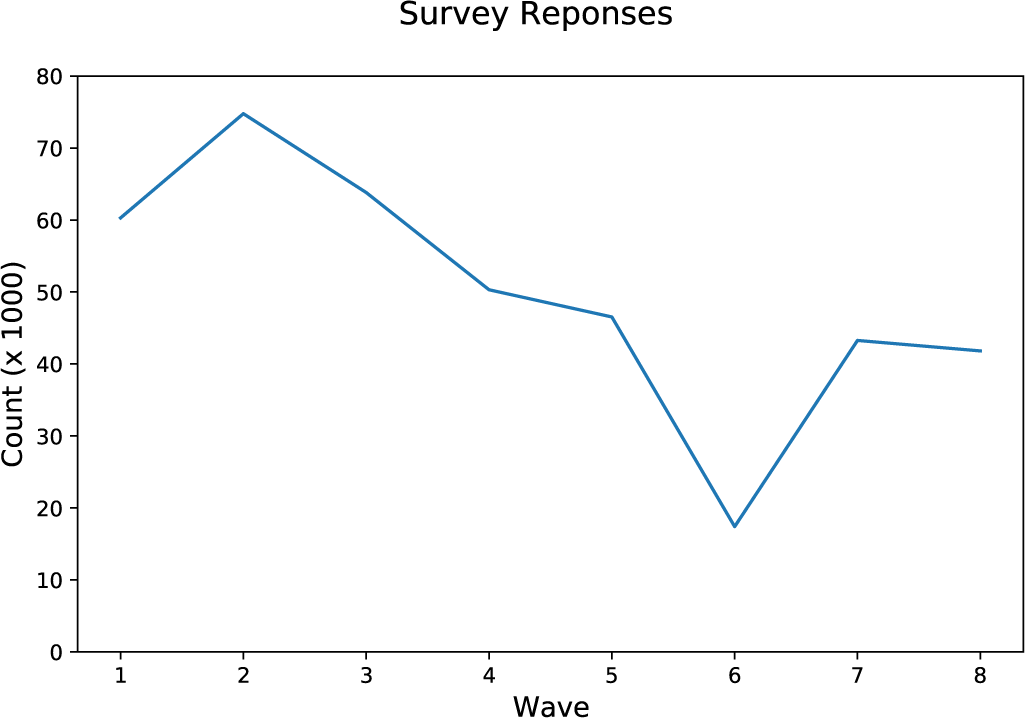
Number of responses per wave of survey.

**Figure S9:**
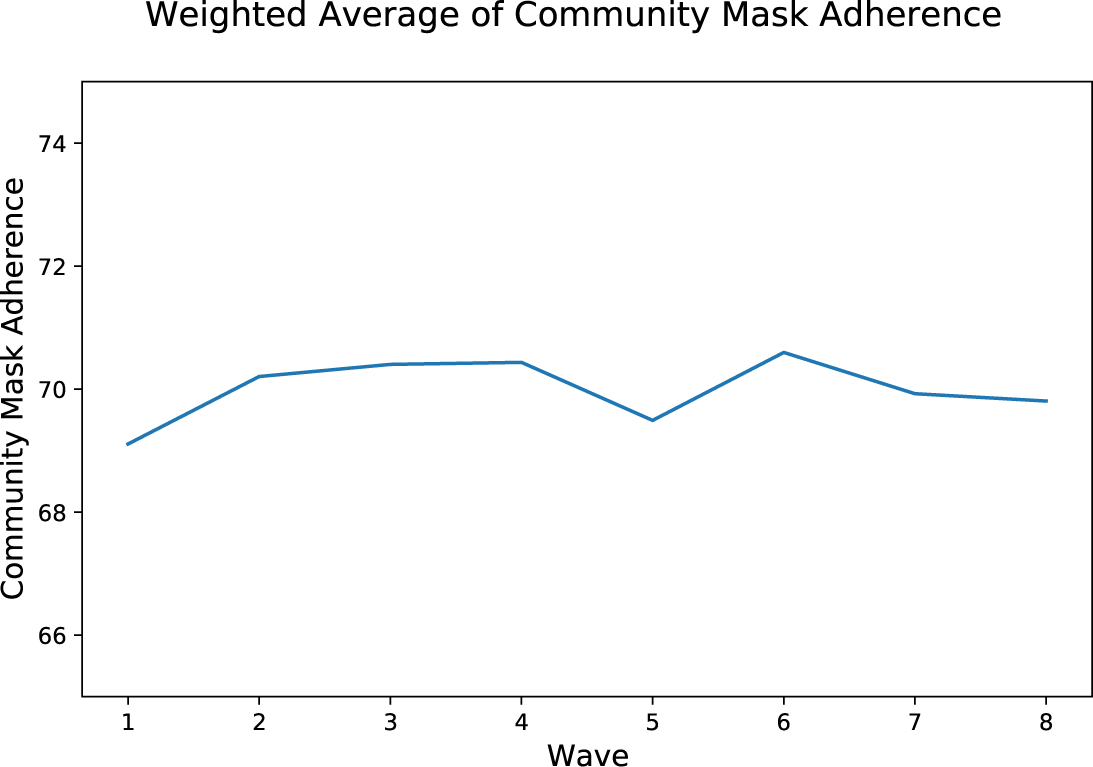
Average (weighted) community mask adherence response per wave.

**Figure S10:**
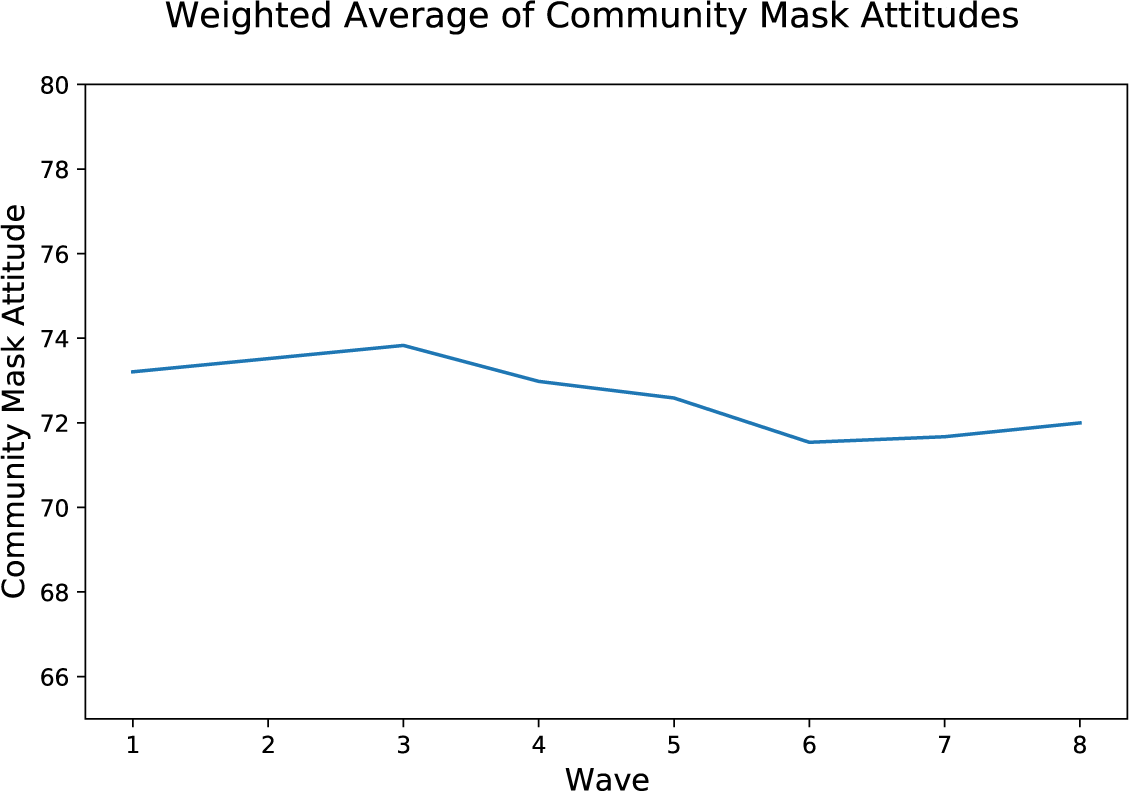
Average (weighted) community mask attitude response per wave.

**Figure S11:**
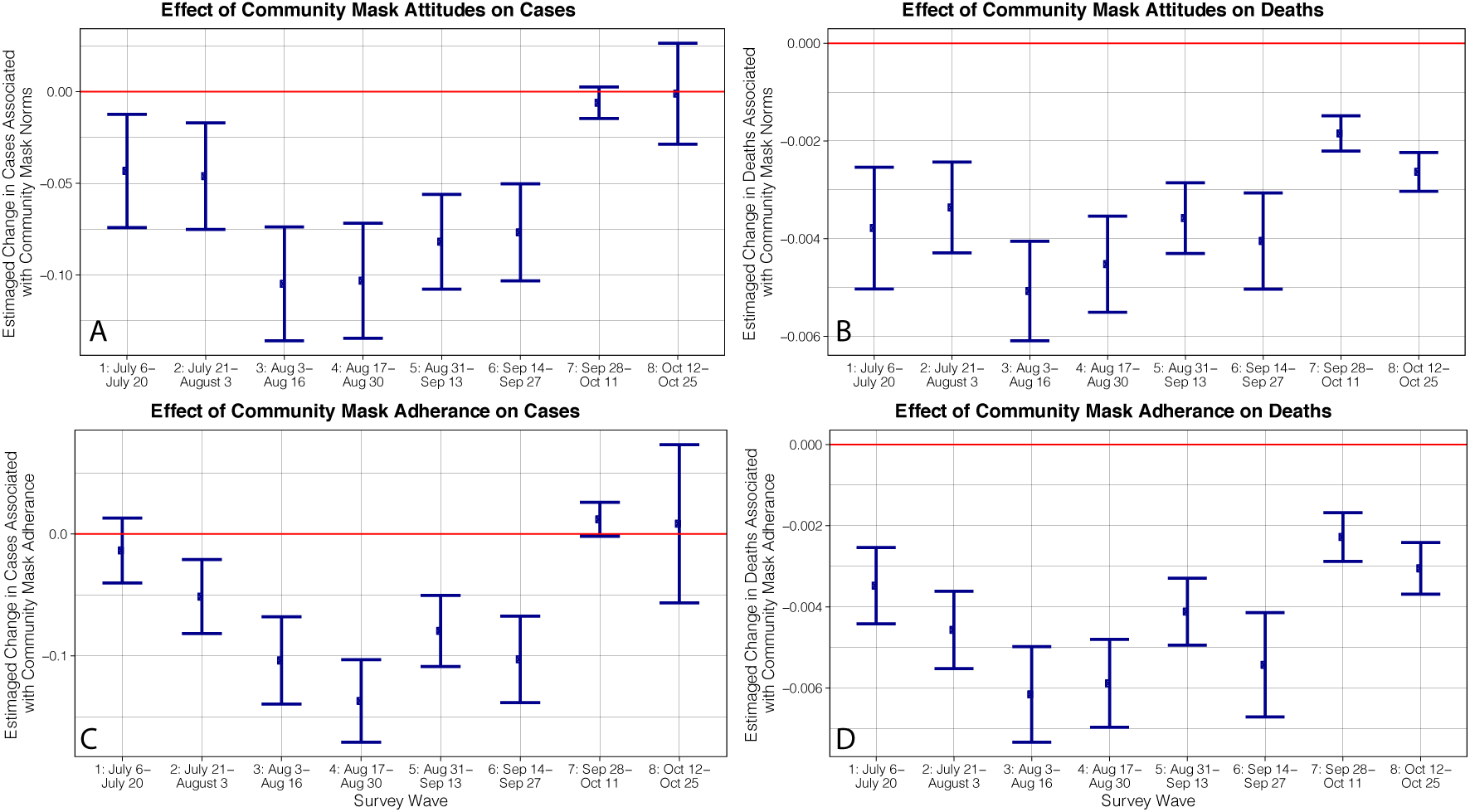
Dis-aggregated (by survey wave) estimates of the effect of community mask attitudes on COVID-19 cases **(A)** and deaths **(B)**, and community mask adherence on COVID-19 cases **(C)** and deaths **(D)** in a 68-country survey with 479K responses. Error bars show a 95% confidence interval. Full regression statistics are shown in tables S16, S17, S18 and S19.

### A.11 Event study and multi-linear regression tables

**Table S4:**
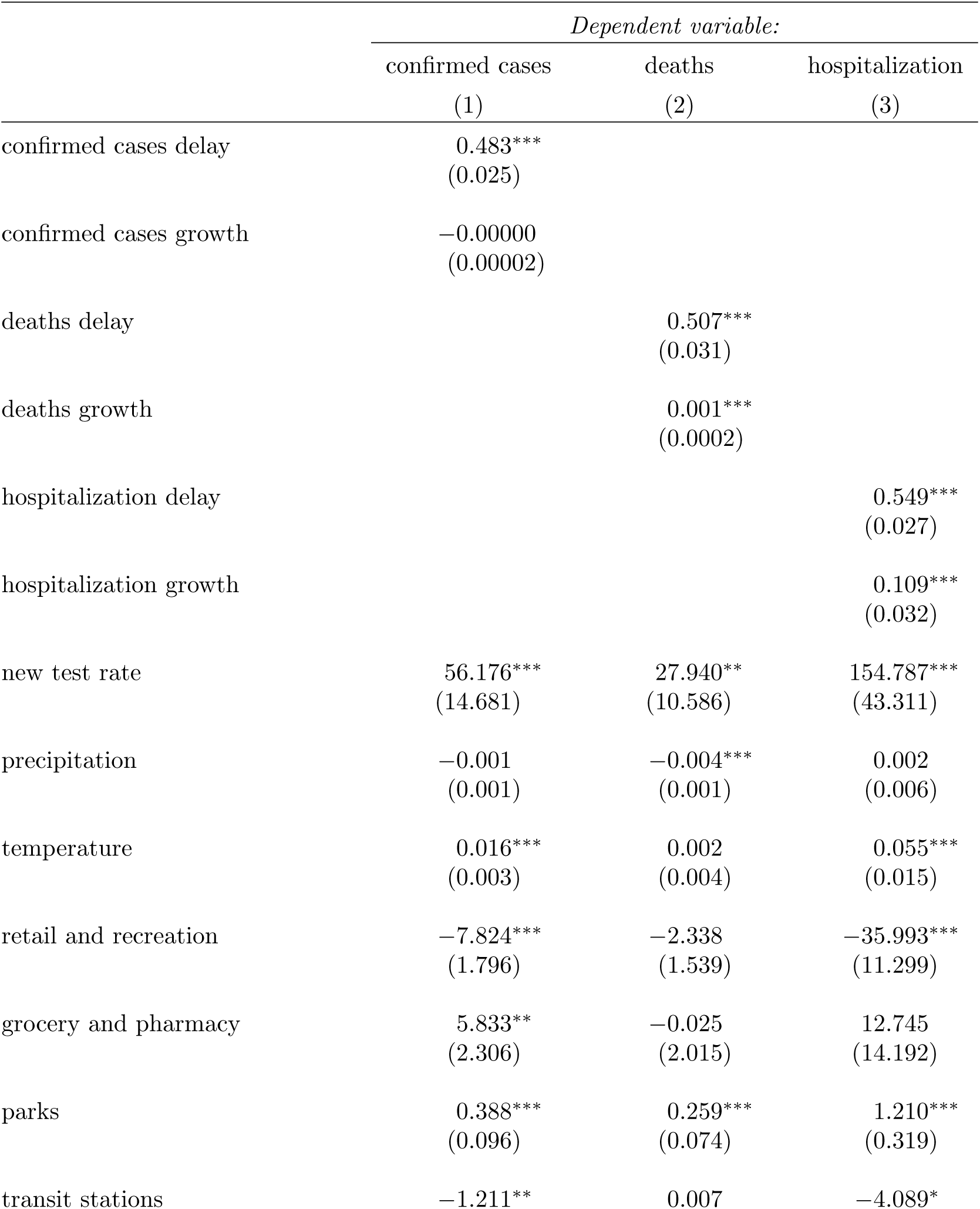
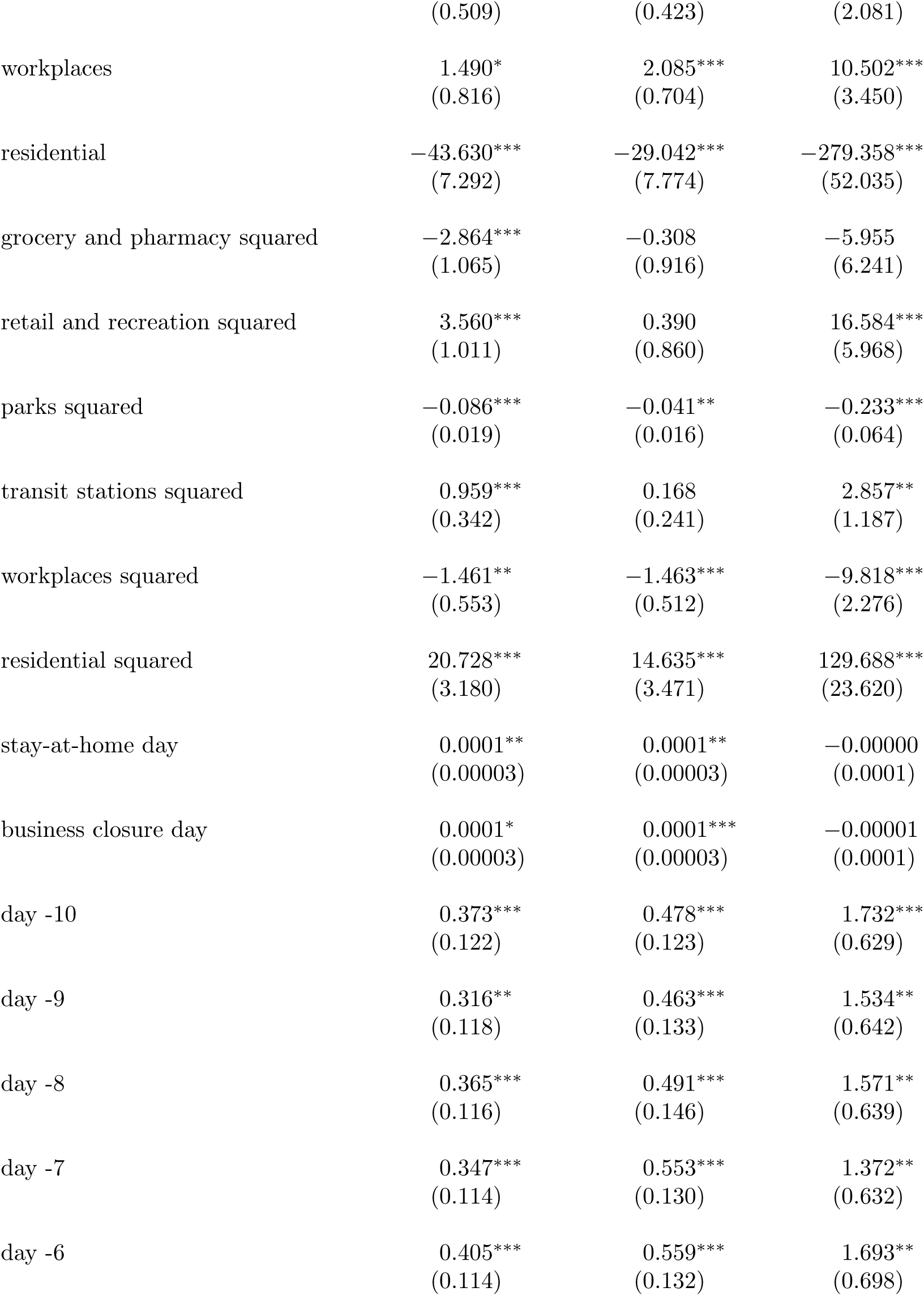
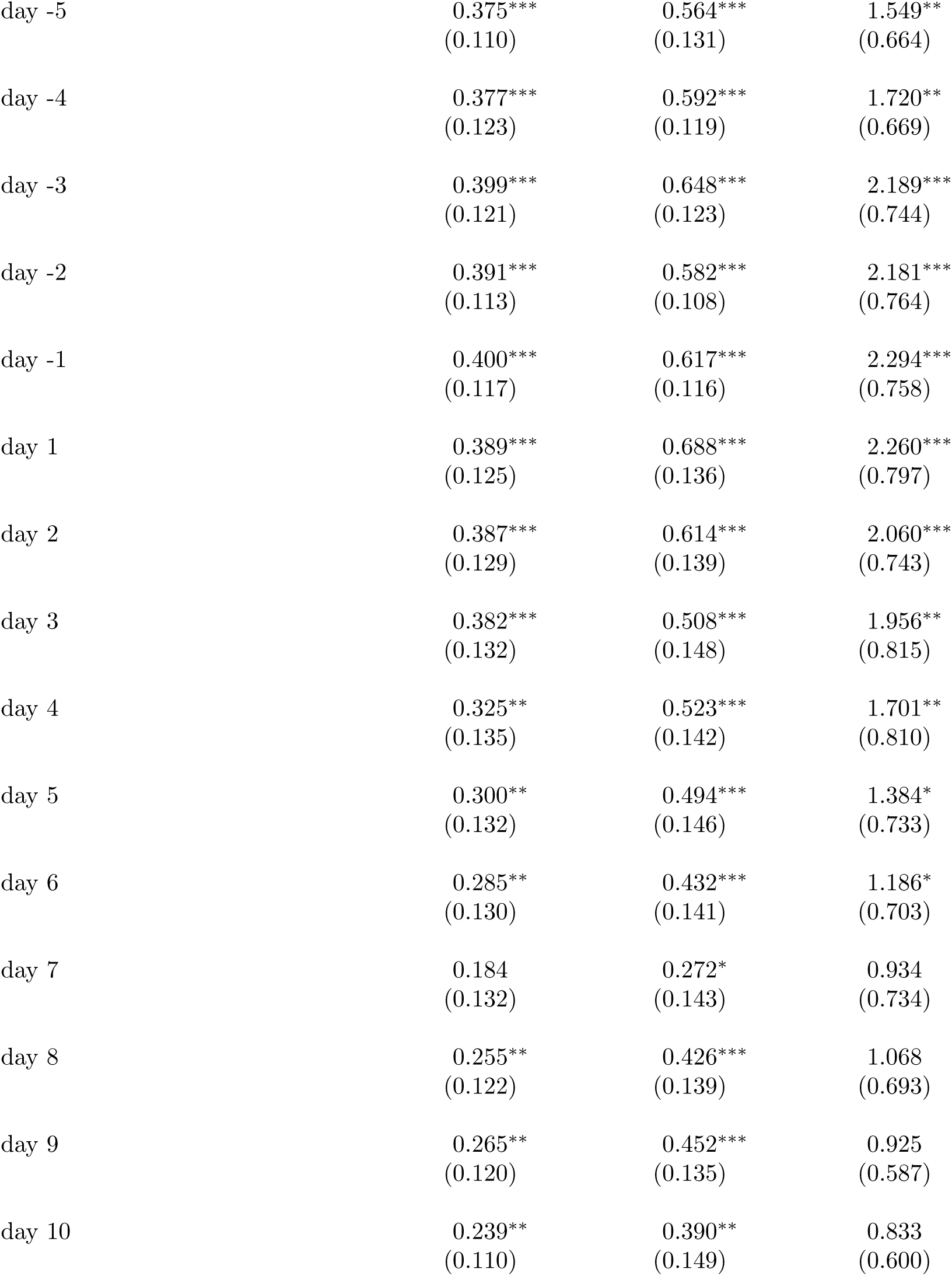
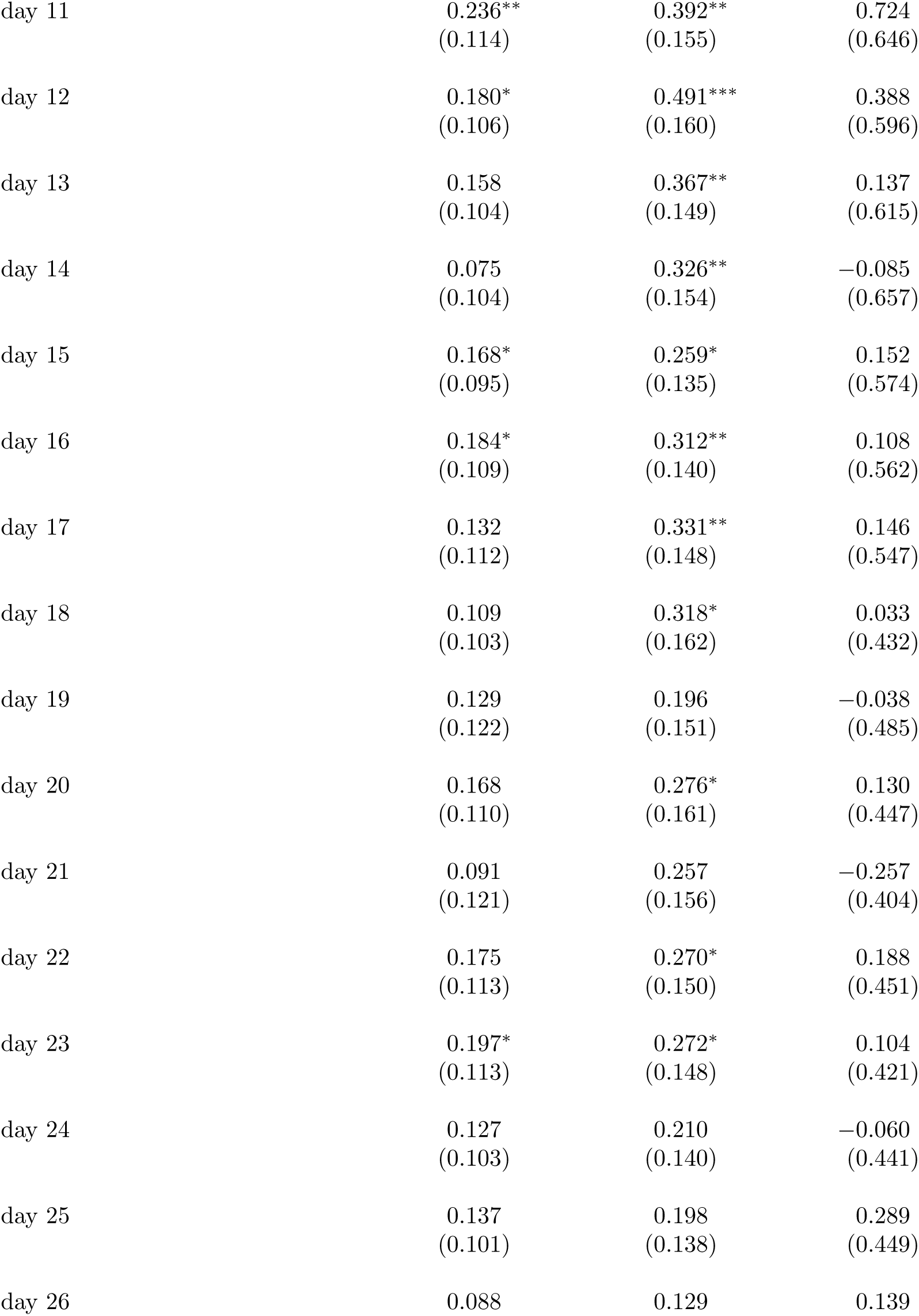
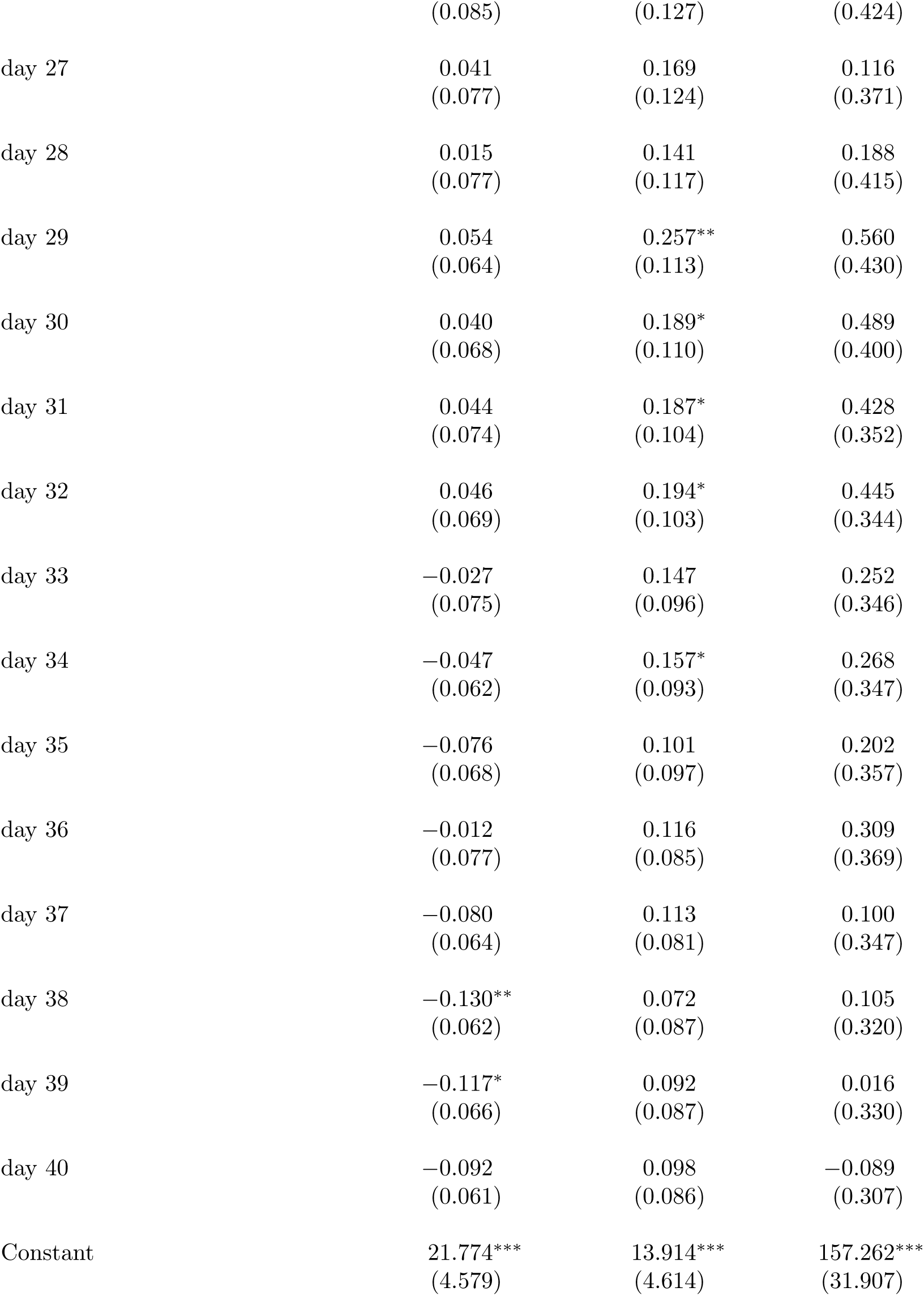
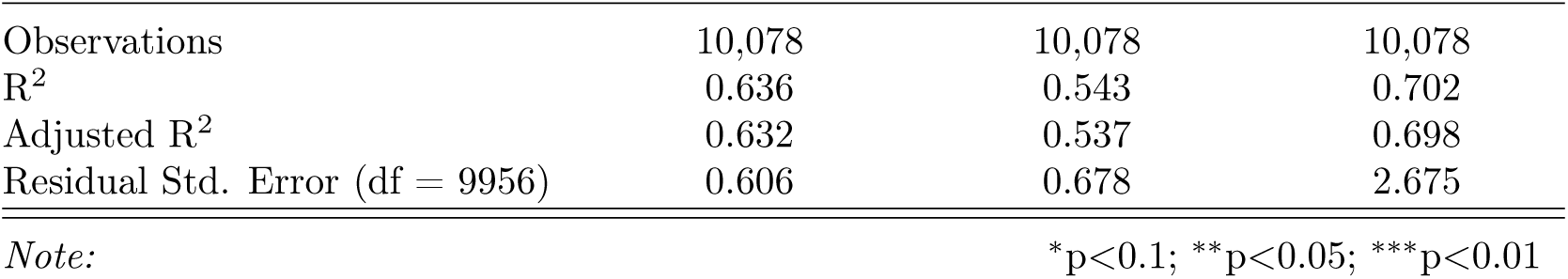
Event Study regression summary statistics for main result (section 2.1 fig. 1)

|  | *Dependent variable:* |  |  |
| --- | --- | --- | --- |
|  | confirmed cases | deaths | hospitalization |
|  | (1) | (2) | (3) |
| confirmed cases delay | 0.483***<br>(0.025) |  |  |
| confirmed cases growth | −0.00000<br>(0.00002) |  |  |
| deaths delay |  | 0.507***<br>(0.031) |  |
| deaths growth |  | 0.001***<br>(0.0002) |  |
| hospitalization delay |  |  | 0.549***<br>(0.027) |
| hospitalization growth |  |  | 0.109***<br>(0.032) |
| new test rate | 56.176***<br>(14.681) | 27.940**<br>(10.586) | 154.787***<br>(43.311) |
| precipitation | −0.001<br>(0.001) | −0.004***<br>(0.001) | 0.002<br>(0.006) |
| temperature | 0.016***<br>(0.003) | 0.002<br>(0.004) | 0.055***<br>(0.015) |
| retail and recreation | −7.824***<br>(1.796) | −2.338<br>(1.539) | −35.993***<br>(11.299) |
| grocery and pharmacy | 5.833**<br>(2.306) | −0.025<br>(2.015) | 12.745<br>(14.192) |
| parks | 0.388***<br>(0.096) | 0.259***<br>(0.074) | 1.210***<br>(0.319) |
| transit stations | −1.211** | 0.007 | −4.089* |
|  | (0.509) | (0.423) | (2.081) |
| workplaces | 1.490*<br>(0.816) | 2.085***<br>(0.704) | 10.502***<br>(3.450) |
| residential | -43.630***<br>(7.292) | -29.042***<br>(7.774) | -279.358***<br>(52.035) |
| grocery and pharmacy squared | -2.864***<br>(1.065) | -0.308<br>(0.916) | -5.955<br>(6.241) |
| retail and recreation squared | 3.560***<br>(1.011) | 0.390<br>(0.860) | 16.584***<br>(5.968) |
| parks squared | -0.086***<br>(0.019) | -0.041**<br>(0.016) | -0.233***<br>(0.064) |
| transit stations squared | 0.959***<br>(0.342) | 0.168<br>(0.241) | 2.857**<br>(1.187) |
| workplaces squared | -1.461**<br>(0.553) | -1.463***<br>(0.512) | -9.818***<br>(2.276) |
| residential squared | 20.728***<br>(3.180) | 14.635***<br>(3.471) | 129.688***<br>(23.620) |
| stay-at-home day | 0.0001**<br>(0.00003) | 0.0001**<br>(0.00003) | -0.00000<br>(0.0001) |
| business closure day | 0.0001*<br>(0.00003) | 0.0001***<br>(0.00003) | -0.00001<br>(0.0001) |
| day -10 | 0.373***<br>(0.122) | 0.478***<br>(0.123) | 1.732***<br>(0.629) |
| day -9 | 0.316**<br>(0.118) | 0.463***<br>(0.133) | 1.534**<br>(0.642) |
| day -8 | 0.365***<br>(0.116) | 0.491***<br>(0.146) | 1.571**<br>(0.639) |
| day -7 | 0.347***<br>(0.114) | 0.553***<br>(0.130) | 1.372**<br>(0.632) |
| day -6 | 0.405***<br>(0.114) | 0.559***<br>(0.132) | 1.693**<br>(0.698) |
| day -5 | 0.375***<br>(0.110) | 0.564***<br>(0.131) | 1.549**<br>(0.664) |
| day -4 | 0.377***<br>(0.123) | 0.592***<br>(0.119) | 1.720**<br>(0.669) |
| day -3 | 0.399***<br>(0.121) | 0.648***<br>(0.123) | 2.189***<br>(0.744) |
| day -2 | 0.391***<br>(0.113) | 0.582***<br>(0.108) | 2.181***<br>(0.764) |
| day -1 | 0.400***<br>(0.117) | 0.617***<br>(0.116) | 2.294***<br>(0.758) |
| day 1 | 0.389***<br>(0.125) | 0.688***<br>(0.136) | 2.260***<br>(0.797) |
| day 2 | 0.387***<br>(0.129) | 0.614***<br>(0.139) | 2.060***<br>(0.743) |
| day 3 | 0.382***<br>(0.132) | 0.508***<br>(0.148) | 1.956**<br>(0.815) |
| day 4 | 0.325**<br>(0.135) | 0.523***<br>(0.142) | 1.701**<br>(0.810) |
| day 5 | 0.300**<br>(0.132) | 0.494***<br>(0.146) | 1.384*<br>(0.733) |
| day 6 | 0.285**<br>(0.130) | 0.432***<br>(0.141) | 1.186*<br>(0.703) |
| day 7 | 0.184<br>(0.132) | 0.272*<br>(0.143) | 0.934<br>(0.734) |
| day 8 | 0.255**<br>(0.122) | 0.426***<br>(0.139) | 1.068<br>(0.693) |
| day 9 | 0.265**<br>(0.120) | 0.452***<br>(0.135) | 0.925<br>(0.587) |
| day 10 | 0.239**<br>(0.110) | 0.390**<br>(0.149) | 0.833<br>(0.600) |
| day 11 | 0.236**<br>(0.114) | 0.392**<br>(0.155) | 0.724<br>(0.646) |
| day 12 | 0.180*<br>(0.106) | 0.491***<br>(0.160) | 0.388<br>(0.596) |
| day 13 | 0.158<br>(0.104) | 0.367**<br>(0.149) | 0.137<br>(0.615) |
| day 14 | 0.075<br>(0.104) | 0.326**<br>(0.154) | −0.085<br>(0.657) |
| day 15 | 0.168*<br>(0.095) | 0.259*<br>(0.135) | 0.152<br>(0.574) |
| day 16 | 0.184*<br>(0.109) | 0.312**<br>(0.140) | 0.108<br>(0.562) |
| day 17 | 0.132<br>(0.112) | 0.331**<br>(0.148) | 0.146<br>(0.547) |
| day 18 | 0.109<br>(0.103) | 0.318*<br>(0.162) | 0.033<br>(0.432) |
| day 19 | 0.129<br>(0.122) | 0.196<br>(0.151) | −0.038<br>(0.485) |
| day 20 | 0.168<br>(0.110) | 0.276*<br>(0.161) | 0.130<br>(0.447) |
| day 21 | 0.091<br>(0.121) | 0.257<br>(0.156) | −0.257<br>(0.404) |
| day 22 | 0.175<br>(0.113) | 0.270*<br>(0.150) | 0.188<br>(0.451) |
| day 23 | 0.197*<br>(0.113) | 0.272*<br>(0.148) | 0.104<br>(0.421) |
| day 24 | 0.127<br>(0.103) | 0.210<br>(0.140) | −0.060<br>(0.441) |
| day 25 | 0.137<br>(0.101) | 0.198<br>(0.138) | 0.289<br>(0.449) |
| day 26 | 0.088 | 0.129 | 0.139 |
|  | (0.085) | (0.127) | (0.424) |
| day 27 | 0.041<br>(0.077) | 0.169<br>(0.124) | 0.116<br>(0.371) |
| day 28 | 0.015<br>(0.077) | 0.141<br>(0.117) | 0.188<br>(0.415) |
| day 29 | 0.054<br>(0.064) | 0.257**<br>(0.113) | 0.560<br>(0.430) |
| day 30 | 0.040<br>(0.068) | 0.189*<br>(0.110) | 0.489<br>(0.400) |
| day 31 | 0.044<br>(0.074) | 0.187*<br>(0.104) | 0.428<br>(0.352) |
| day 32 | 0.046<br>(0.069) | 0.194*<br>(0.103) | 0.445<br>(0.344) |
| day 33 | −0.027<br>(0.075) | 0.147<br>(0.096) | 0.252<br>(0.346) |
| day 34 | −0.047<br>(0.062) | 0.157*<br>(0.093) | 0.268<br>(0.347) |
| day 35 | −0.076<br>(0.068) | 0.101<br>(0.097) | 0.202<br>(0.357) |
| day 36 | −0.012<br>(0.077) | 0.116<br>(0.085) | 0.309<br>(0.369) |
| day 37 | −0.080<br>(0.064) | 0.113<br>(0.081) | 0.100<br>(0.347) |
| day 38 | −0.130**<br>(0.062) | 0.072<br>(0.087) | 0.105<br>(0.320) |
| day 39 | −0.117*<br>(0.066) | 0.092<br>(0.087) | 0.016<br>(0.330) |
| day 40 | −0.092<br>(0.061) | 0.098<br>(0.086) | −0.089<br>(0.307) |
| Constant | 21.774***<br>(4.579) | 13.914***<br>(4.614) | 157.262***<br>(31.907) |
| Observations | 10,078 | 10,078 | 10,078 |
| R<sup>2</sup> | 0.636 | 0.543 | 0.702 |
| Adjusted R<sup>2</sup> | 0.632 | 0.537 | 0.698 |
| Residual Std. Error (df = 9956) | 0.606 | 0.678 | 2.675 |

*Note:*
\*p<0.1; \*\*p<0.05; \*\*\*p<0.01

**Table S5:**
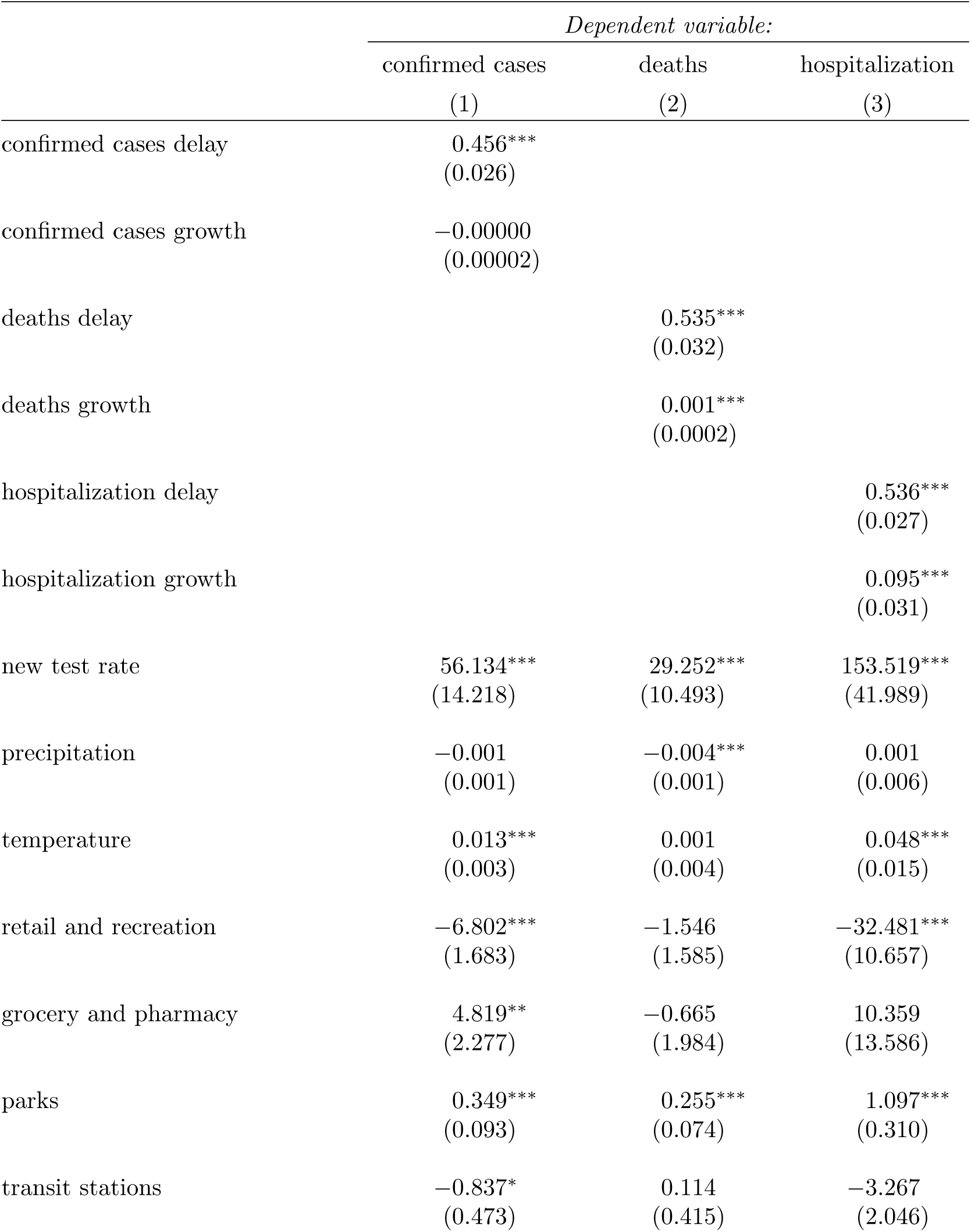
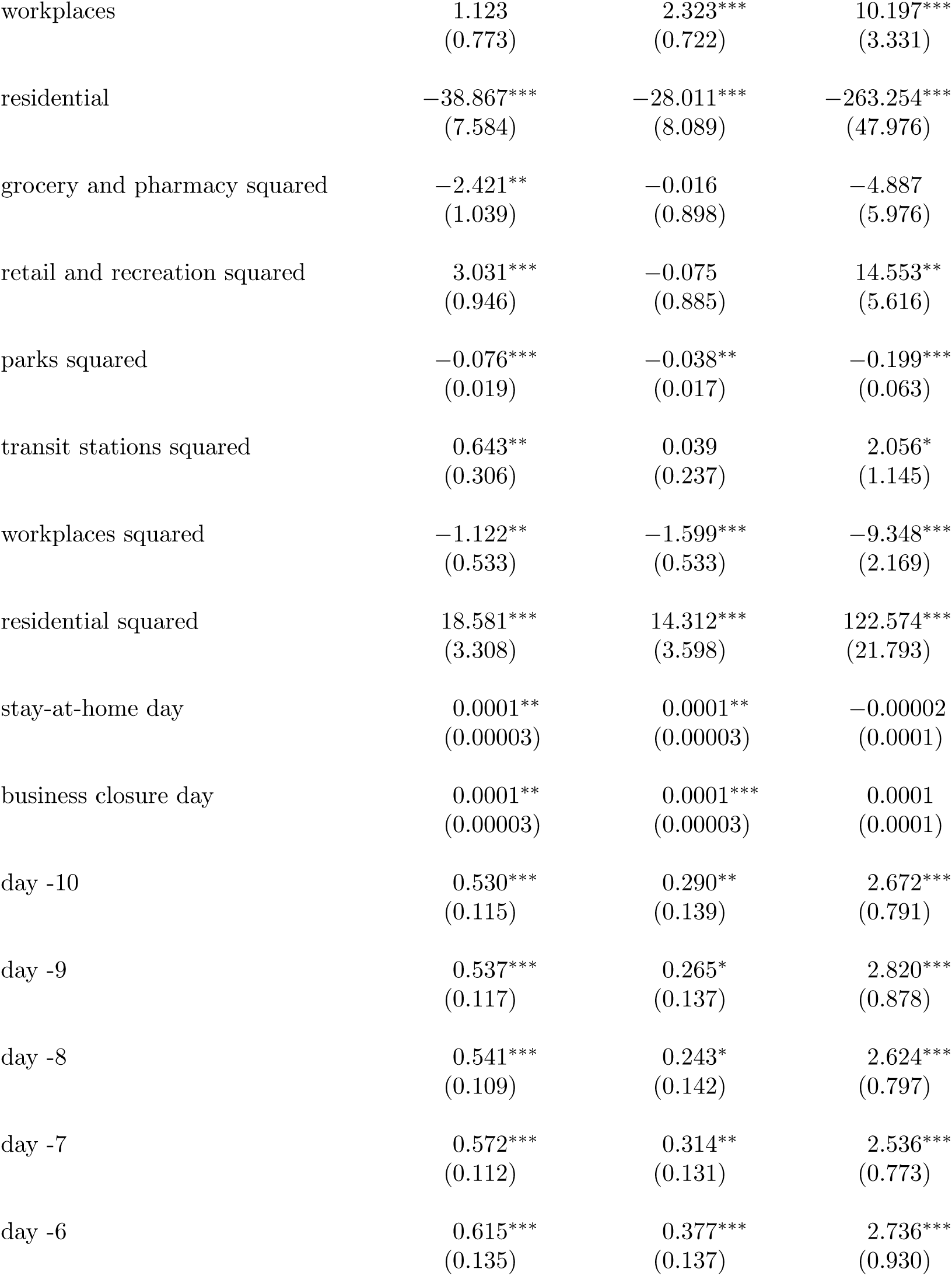
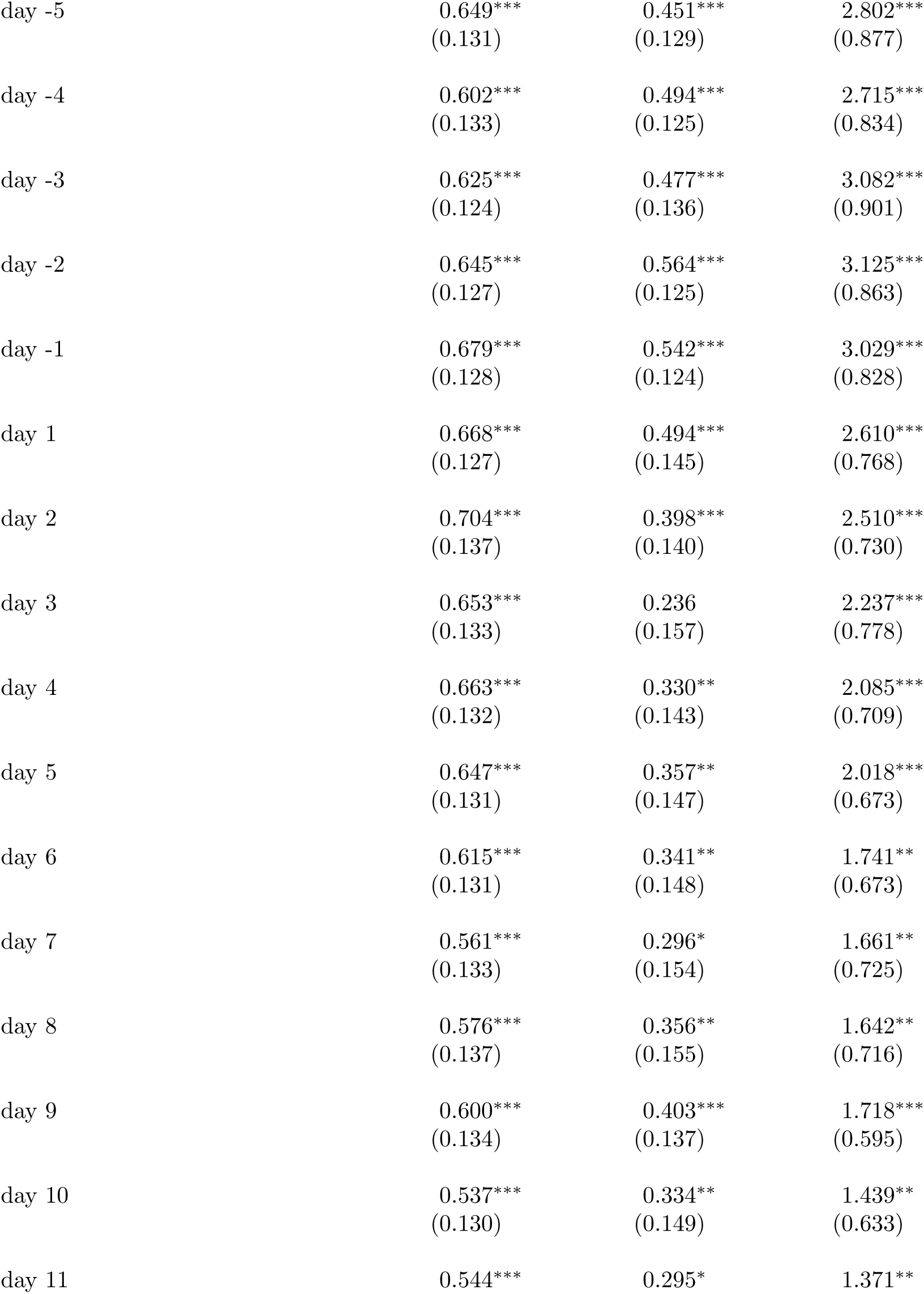
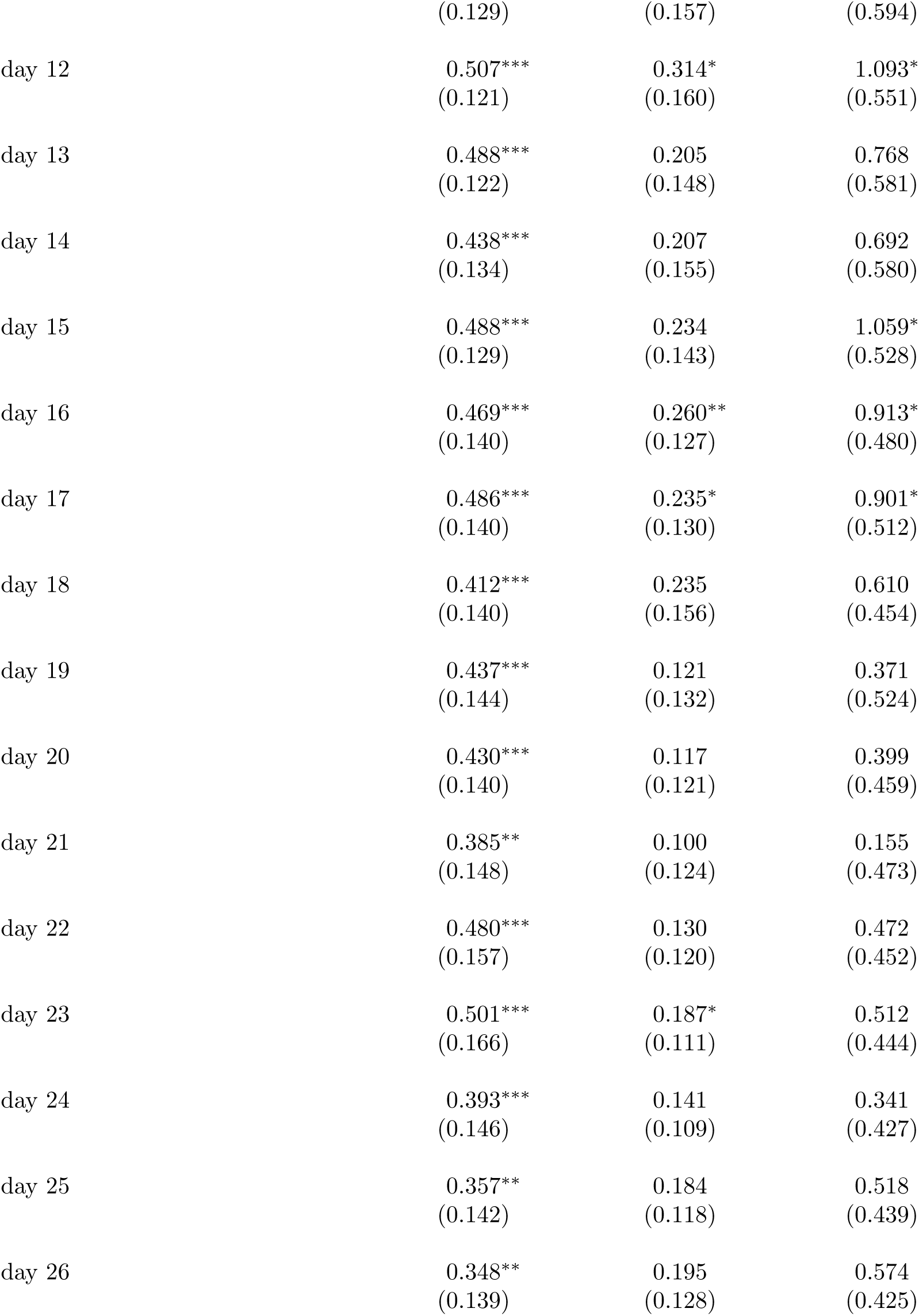
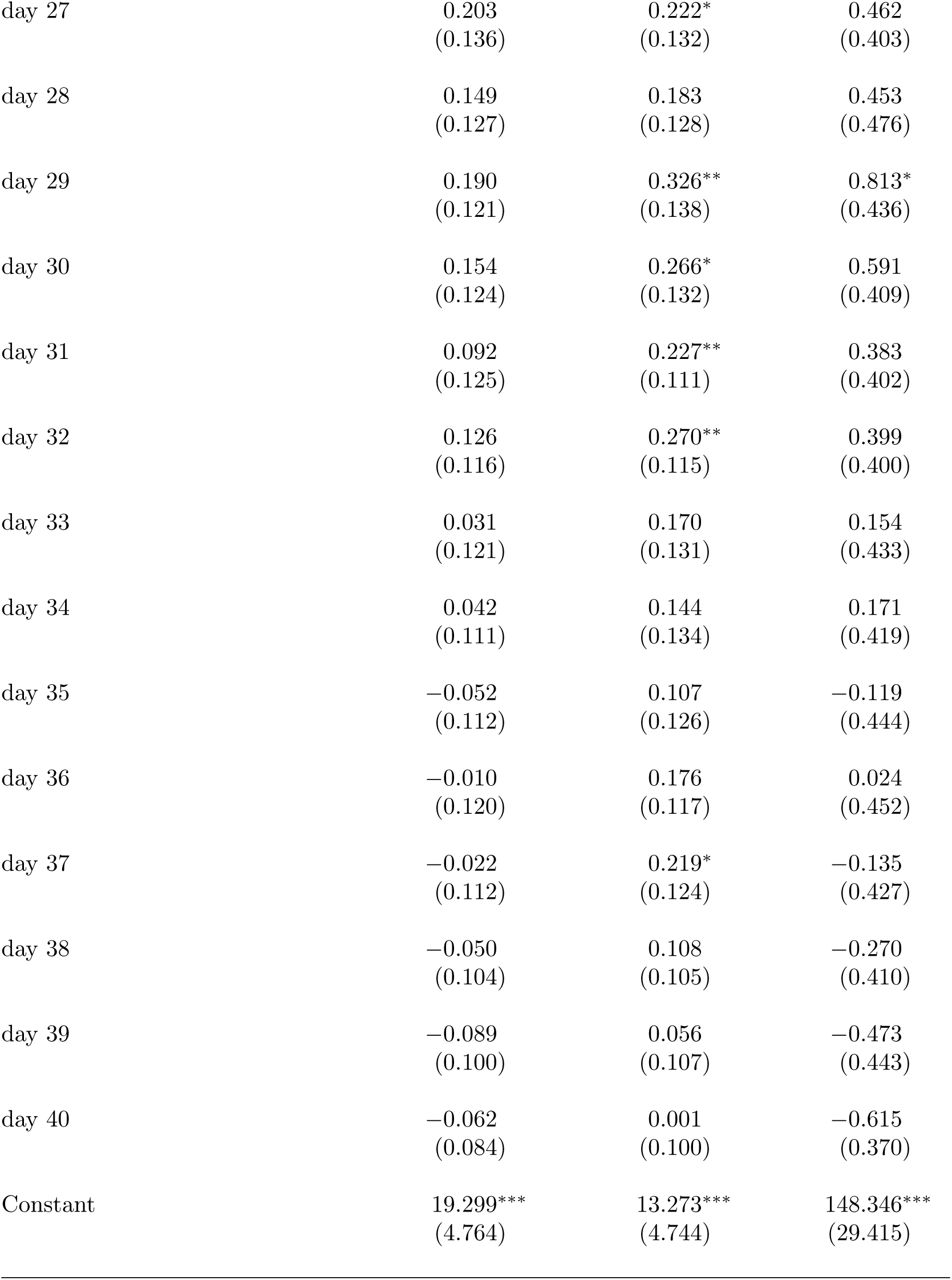
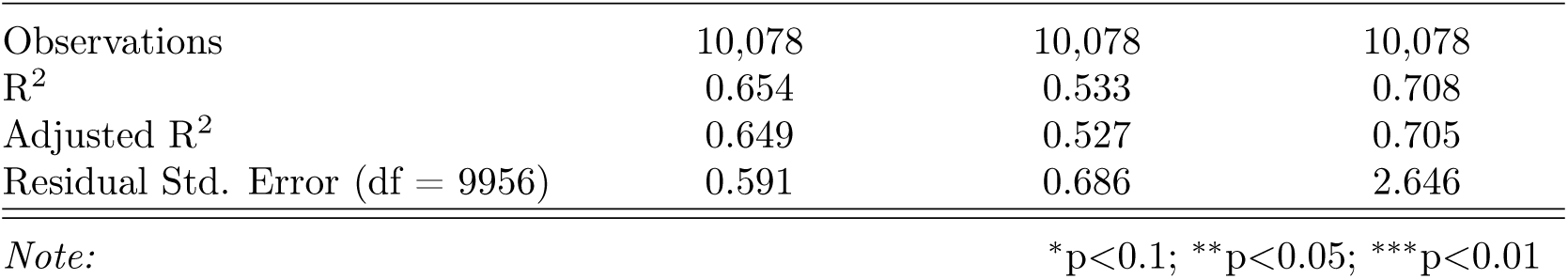
Event Study regression summary statistics for robustness check (fig. 2) where we only consider mask mandates that require the public to wear masks.

|  | *Dependent variable:* |  |  |
| --- | --- | --- | --- |
|  | confirmed cases | deaths | hospitalization |
|  | (1) | (2) | (3) |
| confirmed cases delay | 0.456***<br>(0.026) |  |  |
| confirmed cases growth | -0.00000<br>(0.00002) |  |  |
| deaths delay |  | 0.535***<br>(0.032) |  |
| deaths growth |  | 0.001***<br>(0.0002) |  |
| hospitalization delay |  |  | 0.536***<br>(0.027) |
| hospitalization growth |  |  | 0.095***<br>(0.031) |
| new test rate | 56.134***<br>(14.218) | 29.252***<br>(10.493) | 153.519***<br>(41.989) |
| precipitation | -0.001<br>(0.001) | -0.004***<br>(0.001) | 0.001<br>(0.006) |
| temperature | 0.013***<br>(0.003) | 0.001<br>(0.004) | 0.048***<br>(0.015) |
| retail and recreation | -6.802***<br>(1.683) | -1.546<br>(1.585) | -32.481***<br>(10.657) |
| grocery and pharmacy | 4.819**<br>(2.277) | -0.665<br>(1.984) | 10.359<br>(13.586) |
| parks | 0.349***<br>(0.093) | 0.255***<br>(0.074) | 1.097***<br>(0.310) |
| transit stations | -0.837*<br>(0.473) | 0.114<br>(0.415) | -3.267<br>(2.046) |
| workplaces | 1.123<br>(0.773) | 2.323***<br>(0.722) | 10.197***<br>(3.331) |
| residential | -38.867***<br>(7.584) | -28.011***<br>(8.089) | -263.254***<br>(47.976) |
| grocery and pharmacy squared | -2.421**<br>(1.039) | -0.016<br>(0.898) | -4.887<br>(5.976) |
| retail and recreation squared | 3.031***<br>(0.946) | -0.075<br>(0.885) | 14.553**<br>(5.616) |
| parks squared | -0.076***<br>(0.019) | -0.038**<br>(0.017) | -0.199***<br>(0.063) |
| transit stations squared | 0.643**<br>(0.306) | 0.039<br>(0.237) | 2.056*<br>(1.145) |
| workplaces squared | -1.122**<br>(0.533) | -1.599***<br>(0.533) | -9.348***<br>(2.169) |
| residential squared | 18.581***<br>(3.308) | 14.312***<br>(3.598) | 122.574***<br>(21.793) |
| stay-at-home day | 0.0001**<br>(0.00003) | 0.0001**<br>(0.00003) | -0.00002<br>(0.0001) |
| business closure day | 0.0001**<br>(0.00003) | 0.0001***<br>(0.00003) | 0.0001<br>(0.0001) |
| day -10 | 0.530***<br>(0.115) | 0.290**<br>(0.139) | 2.672***<br>(0.791) |
| day -9 | 0.537***<br>(0.117) | 0.265*<br>(0.137) | 2.820***<br>(0.878) |
| day -8 | 0.541***<br>(0.109) | 0.243*<br>(0.142) | 2.624***<br>(0.797) |
| day -7 | 0.572***<br>(0.112) | 0.314**<br>(0.131) | 2.536***<br>(0.773) |
| day -6 | 0.615***<br>(0.135) | 0.377***<br>(0.137) | 2.736***<br>(0.930) |
| day -5 | 0.649***<br>(0.131) | 0.451***<br>(0.129) | 2.802***<br>(0.877) |
| day -4 | 0.602***<br>(0.133) | 0.494***<br>(0.125) | 2.715***<br>(0.834) |
| day -3 | 0.625***<br>(0.124) | 0.477***<br>(0.136) | 3.082***<br>(0.901) |
| day -2 | 0.645***<br>(0.127) | 0.564***<br>(0.125) | 3.125***<br>(0.863) |
| day -1 | 0.679***<br>(0.128) | 0.542***<br>(0.124) | 3.029***<br>(0.828) |
| day 1 | 0.668***<br>(0.127) | 0.494***<br>(0.145) | 2.610***<br>(0.768) |
| day 2 | 0.704***<br>(0.137) | 0.398***<br>(0.140) | 2.510***<br>(0.730) |
| day 3 | 0.653***<br>(0.133) | 0.236<br>(0.157) | 2.237***<br>(0.778) |
| day 4 | 0.663***<br>(0.132) | 0.330**<br>(0.143) | 2.085***<br>(0.709) |
| day 5 | 0.647***<br>(0.131) | 0.357**<br>(0.147) | 2.018***<br>(0.673) |
| day 6 | 0.615***<br>(0.131) | 0.341**<br>(0.148) | 1.741**<br>(0.673) |
| day 7 | 0.561***<br>(0.133) | 0.296*<br>(0.154) | 1.661**<br>(0.725) |
| day 8 | 0.576***<br>(0.137) | 0.356**<br>(0.155) | 1.642**<br>(0.716) |
| day 9 | 0.600***<br>(0.134) | 0.403***<br>(0.137) | 1.718***<br>(0.595) |
| day 10 | 0.537***<br>(0.130) | 0.334**<br>(0.149) | 1.439**<br>(0.633) |
| day 11 | 0.544*** | 0.295* | 1.371** |
|  | (0.129) | (0.157) | (0.594) |
| day 12 | 0.507***<br>(0.121) | 0.314*<br>(0.160) | 1.093*<br>(0.551) |
| day 13 | 0.488***<br>(0.122) | 0.205<br>(0.148) | 0.768<br>(0.581) |
| day 14 | 0.438***<br>(0.134) | 0.207<br>(0.155) | 0.692<br>(0.580) |
| day 15 | 0.488***<br>(0.129) | 0.234<br>(0.143) | 1.059*<br>(0.528) |
| day 16 | 0.469***<br>(0.140) | 0.260**<br>(0.127) | 0.913*<br>(0.480) |
| day 17 | 0.486***<br>(0.140) | 0.235*<br>(0.130) | 0.901*<br>(0.512) |
| day 18 | 0.412***<br>(0.140) | 0.235<br>(0.156) | 0.610<br>(0.454) |
| day 19 | 0.437***<br>(0.144) | 0.121<br>(0.132) | 0.371<br>(0.524) |
| day 20 | 0.430***<br>(0.140) | 0.117<br>(0.121) | 0.399<br>(0.459) |
| day 21 | 0.385**<br>(0.148) | 0.100<br>(0.124) | 0.155<br>(0.473) |
| day 22 | 0.480***<br>(0.157) | 0.130<br>(0.120) | 0.472<br>(0.452) |
| day 23 | 0.501***<br>(0.166) | 0.187*<br>(0.111) | 0.512<br>(0.444) |
| day 24 | 0.393***<br>(0.146) | 0.141<br>(0.109) | 0.341<br>(0.427) |
| day 25 | 0.357**<br>(0.142) | 0.184<br>(0.118) | 0.518<br>(0.439) |
| day 26 | 0.348**<br>(0.139) | 0.195<br>(0.128) | 0.574<br>(0.425) |
| day 27 | 0.203<br>(0.136) | 0.222*<br>(0.132) | 0.462<br>(0.403) |
| day 28 | 0.149<br>(0.127) | 0.183<br>(0.128) | 0.453<br>(0.476) |
| day 29 | 0.190<br>(0.121) | 0.326**<br>(0.138) | 0.813*<br>(0.436) |
| day 30 | 0.154<br>(0.124) | 0.266*<br>(0.132) | 0.591<br>(0.409) |
| day 31 | 0.092<br>(0.125) | 0.227**<br>(0.111) | 0.383<br>(0.402) |
| day 32 | 0.126<br>(0.116) | 0.270**<br>(0.115) | 0.399<br>(0.400) |
| day 33 | 0.031<br>(0.121) | 0.170<br>(0.131) | 0.154<br>(0.433) |
| day 34 | 0.042<br>(0.111) | 0.144<br>(0.134) | 0.171<br>(0.419) |
| day 35 | −0.052<br>(0.112) | 0.107<br>(0.126) | −0.119<br>(0.444) |
| day 36 | −0.010<br>(0.120) | 0.176<br>(0.117) | 0.024<br>(0.452) |
| day 37 | −0.022<br>(0.112) | 0.219*<br>(0.124) | −0.135<br>(0.427) |
| day 38 | −0.050<br>(0.104) | 0.108<br>(0.105) | −0.270<br>(0.410) |
| day 39 | −0.089<br>(0.100) | 0.056<br>(0.107) | −0.473<br>(0.443) |
| day 40 | −0.062<br>(0.084) | 0.001<br>(0.100) | −0.615<br>(0.370) |
| Constant | 19.299***<br>(4.764) | 13.273***<br>(4.744) | 148.346***<br>(29.415) |
| Observations | 10,078 | 10,078 | 10,078 |
| R<sup>2</sup> | 0.654 | 0.533 | 0.708 |
| Adjusted R<sup>2</sup> | 0.649 | 0.527 | 0.705 |
| Residual Std. Error (df = 9956) | 0.591 | 0.686 | 2.646 |
| *Note:* |  | *p<0.1; **p<0.05; ***p<0.01 |  |

**Table S6:**
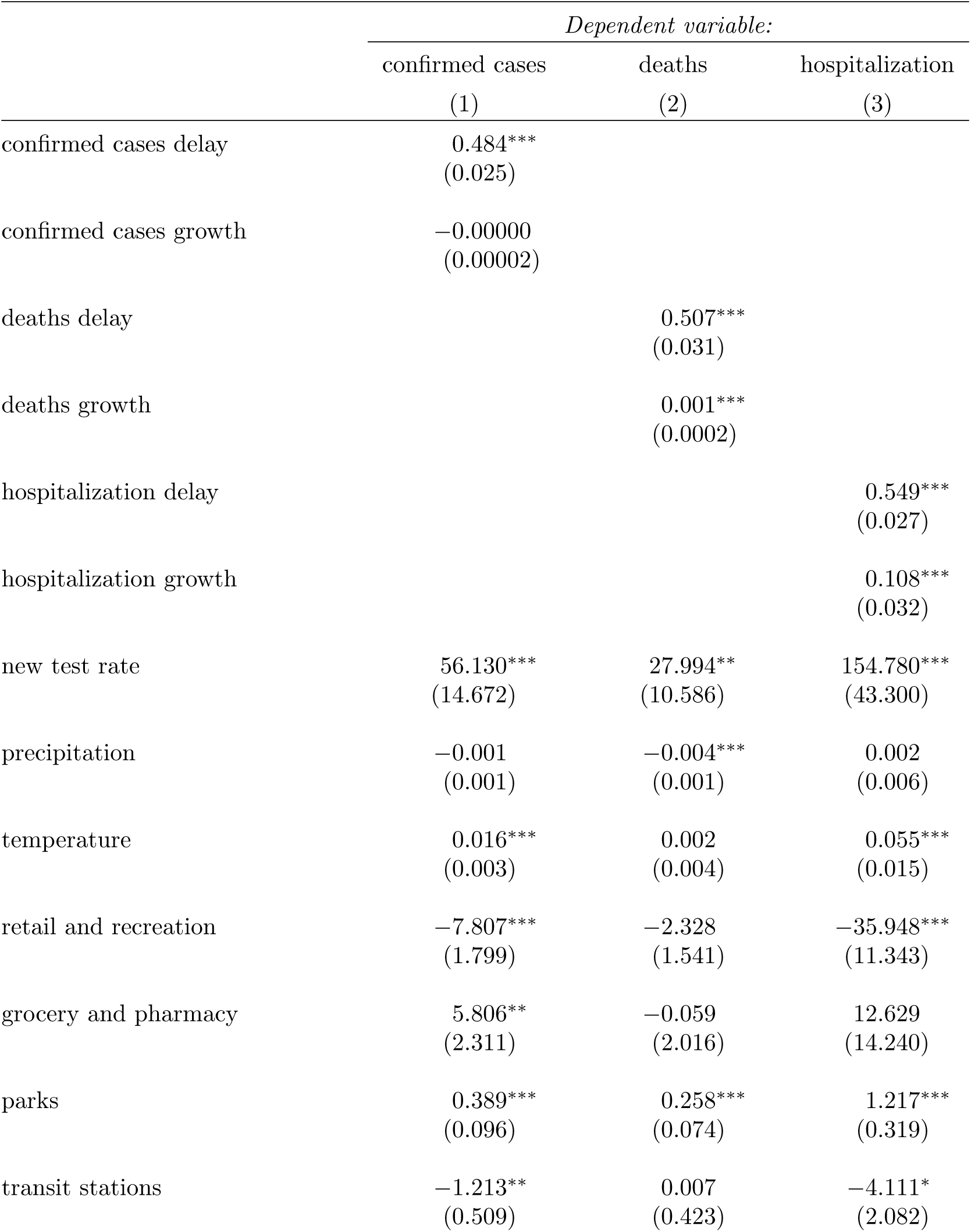
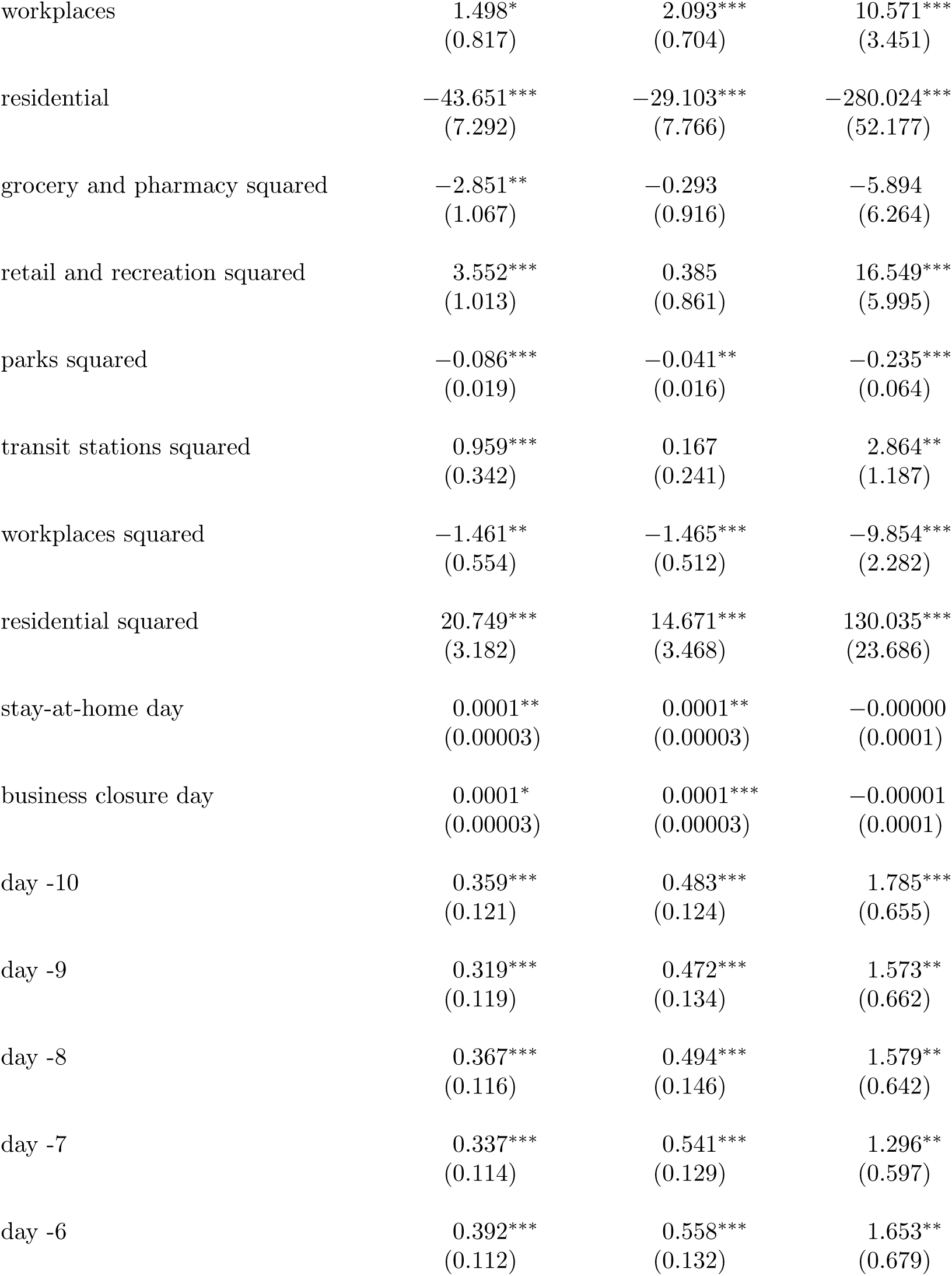
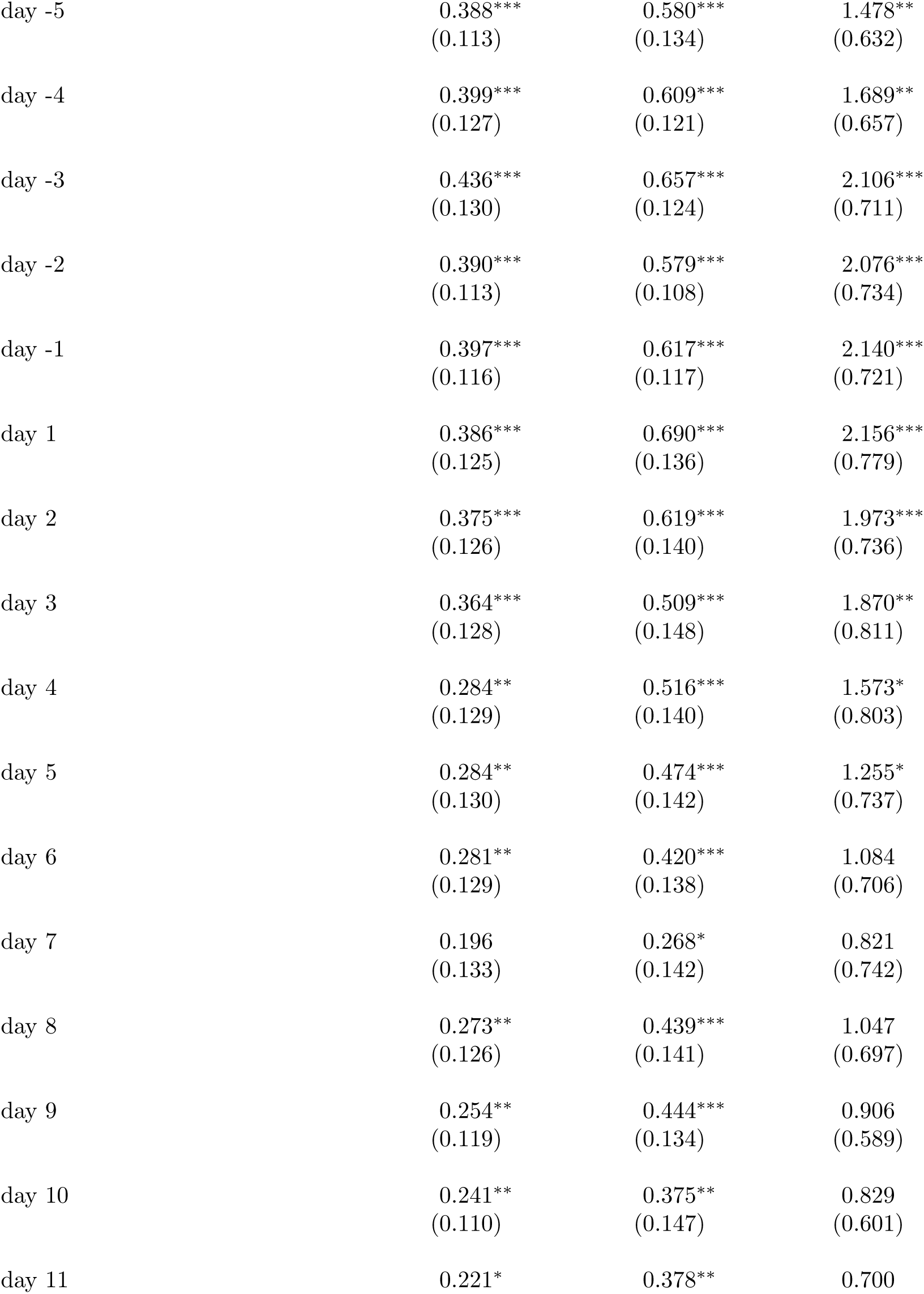
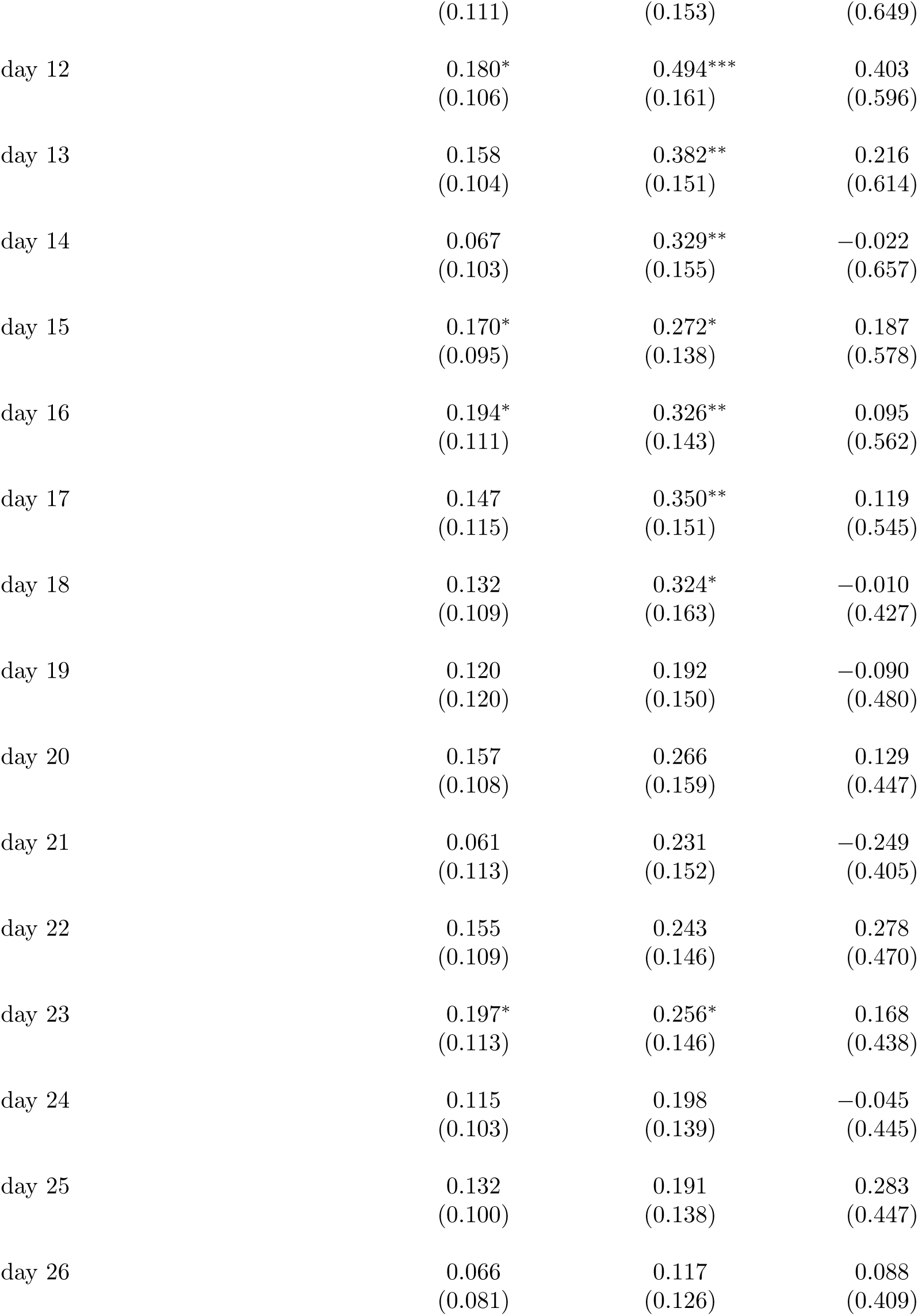
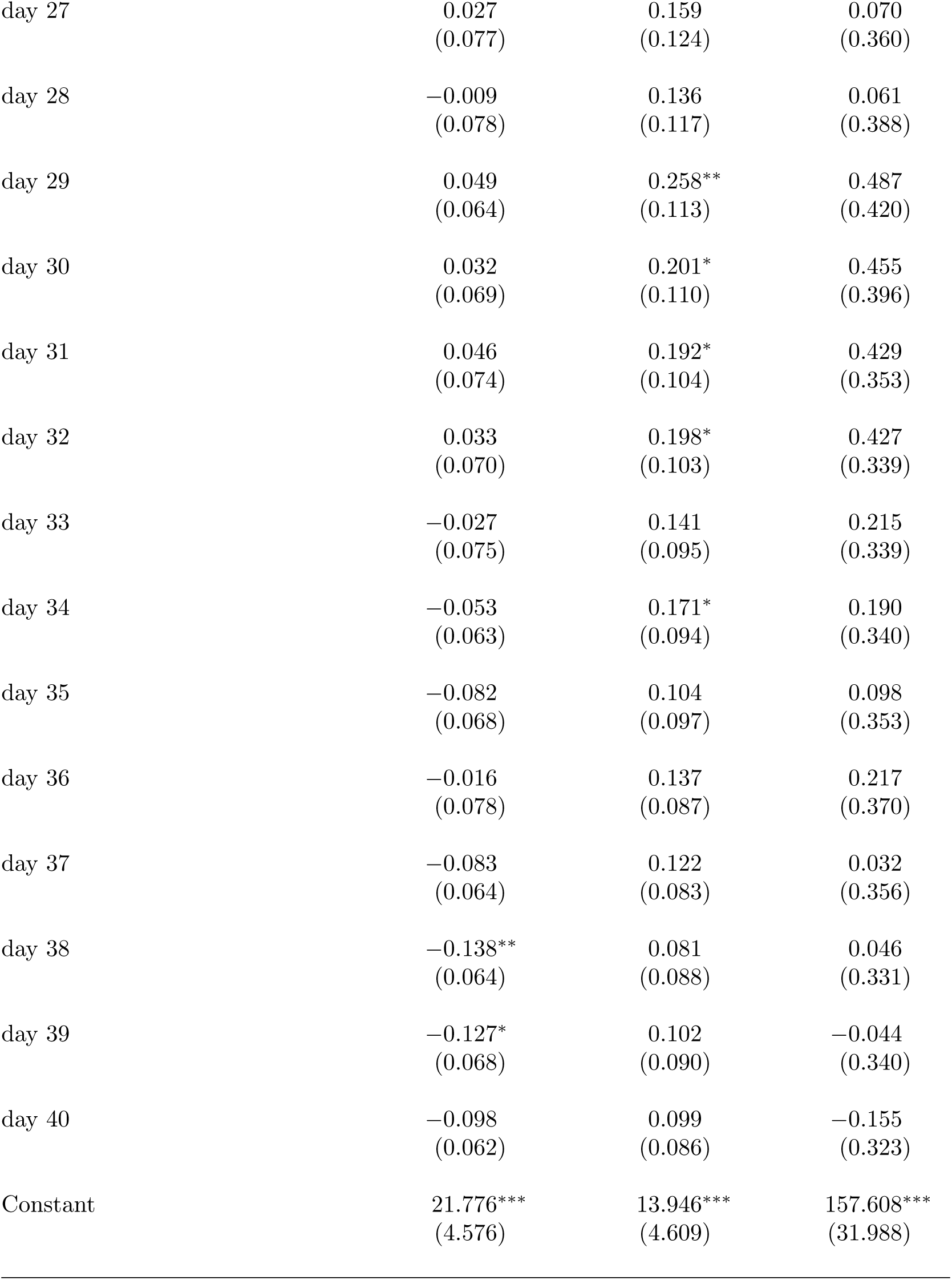
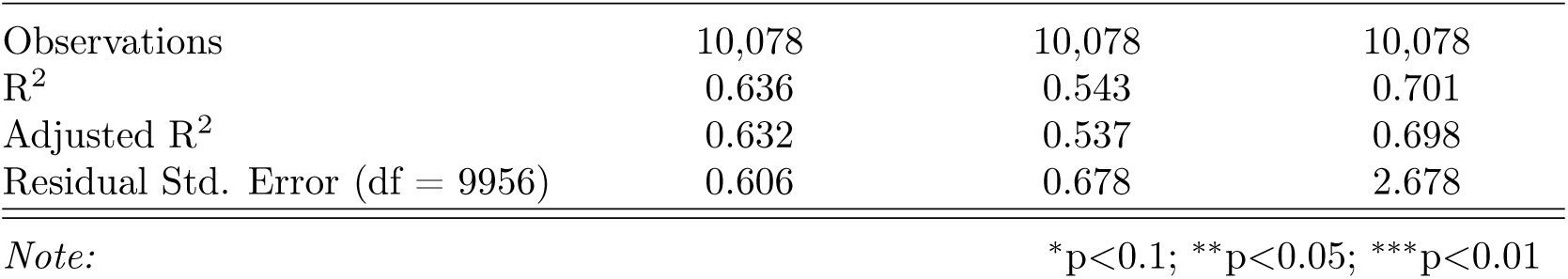
Event Study regression summary statistics for robustness check (fig. 2) where we only consider mask mandates that require business employees to wear masks.

**Table S7:**
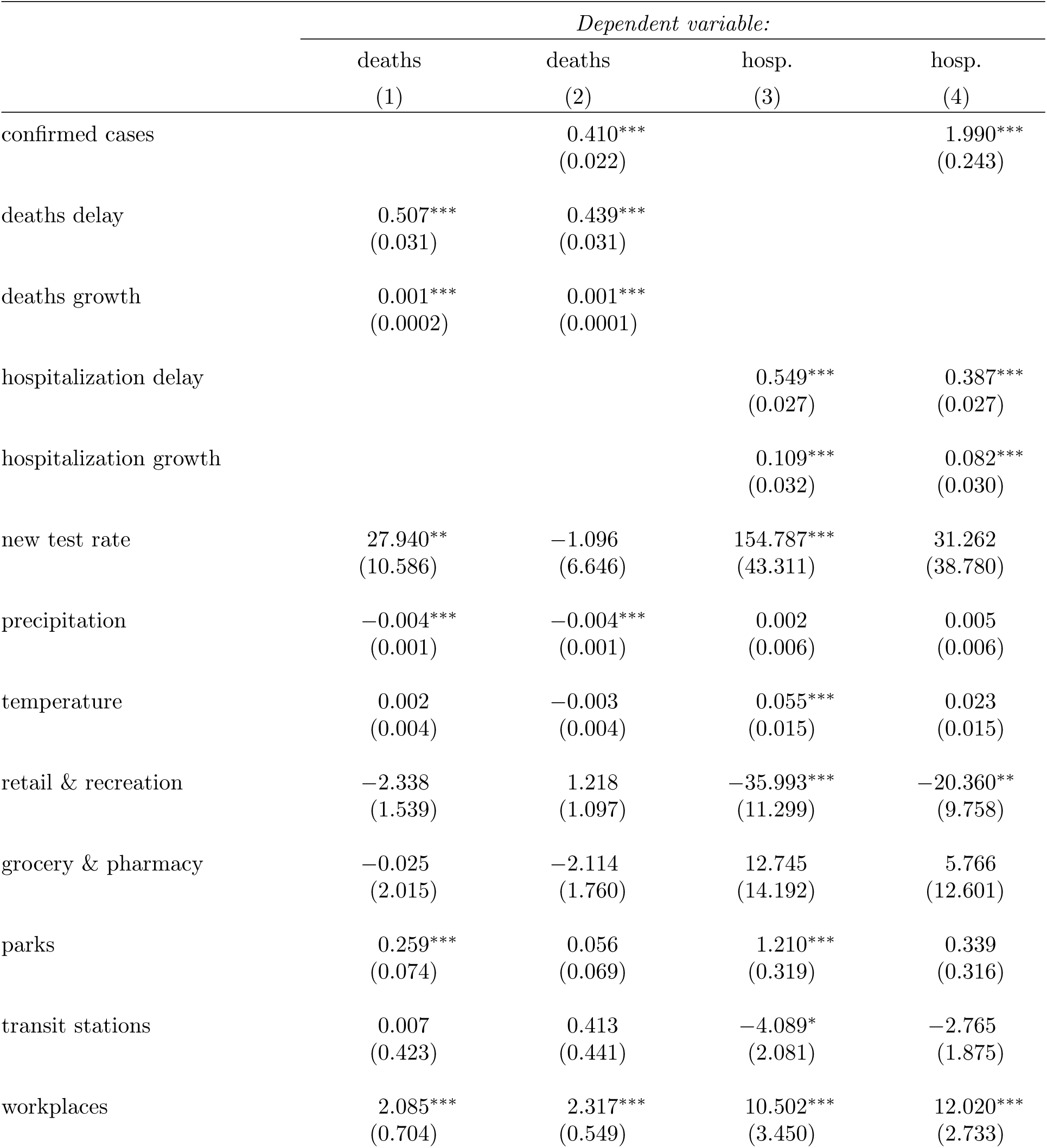
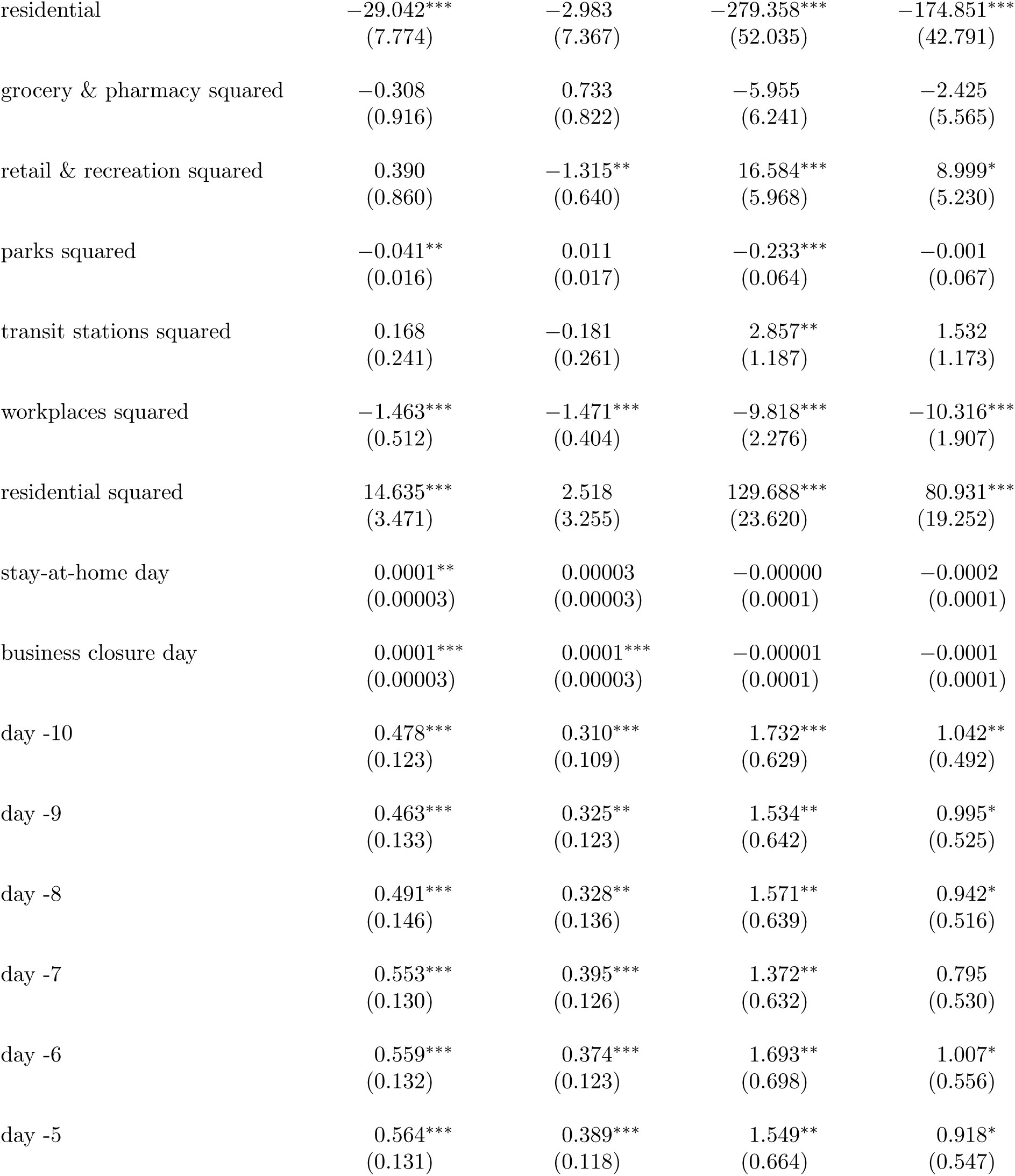
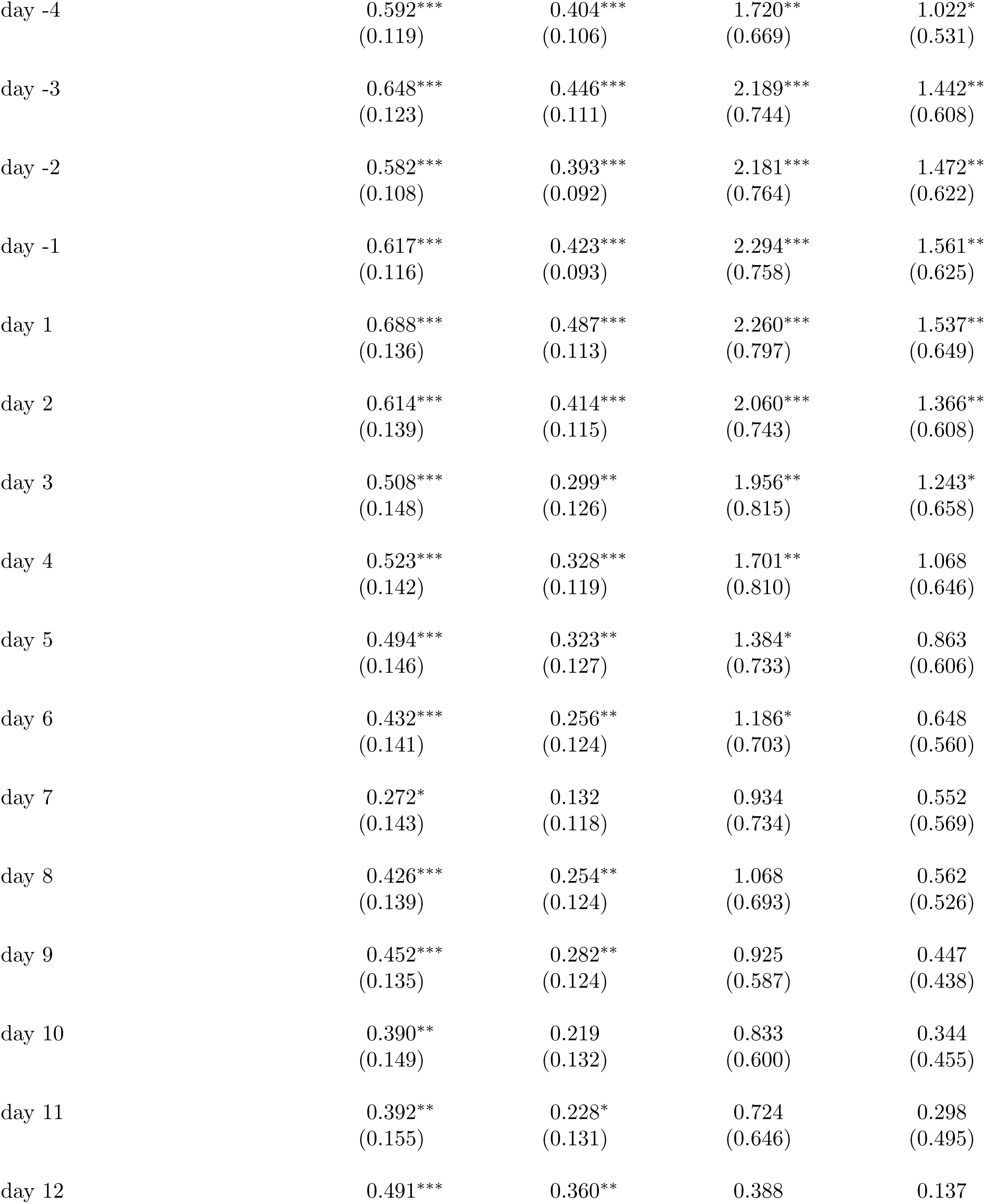
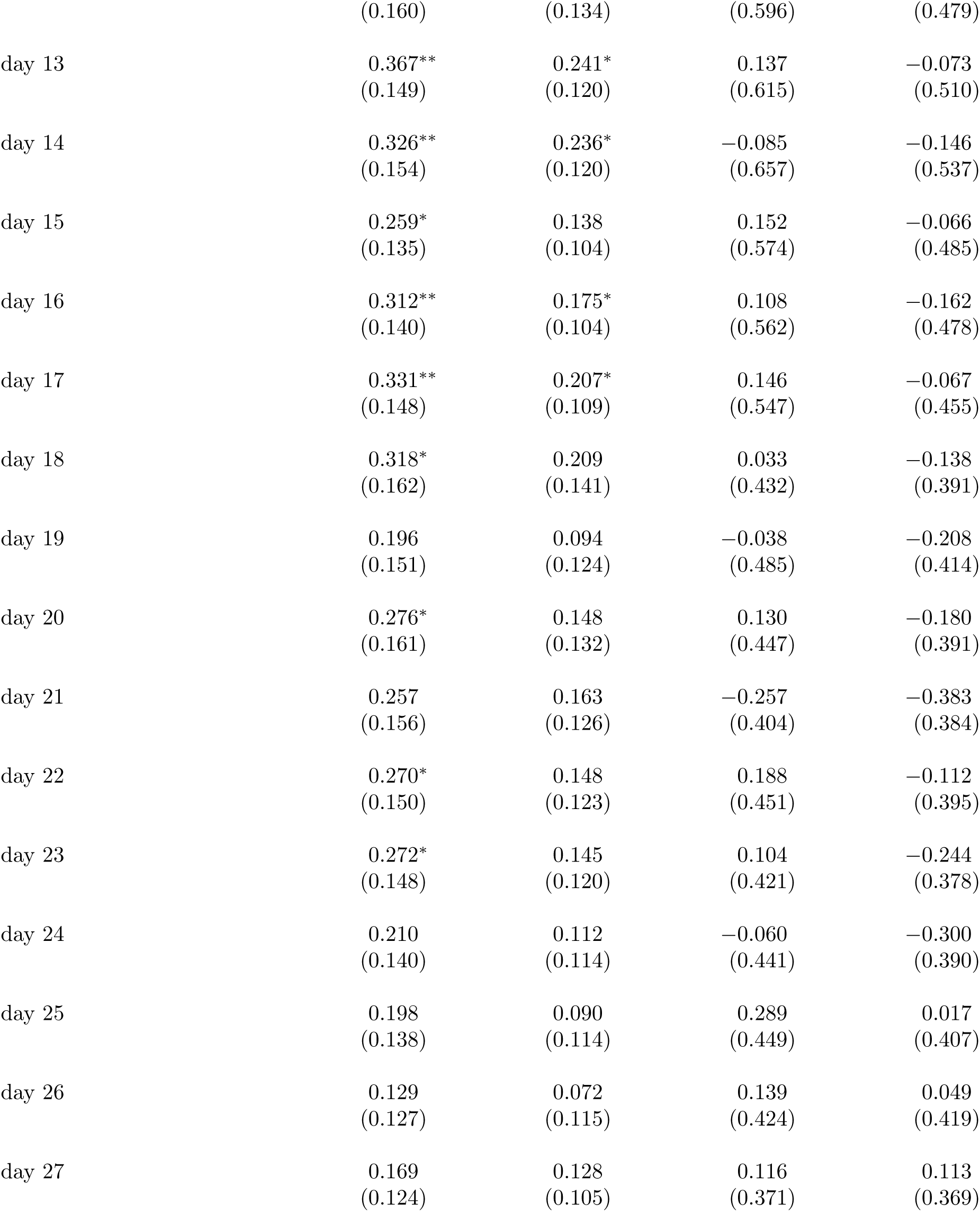
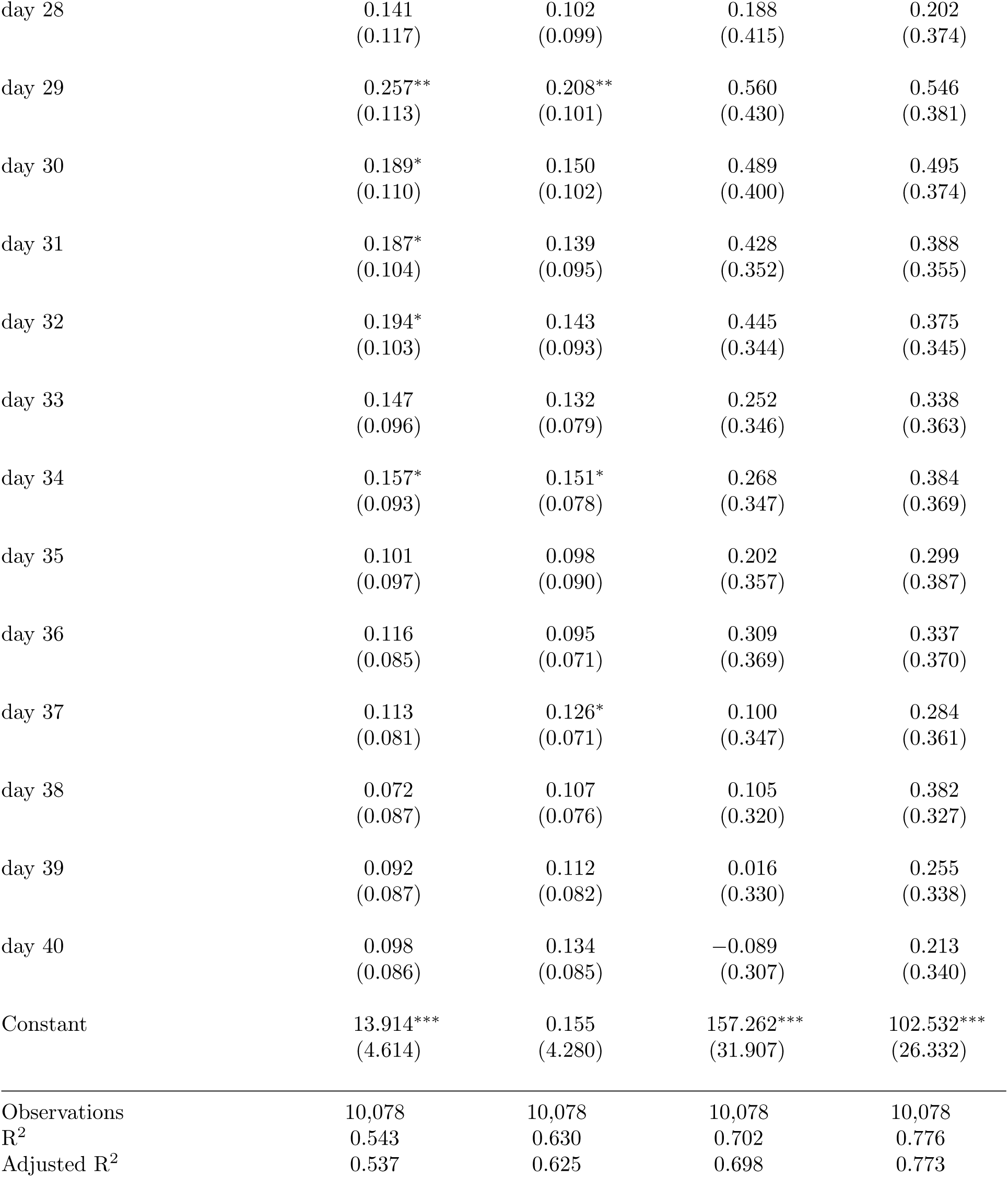
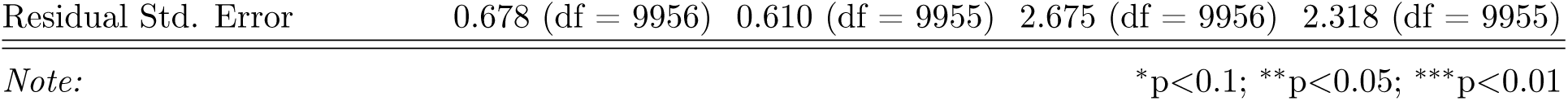
Event Study regression summary statistics for robustness check (fig. S6) where we compare the effect of mask mandates on deaths and hospitalization with and without controlling for cases.

|  | *Dependent variable:* |  |  |  |
| --- | --- | --- | --- | --- |
|  | deaths | deaths | hosp. | hosp. |
|  | (1) | (2) | (3) | (4) |
| confirmed cases |  | 0.410***<br>(0.022) |  | 1.990***<br>(0.243) |
| deaths delay | 0.507***<br>(0.031) | 0.439***<br>(0.031) |  |  |
| deaths growth | 0.001***<br>(0.0002) | 0.001***<br>(0.0001) |  |  |
| hospitalization delay |  |  | 0.549***<br>(0.027) | 0.387***<br>(0.027) |
| hospitalization growth |  |  | 0.109***<br>(0.032) | 0.082***<br>(0.030) |
| new test rate | 27.940**<br>(10.586) | −1.096<br>(6.646) | 154.787***<br>(43.311) | 31.262<br>(38.780) |
| precipitation | −0.004***<br>(0.001) | −0.004***<br>(0.001) | 0.002<br>(0.006) | 0.005<br>(0.006) |
| temperature | 0.002<br>(0.004) | −0.003<br>(0.004) | 0.055***<br>(0.015) | 0.023<br>(0.015) |
| retail & recreation | −2.338<br>(1.539) | 1.218<br>(1.097) | −35.993***<br>(11.299) | −20.360**<br>(9.758) |
| grocery & pharmacy | −0.025<br>(2.015) | −2.114<br>(1.760) | 12.745<br>(14.192) | 5.766<br>(12.601) |
| parks | 0.259***<br>(0.074) | 0.056<br>(0.069) | 1.210***<br>(0.319) | 0.339<br>(0.316) |
| transit stations | 0.007<br>(0.423) | 0.413<br>(0.441) | −4.089*<br>(2.081) | −2.765<br>(1.875) |
| workplaces | 2.085***<br>(0.704) | 2.317***<br>(0.549) | 10.502***<br>(3.450) | 12.020***<br>(2.733) |
| residential | −29.042***<br>(7.774) | −2.983<br>(7.367) | −279.358***<br>(52.035) | −174.851***<br>(42.791) |
| grocery & pharmacy squared | −0.308<br>(0.916) | 0.733<br>(0.822) | −5.955<br>(6.241) | −2.425<br>(5.565) |
| retail & recreation squared | 0.390<br>(0.860) | −1.315**<br>(0.640) | 16.584***<br>(5.968) | 8.999*<br>(5.230) |
| parks squared | −0.041**<br>(0.016) | 0.011<br>(0.017) | −0.233***<br>(0.064) | −0.001<br>(0.067) |
| transit stations squared | 0.168<br>(0.241) | −0.181<br>(0.261) | 2.857**<br>(1.187) | 1.532<br>(1.173) |
| workplaces squared | −1.463***<br>(0.512) | −1.471***<br>(0.404) | −9.818***<br>(2.276) | −10.316***<br>(1.907) |
| residential squared | 14.635***<br>(3.471) | 2.518<br>(3.255) | 129.688***<br>(23.620) | 80.931***<br>(19.252) |
| stay-at-home day | 0.0001**<br>(0.00003) | 0.00003<br>(0.00003) | −0.00000<br>(0.0001) | −0.0002<br>(0.0001) |
| business closure day | 0.0001***<br>(0.00003) | 0.0001***<br>(0.00003) | −0.00001<br>(0.0001) | −0.0001<br>(0.0001) |
| day -10 | 0.478***<br>(0.123) | 0.310***<br>(0.109) | 1.732***<br>(0.629) | 1.042**<br>(0.492) |
| day -9 | 0.463***<br>(0.133) | 0.325**<br>(0.123) | 1.534**<br>(0.642) | 0.995*<br>(0.525) |
| day -8 | 0.491***<br>(0.146) | 0.328**<br>(0.136) | 1.571**<br>(0.639) | 0.942*<br>(0.516) |
| day -7 | 0.553***<br>(0.130) | 0.395***<br>(0.126) | 1.372**<br>(0.632) | 0.795<br>(0.530) |
| day -6 | 0.559***<br>(0.132) | 0.374***<br>(0.123) | 1.693**<br>(0.698) | 1.007*<br>(0.556) |
| day -5 | 0.564***<br>(0.131) | 0.389***<br>(0.118) | 1.549**<br>(0.664) | 0.918*<br>(0.547) |
| day -4 | 0.592***<br>(0.119) | 0.404***<br>(0.106) | 1.720**<br>(0.669) | 1.022*<br>(0.531) |
| day -3 | 0.648***<br>(0.123) | 0.446***<br>(0.111) | 2.189***<br>(0.744) | 1.442**<br>(0.608) |
| day -2 | 0.582***<br>(0.108) | 0.393***<br>(0.092) | 2.181***<br>(0.764) | 1.472**<br>(0.622) |
| day -1 | 0.617***<br>(0.116) | 0.423***<br>(0.093) | 2.294***<br>(0.758) | 1.561**<br>(0.625) |
| day 1 | 0.688***<br>(0.136) | 0.487***<br>(0.113) | 2.260***<br>(0.797) | 1.537**<br>(0.649) |
| day 2 | 0.614***<br>(0.139) | 0.414***<br>(0.115) | 2.060***<br>(0.743) | 1.366**<br>(0.608) |
| day 3 | 0.508***<br>(0.148) | 0.299**<br>(0.126) | 1.956**<br>(0.815) | 1.243*<br>(0.658) |
| day 4 | 0.523***<br>(0.142) | 0.328***<br>(0.119) | 1.701**<br>(0.810) | 1.068<br>(0.646) |
| day 5 | 0.494***<br>(0.146) | 0.323**<br>(0.127) | 1.384*<br>(0.733) | 0.863<br>(0.606) |
| day 6 | 0.432***<br>(0.141) | 0.256**<br>(0.124) | 1.186*<br>(0.703) | 0.648<br>(0.560) |
| day 7 | 0.272*<br>(0.143) | 0.132<br>(0.118) | 0.934<br>(0.734) | 0.552<br>(0.569) |
| day 8 | 0.426***<br>(0.139) | 0.254**<br>(0.124) | 1.068<br>(0.693) | 0.562<br>(0.526) |
| day 9 | 0.452***<br>(0.135) | 0.282**<br>(0.124) | 0.925<br>(0.587) | 0.447<br>(0.438) |
| day 10 | 0.390**<br>(0.149) | 0.219<br>(0.132) | 0.833<br>(0.600) | 0.344<br>(0.455) |
| day 11 | 0.392**<br>(0.155) | 0.228*<br>(0.131) | 0.724<br>(0.646) | 0.298<br>(0.495) |
| day 12 | 0.491*** | 0.360** | 0.388 | 0.137 |
|  | (0.160) | (0.134) | (0.596) | (0.479) |
| day 13 | 0.367**<br>(0.149) | 0.241*<br>(0.120) | 0.137<br>(0.615) | −0.073<br>(0.510) |
| day 14 | 0.326**<br>(0.154) | 0.236*<br>(0.120) | −0.085<br>(0.657) | −0.146<br>(0.537) |
| day 15 | 0.259*<br>(0.135) | 0.138<br>(0.104) | 0.152<br>(0.574) | −0.066<br>(0.485) |
| day 16 | 0.312**<br>(0.140) | 0.175*<br>(0.104) | 0.108<br>(0.562) | −0.162<br>(0.478) |
| day 17 | 0.331**<br>(0.148) | 0.207*<br>(0.109) | 0.146<br>(0.547) | −0.067<br>(0.455) |
| day 18 | 0.318*<br>(0.162) | 0.209<br>(0.141) | 0.033<br>(0.432) | −0.138<br>(0.391) |
| day 19 | 0.196<br>(0.151) | 0.094<br>(0.124) | −0.038<br>(0.485) | −0.208<br>(0.414) |
| day 20 | 0.276*<br>(0.161) | 0.148<br>(0.132) | 0.130<br>(0.447) | −0.180<br>(0.391) |
| day 21 | 0.257<br>(0.156) | 0.163<br>(0.126) | −0.257<br>(0.404) | −0.383<br>(0.384) |
| day 22 | 0.270*<br>(0.150) | 0.148<br>(0.123) | 0.188<br>(0.451) | −0.112<br>(0.395) |
| day 23 | 0.272*<br>(0.148) | 0.145<br>(0.120) | 0.104<br>(0.421) | −0.244<br>(0.378) |
| day 24 | 0.210<br>(0.140) | 0.112<br>(0.114) | −0.060<br>(0.441) | −0.300<br>(0.390) |
| day 25 | 0.198<br>(0.138) | 0.090<br>(0.114) | 0.289<br>(0.449) | 0.017<br>(0.407) |
| day 26 | 0.129<br>(0.127) | 0.072<br>(0.115) | 0.139<br>(0.424) | 0.049<br>(0.419) |
| day 27 | 0.169<br>(0.124) | 0.128<br>(0.105) | 0.116<br>(0.371) | 0.113<br>(0.369) |
| day 28 | 0.141<br>(0.117) | 0.102<br>(0.099) | 0.188<br>(0.415) | 0.202<br>(0.374) |
| day 29 | 0.257**<br>(0.113) | 0.208**<br>(0.101) | 0.560<br>(0.430) | 0.546<br>(0.381) |
| day 30 | 0.189*<br>(0.110) | 0.150<br>(0.102) | 0.489<br>(0.400) | 0.495<br>(0.374) |
| day 31 | 0.187*<br>(0.104) | 0.139<br>(0.095) | 0.428<br>(0.352) | 0.388<br>(0.355) |
| day 32 | 0.194*<br>(0.103) | 0.143<br>(0.093) | 0.445<br>(0.344) | 0.375<br>(0.345) |
| day 33 | 0.147<br>(0.096) | 0.132<br>(0.079) | 0.252<br>(0.346) | 0.338<br>(0.363) |
| day 34 | 0.157*<br>(0.093) | 0.151*<br>(0.078) | 0.268<br>(0.347) | 0.384<br>(0.369) |
| day 35 | 0.101<br>(0.097) | 0.098<br>(0.090) | 0.202<br>(0.357) | 0.299<br>(0.387) |
| day 36 | 0.116<br>(0.085) | 0.095<br>(0.071) | 0.309<br>(0.369) | 0.337<br>(0.370) |
| day 37 | 0.113<br>(0.081) | 0.126*<br>(0.071) | 0.100<br>(0.347) | 0.284<br>(0.361) |
| day 38 | 0.072<br>(0.087) | 0.107<br>(0.076) | 0.105<br>(0.320) | 0.382<br>(0.327) |
| day 39 | 0.092<br>(0.087) | 0.112<br>(0.082) | 0.016<br>(0.330) | 0.255<br>(0.338) |
| day 40 | 0.098<br>(0.086) | 0.134<br>(0.085) | -0.089<br>(0.307) | 0.213<br>(0.340) |
| Constant | 13.914***<br>(4.614) | 0.155<br>(4.280) | 157.262***<br>(31.907) | 102.532***<br>(26.332) |
| Observations | 10,078 | 10,078 | 10,078 | 10,078 |
| R<sup>2</sup> | 0.543 | 0.630 | 0.702 | 0.776 |
| Adjusted R<sup>2</sup> | 0.537 | 0.625 | 0.698 | 0.773 |
| Residual Std. Error | 0.678 (df = 9956) | 0.610 (df = 9955) | 2.675 (df = 9956) | 2.318 (df = 9955) |

*Note:*
\*p<0.1; \*\*p<0.05; \*\*\*p<0.01

**Table S8:**
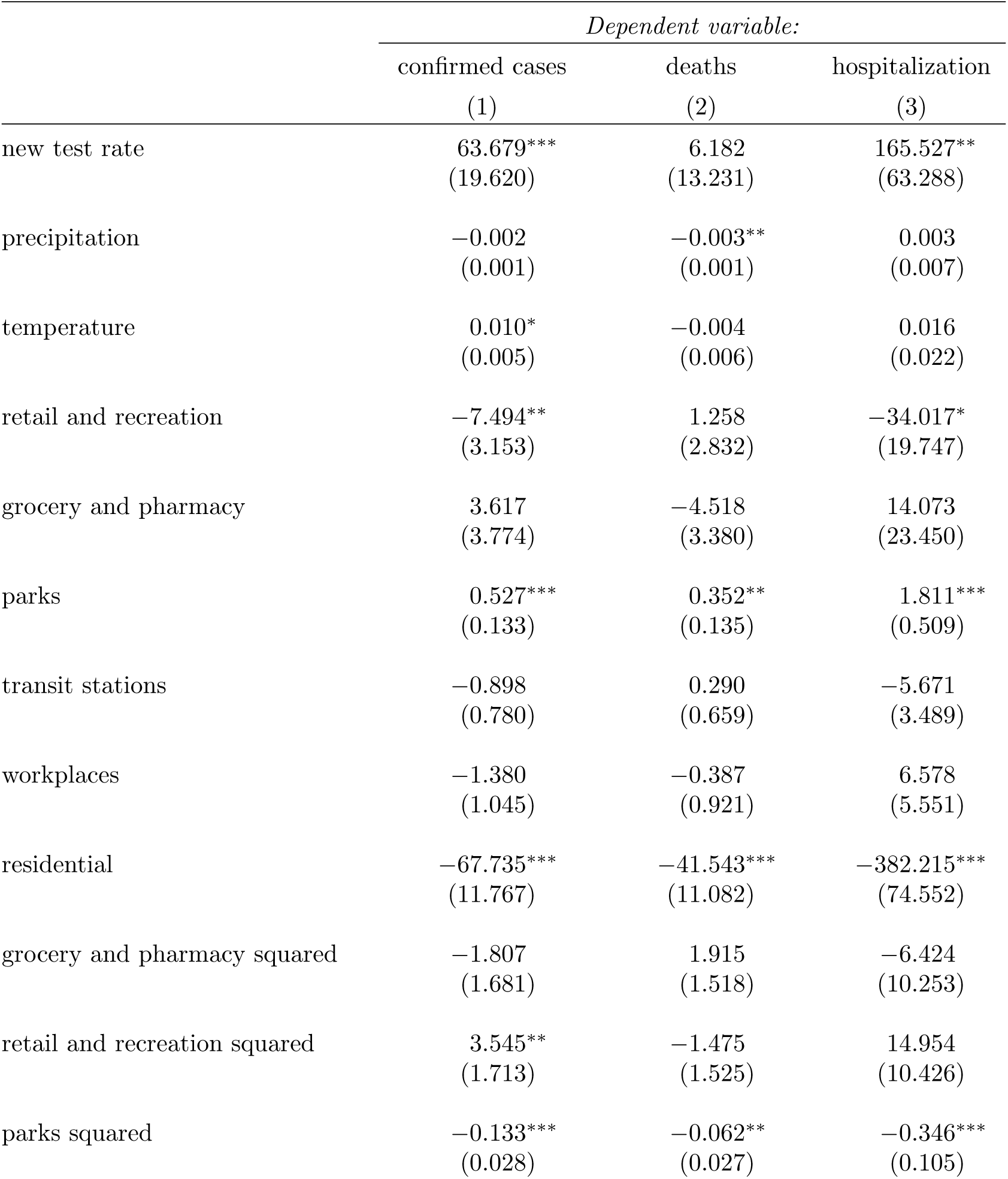
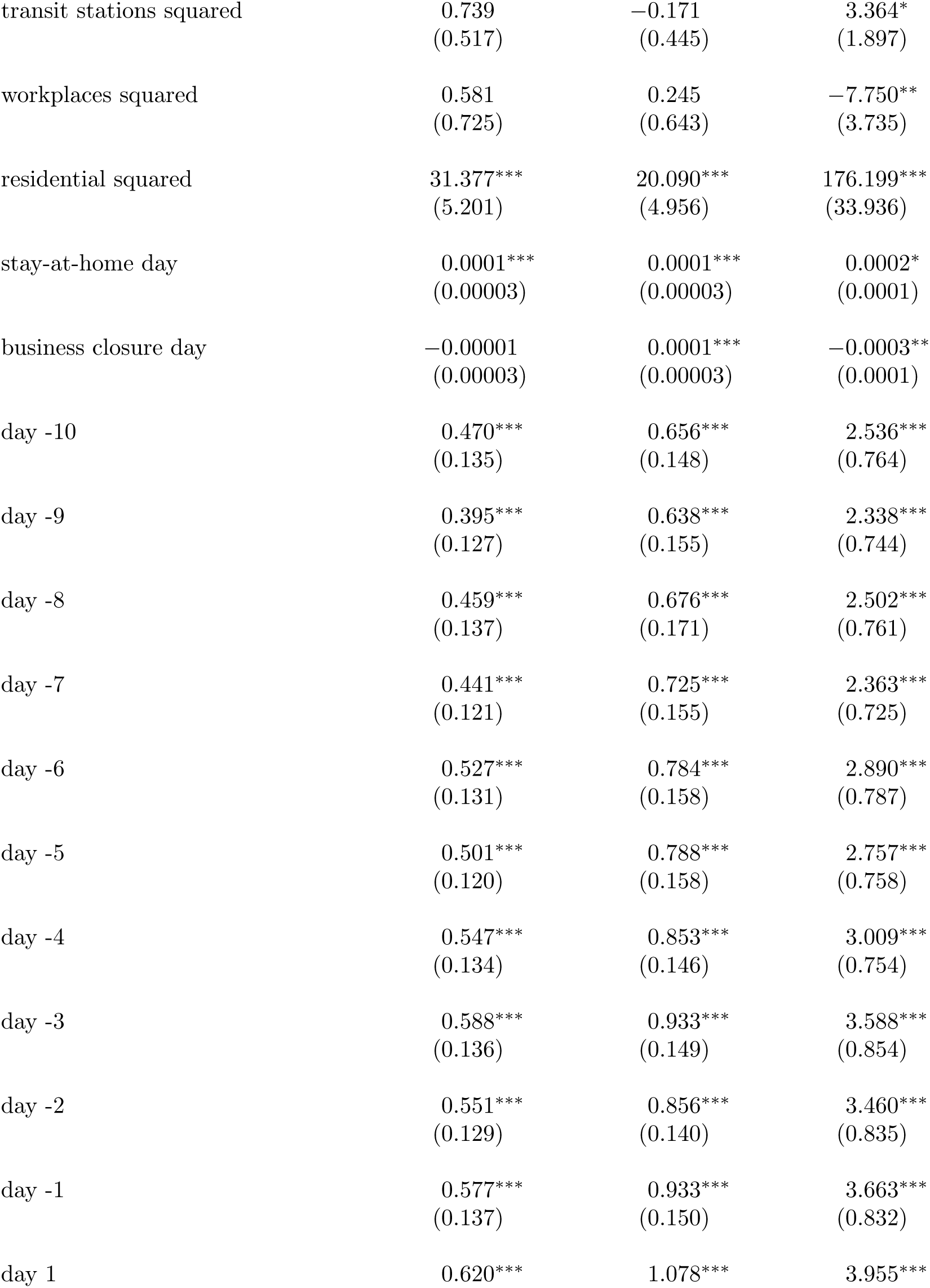
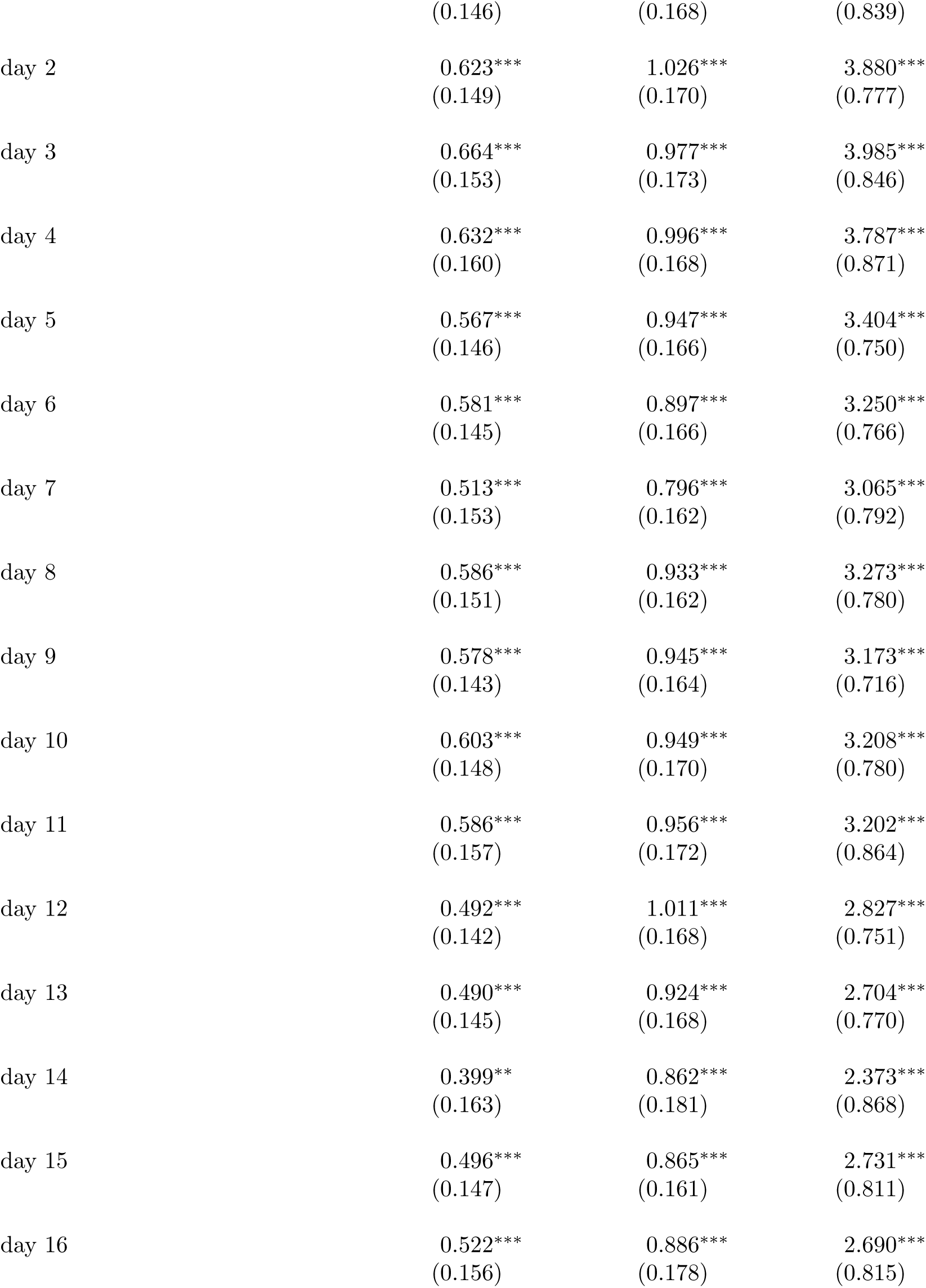
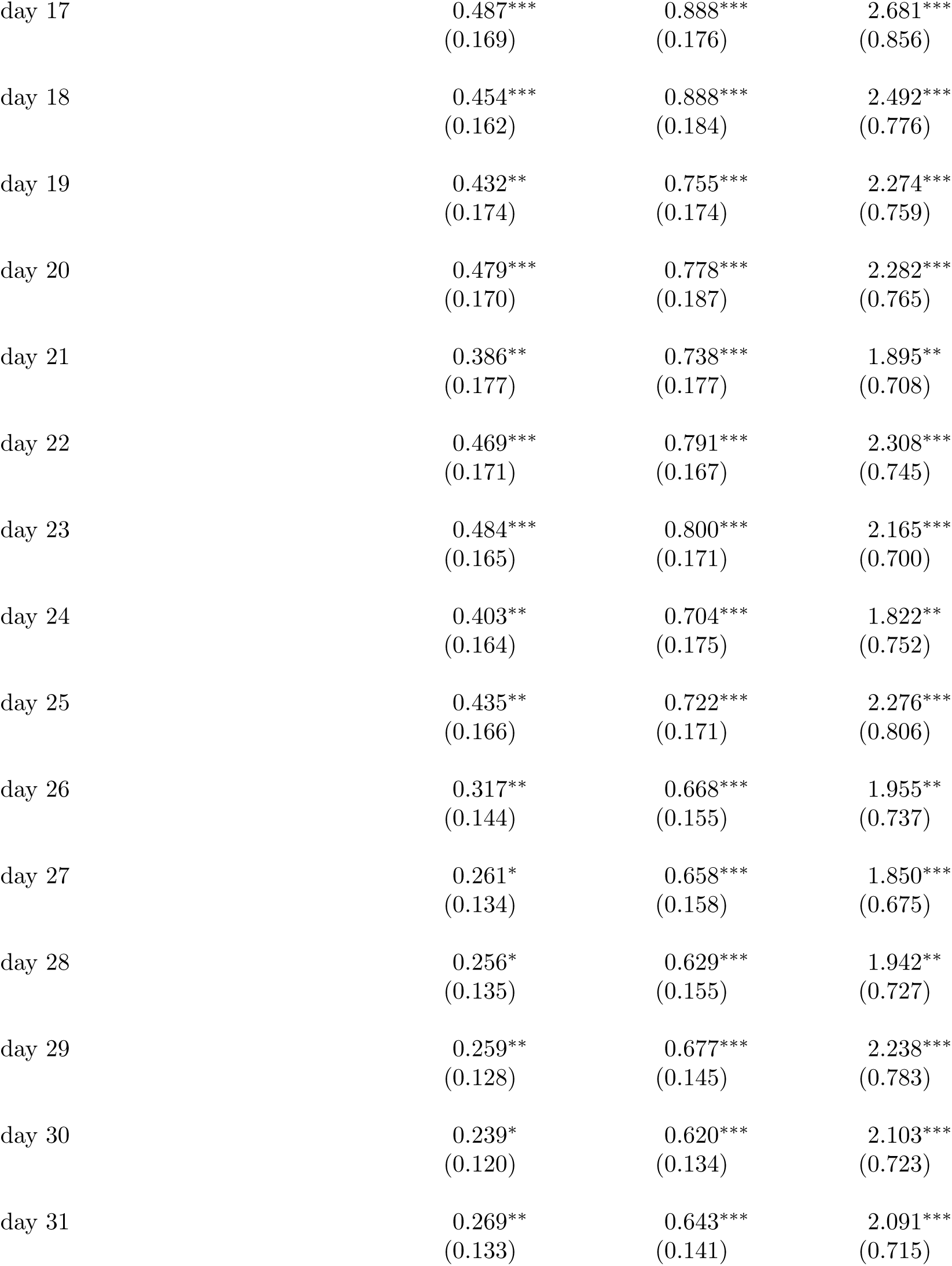
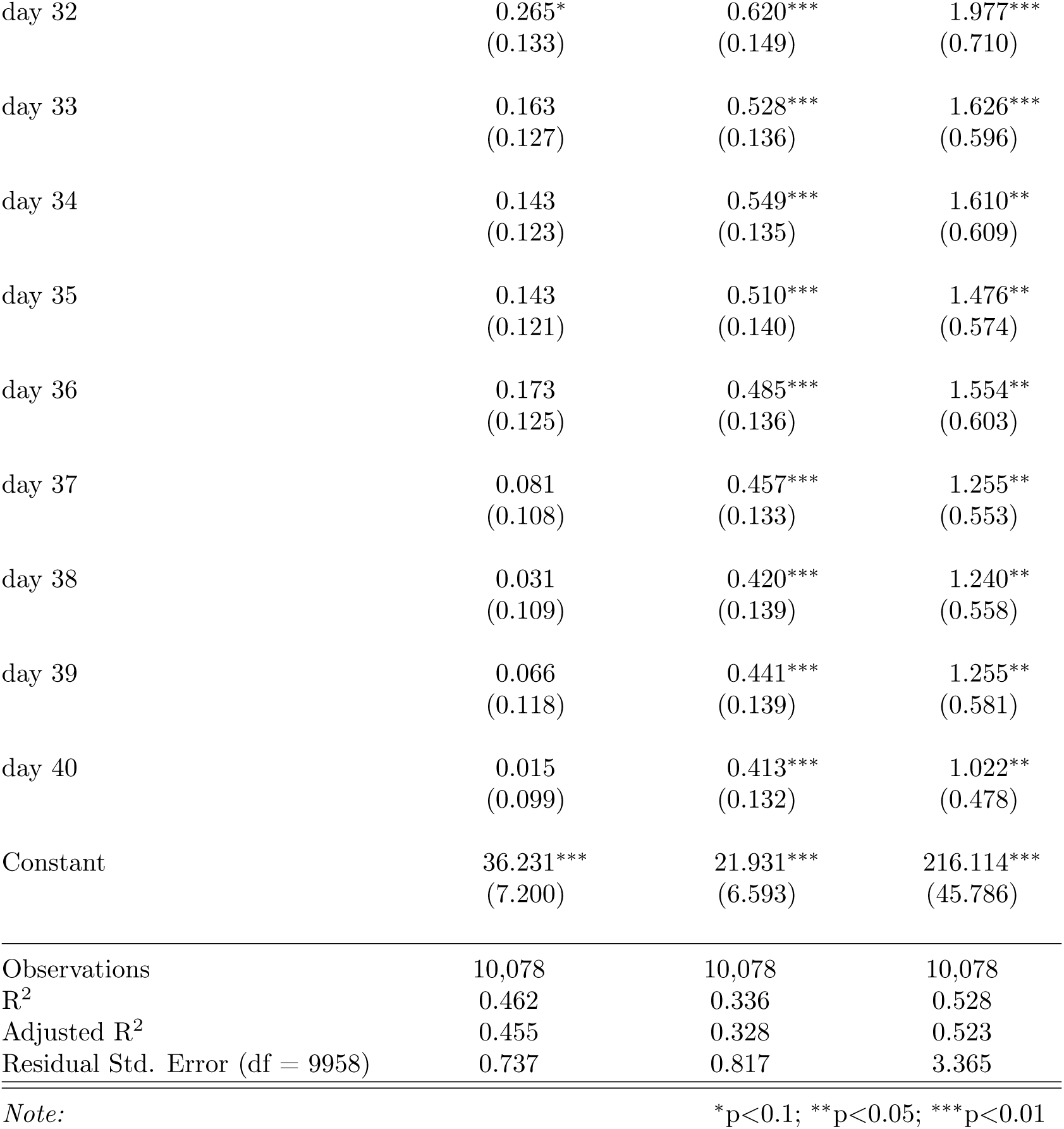
Event Study regression summary statistics for robustness check (Fig S1) where we **do not** control for past outcome values (with a delay of 14 days) and growth rates as a way to minimize confounding due to peoples private behavioral changes to COVID-19 [21].

|  | *Dependent variable:* |  |  |
| --- | --- | --- | --- |
|  | confirmed cases | deaths | hospitalization |
|  | (1) | (2) | (3) |
| new test rate | 63.679***<br>(19.620) | 6.182<br>(13.231) | 165.527**<br>(63.288) |
| precipitation | −0.002<br>(0.001) | −0.003**<br>(0.001) | 0.003<br>(0.007) |
| temperature | 0.010*<br>(0.005) | −0.004<br>(0.006) | 0.016<br>(0.022) |
| retail and recreation | −7.494**<br>(3.153) | 1.258<br>(2.832) | −34.017*<br>(19.747) |
| grocery and pharmacy | 3.617<br>(3.774) | −4.518<br>(3.380) | 14.073<br>(23.450) |
| parks | 0.527***<br>(0.133) | 0.352**<br>(0.135) | 1.811***<br>(0.509) |
| transit stations | −0.898<br>(0.780) | 0.290<br>(0.659) | −5.671<br>(3.489) |
| workplaces | −1.380<br>(1.045) | −0.387<br>(0.921) | 6.578<br>(5.551) |
| residential | −67.735***<br>(11.767) | −41.543***<br>(11.082) | −382.215***<br>(74.552) |
| grocery and pharmacy squared | −1.807<br>(1.681) | 1.915<br>(1.518) | −6.424<br>(10.253) |
| retail and recreation squared | 3.545**<br>(1.713) | −1.475<br>(1.525) | 14.954<br>(10.426) |
| parks squared | −0.133***<br>(0.028) | −0.062**<br>(0.027) | −0.346***<br>(0.105) |
| transit stations squared | 0.739<br>(0.517) | −0.171<br>(0.445) | 3.364*<br>(1.897) |
| workplaces squared | 0.581<br>(0.725) | 0.245<br>(0.643) | −7.750**<br>(3.735) |
| residential squared | 31.377***<br>(5.201) | 20.090***<br>(4.956) | 176.199***<br>(33.936) |
| stay-at-home day | 0.0001***<br>(0.00003) | 0.0001***<br>(0.00003) | 0.0002*<br>(0.0001) |
| business closure day | −0.00001<br>(0.00003) | 0.0001***<br>(0.00003) | −0.0003**<br>(0.0001) |
| day -10 | 0.470***<br>(0.135) | 0.656***<br>(0.148) | 2.536***<br>(0.764) |
| day -9 | 0.395***<br>(0.127) | 0.638***<br>(0.155) | 2.338***<br>(0.744) |
| day -8 | 0.459***<br>(0.137) | 0.676***<br>(0.171) | 2.502***<br>(0.761) |
| day -7 | 0.441***<br>(0.121) | 0.725***<br>(0.155) | 2.363***<br>(0.725) |
| day -6 | 0.527***<br>(0.131) | 0.784***<br>(0.158) | 2.890***<br>(0.787) |
| day -5 | 0.501***<br>(0.120) | 0.788***<br>(0.158) | 2.757***<br>(0.758) |
| day -4 | 0.547***<br>(0.134) | 0.853***<br>(0.146) | 3.009***<br>(0.754) |
| day -3 | 0.588***<br>(0.136) | 0.933***<br>(0.149) | 3.588***<br>(0.854) |
| day -2 | 0.551***<br>(0.129) | 0.856***<br>(0.140) | 3.460***<br>(0.835) |
| day -1 | 0.577***<br>(0.137) | 0.933***<br>(0.150) | 3.663***<br>(0.832) |
| day 1 | 0.620*** | 1.078*** | 3.955*** |
|  | (0.146) | (0.168) | (0.839) |
| day 2 | 0.623***<br>(0.149) | 1.026***<br>(0.170) | 3.880***<br>(0.777) |
| day 3 | 0.664***<br>(0.153) | 0.977***<br>(0.173) | 3.985***<br>(0.846) |
| day 4 | 0.632***<br>(0.160) | 0.996***<br>(0.168) | 3.787***<br>(0.871) |
| day 5 | 0.567***<br>(0.146) | 0.947***<br>(0.166) | 3.404***<br>(0.750) |
| day 6 | 0.581***<br>(0.145) | 0.897***<br>(0.166) | 3.250***<br>(0.766) |
| day 7 | 0.513***<br>(0.153) | 0.796***<br>(0.162) | 3.065***<br>(0.792) |
| day 8 | 0.586***<br>(0.151) | 0.933***<br>(0.162) | 3.273***<br>(0.780) |
| day 9 | 0.578***<br>(0.143) | 0.945***<br>(0.164) | 3.173***<br>(0.716) |
| day 10 | 0.603***<br>(0.148) | 0.949***<br>(0.170) | 3.208***<br>(0.780) |
| day 11 | 0.586***<br>(0.157) | 0.956***<br>(0.172) | 3.202***<br>(0.864) |
| day 12 | 0.492***<br>(0.142) | 1.011***<br>(0.168) | 2.827***<br>(0.751) |
| day 13 | 0.490***<br>(0.145) | 0.924***<br>(0.168) | 2.704***<br>(0.770) |
| day 14 | 0.399**<br>(0.163) | 0.862***<br>(0.181) | 2.373***<br>(0.868) |
| day 15 | 0.496***<br>(0.147) | 0.865***<br>(0.161) | 2.731***<br>(0.811) |
| day 16 | 0.522***<br>(0.156) | 0.886***<br>(0.178) | 2.690***<br>(0.815) |
| day 17 | 0.487***<br>(0.169) | 0.888***<br>(0.176) | 2.681***<br>(0.856) |
| day 18 | 0.454***<br>(0.162) | 0.888***<br>(0.184) | 2.492***<br>(0.776) |
| day 19 | 0.432**<br>(0.174) | 0.755***<br>(0.174) | 2.274***<br>(0.759) |
| day 20 | 0.479***<br>(0.170) | 0.778***<br>(0.187) | 2.282***<br>(0.765) |
| day 21 | 0.386**<br>(0.177) | 0.738***<br>(0.177) | 1.895**<br>(0.708) |
| day 22 | 0.469***<br>(0.171) | 0.791***<br>(0.167) | 2.308***<br>(0.745) |
| day 23 | 0.484***<br>(0.165) | 0.800***<br>(0.171) | 2.165***<br>(0.700) |
| day 24 | 0.403**<br>(0.164) | 0.704***<br>(0.175) | 1.822**<br>(0.752) |
| day 25 | 0.435**<br>(0.166) | 0.722***<br>(0.171) | 2.276***<br>(0.806) |
| day 26 | 0.317**<br>(0.144) | 0.668***<br>(0.155) | 1.955**<br>(0.737) |
| day 27 | 0.261*<br>(0.134) | 0.658***<br>(0.158) | 1.850***<br>(0.675) |
| day 28 | 0.256*<br>(0.135) | 0.629***<br>(0.155) | 1.942**<br>(0.727) |
| day 29 | 0.259**<br>(0.128) | 0.677***<br>(0.145) | 2.238***<br>(0.783) |
| day 30 | 0.239*<br>(0.120) | 0.620***<br>(0.134) | 2.103***<br>(0.723) |
| day 31 | 0.269**<br>(0.133) | 0.643***<br>(0.141) | 2.091***<br>(0.715) |
| day 32 | 0.265*<br>(0.133) | 0.620***<br>(0.149) | 1.977***<br>(0.710) |
| day 33 | 0.163<br>(0.127) | 0.528***<br>(0.136) | 1.626***<br>(0.596) |
| day 34 | 0.143<br>(0.123) | 0.549***<br>(0.135) | 1.610**<br>(0.609) |
| day 35 | 0.143<br>(0.121) | 0.510***<br>(0.140) | 1.476**<br>(0.574) |
| day 36 | 0.173<br>(0.125) | 0.485***<br>(0.136) | 1.554**<br>(0.603) |
| day 37 | 0.081<br>(0.108) | 0.457***<br>(0.133) | 1.255**<br>(0.553) |
| day 38 | 0.031<br>(0.109) | 0.420***<br>(0.139) | 1.240**<br>(0.558) |
| day 39 | 0.066<br>(0.118) | 0.441***<br>(0.139) | 1.255**<br>(0.581) |
| day 40 | 0.015<br>(0.099) | 0.413***<br>(0.132) | 1.022**<br>(0.478) |
| Constant | 36.231***<br>(7.200) | 21.931***<br>(6.593) | 216.114***<br>(45.786) |
| Observations | 10,078 | 10,078 | 10,078 |
| R<sup>2</sup> | 0.462 | 0.336 | 0.528 |
| Adjusted R<sup>2</sup> | 0.455 | 0.328 | 0.523 |
| Residual Std. Error (df = 9958) | 0.737 | 0.817 | 3.365 |

*Note:*
\*p<0.1; \*\*p<0.05; \*\*\*p<0.01

**Table S9:**
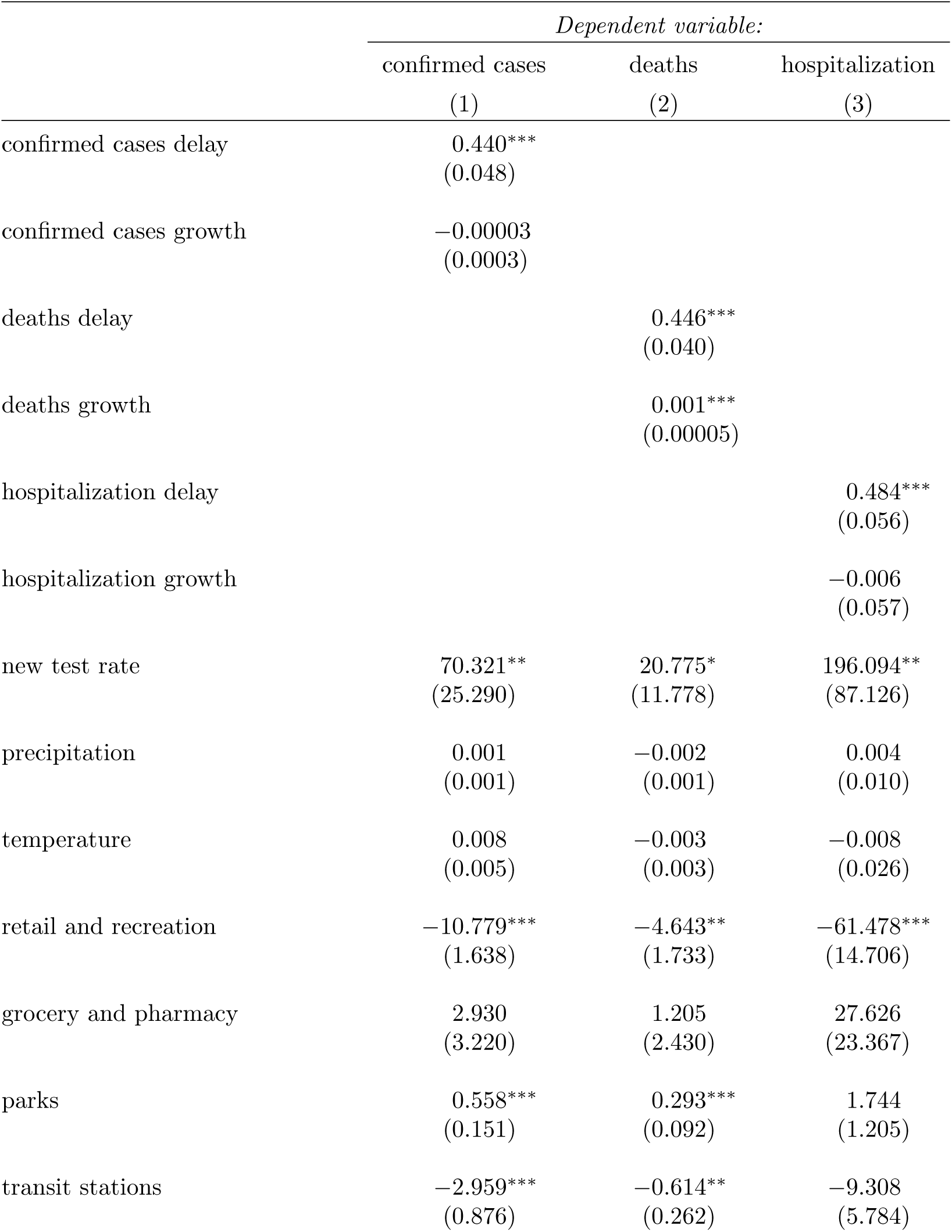
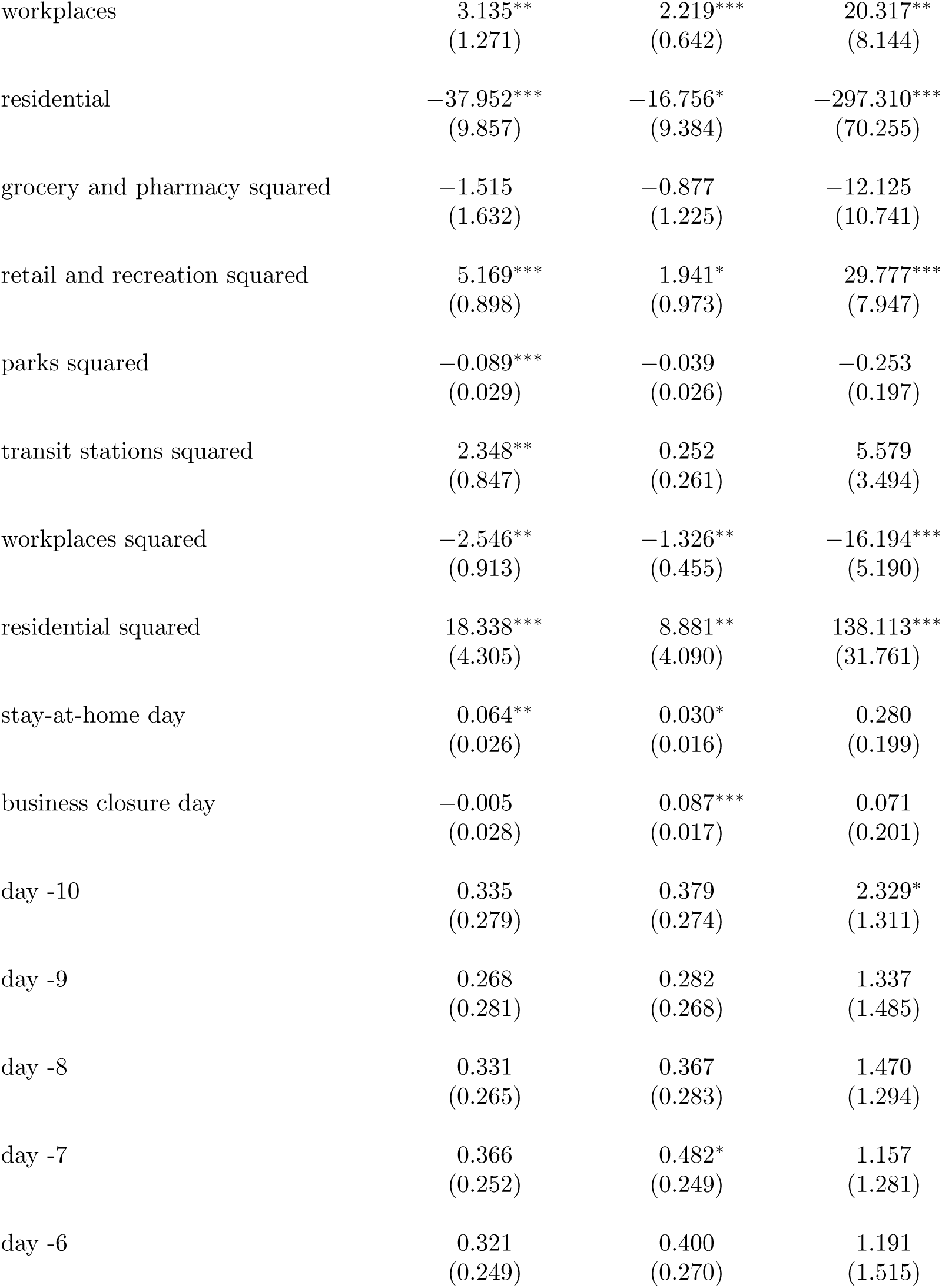
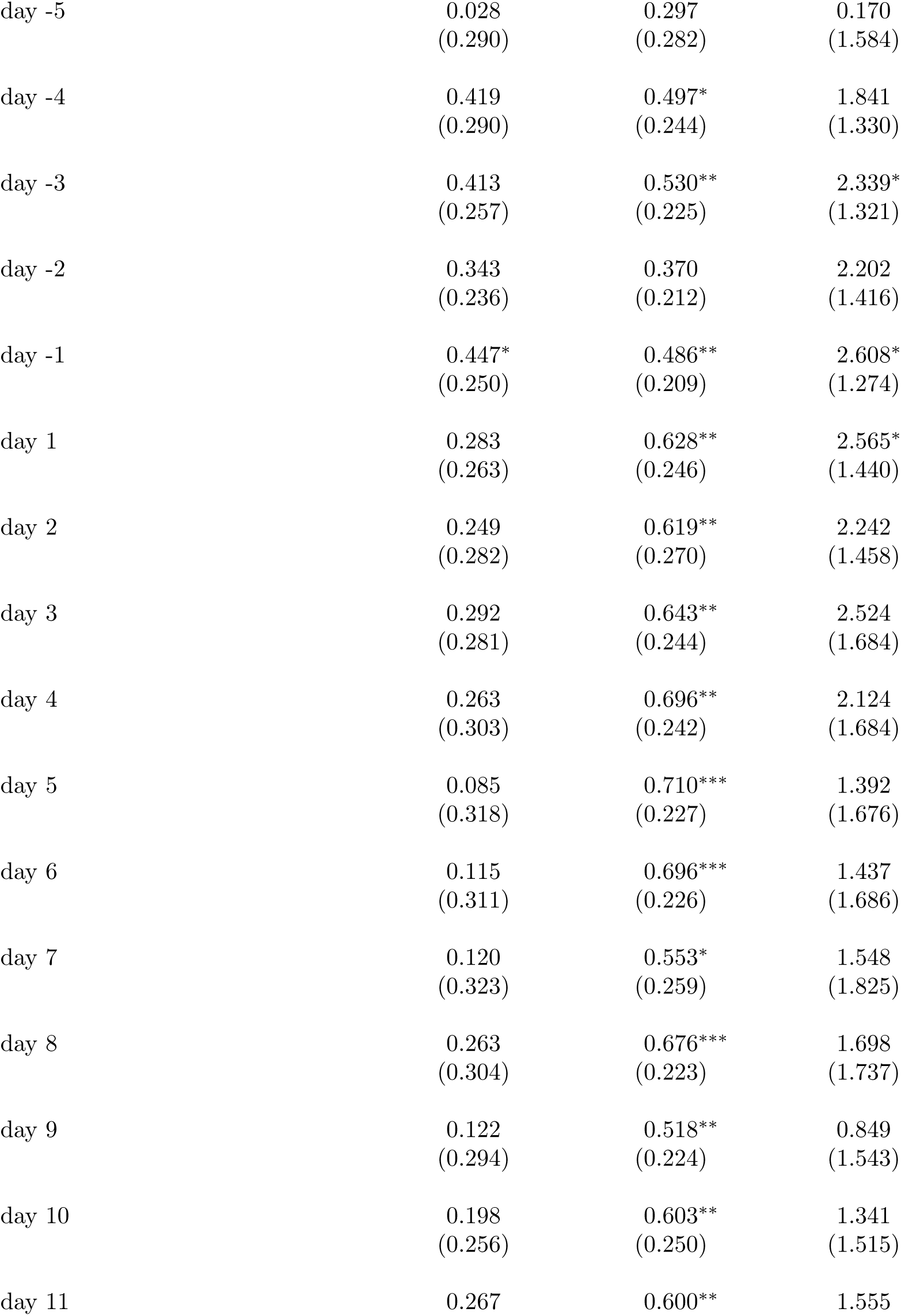
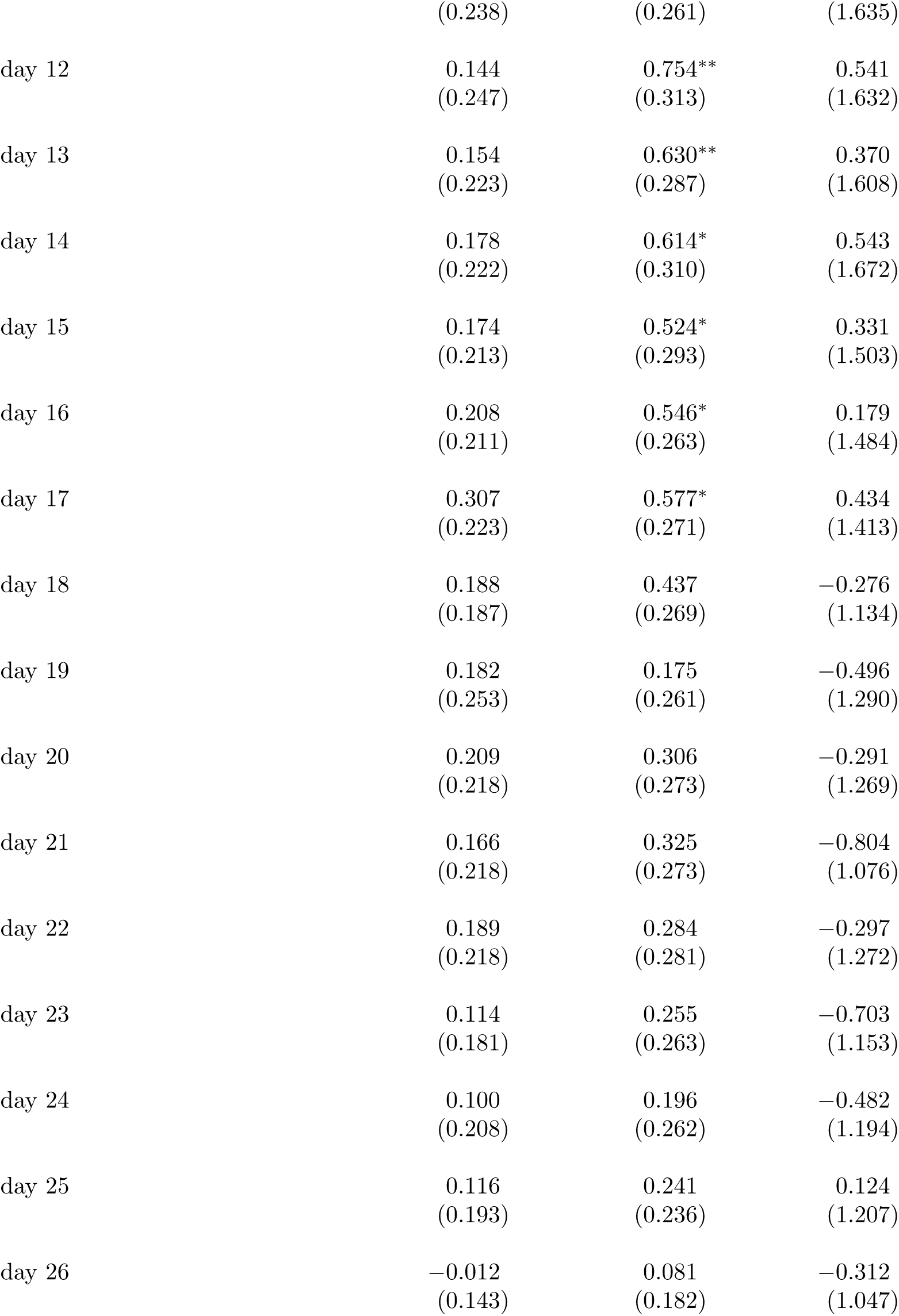
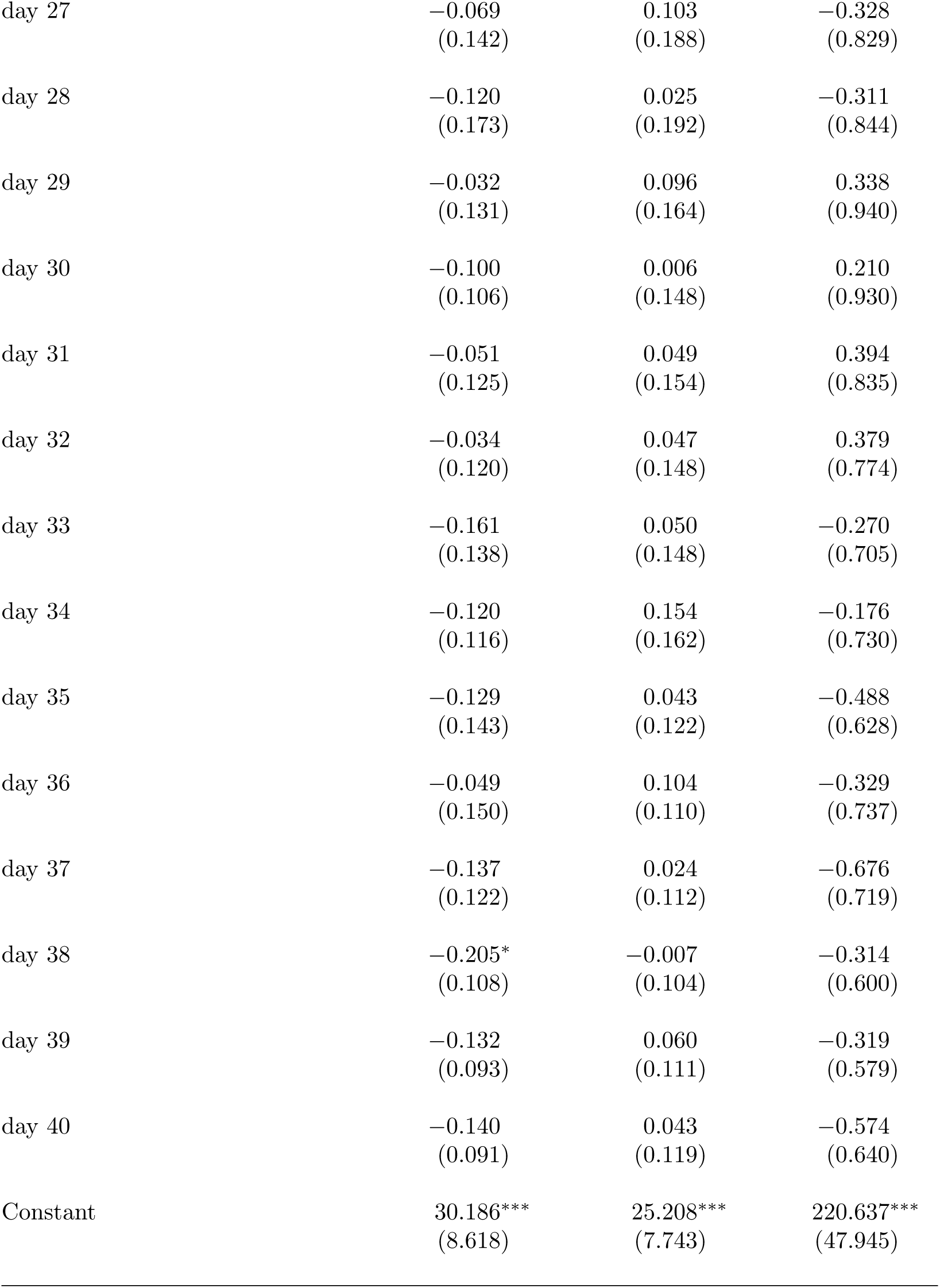
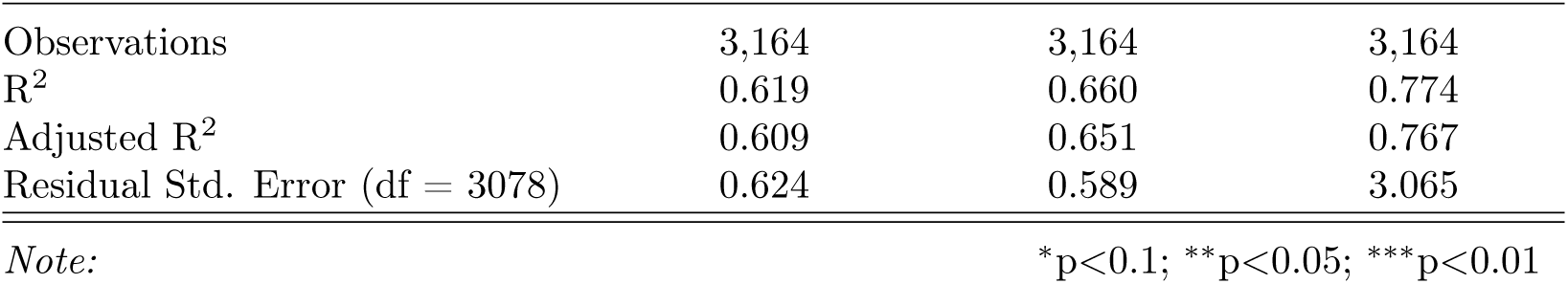
Event Study regression summary statistics of fig. 3 (for earlier states only, i.e. NY, NJ, MA, CT, MI, DC, RI, IL, WA, PA, GA, VT, MD, FL, LA).

|  | *Dependent variable:* |  |  |
| --- | --- | --- | --- |
|  | confirmed cases | deaths | hospitalization |
|  | (1) | (2) | (3) |
| confirmed cases delay | 0.440***<br>(0.048) |  |  |
| confirmed cases growth | −0.00003<br>(0.0003) |  |  |
| deaths delay |  | 0.446***<br>(0.040) |  |
| deaths growth |  | 0.001***<br>(0.00005) |  |
| hospitalization delay |  |  | 0.484***<br>(0.056) |
| hospitalization growth |  |  | −0.006<br>(0.057) |
| new test rate | 70.321**<br>(25.290) | 20.775*<br>(11.778) | 196.094**<br>(87.126) |
| precipitation | 0.001<br>(0.001) | −0.002<br>(0.001) | 0.004<br>(0.010) |
| temperature | 0.008<br>(0.005) | −0.003<br>(0.003) | −0.008<br>(0.026) |
| retail and recreation | −10.779***<br>(1.638) | −4.643**<br>(1.733) | −61.478***<br>(14.706) |
| grocery and pharmacy | 2.930<br>(3.220) | 1.205<br>(2.430) | 27.626<br>(23.367) |
| parks | 0.558***<br>(0.151) | 0.293***<br>(0.092) | 1.744<br>(1.205) |
| transit stations | −2.959***<br>(0.876) | −0.614**<br>(0.262) | −9.308<br>(5.784) |
| workplaces | 3.135**<br>(1.271) | 2.219***<br>(0.642) | 20.317**<br>(8.144) |
| residential | -37.952***<br>(9.857) | -16.756*<br>(9.384) | -297.310***<br>(70.255) |
| grocery and pharmacy squared | -1.515<br>(1.632) | -0.877<br>(1.225) | -12.125<br>(10.741) |
| retail and recreation squared | 5.169***<br>(0.898) | 1.941*<br>(0.973) | 29.777***<br>(7.947) |
| parks squared | -0.089***<br>(0.029) | -0.039<br>(0.026) | -0.253<br>(0.197) |
| transit stations squared | 2.348**<br>(0.847) | 0.252<br>(0.261) | 5.579<br>(3.494) |
| workplaces squared | -2.546**<br>(0.913) | -1.326**<br>(0.455) | -16.194***<br>(5.190) |
| residential squared | 18.338***<br>(4.305) | 8.881**<br>(4.090) | 138.113***<br>(31.761) |
| stay-at-home day | 0.064**<br>(0.026) | 0.030*<br>(0.016) | 0.280<br>(0.199) |
| business closure day | -0.005<br>(0.028) | 0.087***<br>(0.017) | 0.071<br>(0.201) |
| day -10 | 0.335<br>(0.279) | 0.379<br>(0.274) | 2.329*<br>(1.311) |
| day -9 | 0.268<br>(0.281) | 0.282<br>(0.268) | 1.337<br>(1.485) |
| day -8 | 0.331<br>(0.265) | 0.367<br>(0.283) | 1.470<br>(1.294) |
| day -7 | 0.366<br>(0.252) | 0.482*<br>(0.249) | 1.157<br>(1.281) |
| day -6 | 0.321<br>(0.249) | 0.400<br>(0.270) | 1.191<br>(1.515) |
| day -5 | 0.028<br>(0.290) | 0.297<br>(0.282) | 0.170<br>(1.584) |
| day -4 | 0.419<br>(0.290) | 0.497*<br>(0.244) | 1.841<br>(1.330) |
| day -3 | 0.413<br>(0.257) | 0.530**<br>(0.225) | 2.339*<br>(1.321) |
| day -2 | 0.343<br>(0.236) | 0.370<br>(0.212) | 2.202<br>(1.416) |
| day -1 | 0.447*<br>(0.250) | 0.486**<br>(0.209) | 2.608*<br>(1.274) |
| day 1 | 0.283<br>(0.263) | 0.628**<br>(0.246) | 2.565*<br>(1.440) |
| day 2 | 0.249<br>(0.282) | 0.619**<br>(0.270) | 2.242<br>(1.458) |
| day 3 | 0.292<br>(0.281) | 0.643**<br>(0.244) | 2.524<br>(1.684) |
| day 4 | 0.263<br>(0.303) | 0.696**<br>(0.242) | 2.124<br>(1.684) |
| day 5 | 0.085<br>(0.318) | 0.710***<br>(0.227) | 1.392<br>(1.676) |
| day 6 | 0.115<br>(0.311) | 0.696***<br>(0.226) | 1.437<br>(1.686) |
| day 7 | 0.120<br>(0.323) | 0.553*<br>(0.259) | 1.548<br>(1.825) |
| day 8 | 0.263<br>(0.304) | 0.676***<br>(0.223) | 1.698<br>(1.737) |
| day 9 | 0.122<br>(0.294) | 0.518**<br>(0.224) | 0.849<br>(1.543) |
| day 10 | 0.198<br>(0.256) | 0.603**<br>(0.250) | 1.341<br>(1.515) |
| day 11 | 0.267 | 0.600** | 1.555 |
|  | (0.238) | (0.261) | (1.635) |
| day 12 | 0.144<br>(0.247) | 0.754**<br>(0.313) | 0.541<br>(1.632) |
| day 13 | 0.154<br>(0.223) | 0.630**<br>(0.287) | 0.370<br>(1.608) |
| day 14 | 0.178<br>(0.222) | 0.614*<br>(0.310) | 0.543<br>(1.672) |
| day 15 | 0.174<br>(0.213) | 0.524*<br>(0.293) | 0.331<br>(1.503) |
| day 16 | 0.208<br>(0.211) | 0.546*<br>(0.263) | 0.179<br>(1.484) |
| day 17 | 0.307<br>(0.223) | 0.577*<br>(0.271) | 0.434<br>(1.413) |
| day 18 | 0.188<br>(0.187) | 0.437<br>(0.269) | −0.276<br>(1.134) |
| day 19 | 0.182<br>(0.253) | 0.175<br>(0.261) | −0.496<br>(1.290) |
| day 20 | 0.209<br>(0.218) | 0.306<br>(0.273) | −0.291<br>(1.269) |
| day 21 | 0.166<br>(0.218) | 0.325<br>(0.273) | −0.804<br>(1.076) |
| day 22 | 0.189<br>(0.218) | 0.284<br>(0.281) | −0.297<br>(1.272) |
| day 23 | 0.114<br>(0.181) | 0.255<br>(0.263) | −0.703<br>(1.153) |
| day 24 | 0.100<br>(0.208) | 0.196<br>(0.262) | −0.482<br>(1.194) |
| day 25 | 0.116<br>(0.193) | 0.241<br>(0.236) | 0.124<br>(1.207) |
| day 26 | −0.012<br>(0.143) | 0.081<br>(0.182) | −0.312<br>(1.047) |
| day 27 | −0.069<br>(0.142) | 0.103<br>(0.188) | −0.328<br>(0.829) |
| day 28 | −0.120<br>(0.173) | 0.025<br>(0.192) | −0.311<br>(0.844) |
| day 29 | −0.032<br>(0.131) | 0.096<br>(0.164) | 0.338<br>(0.940) |
| day 30 | −0.100<br>(0.106) | 0.006<br>(0.148) | 0.210<br>(0.930) |
| day 31 | −0.051<br>(0.125) | 0.049<br>(0.154) | 0.394<br>(0.835) |
| day 32 | −0.034<br>(0.120) | 0.047<br>(0.148) | 0.379<br>(0.774) |
| day 33 | −0.161<br>(0.138) | 0.050<br>(0.148) | −0.270<br>(0.705) |
| day 34 | −0.120<br>(0.116) | 0.154<br>(0.162) | −0.176<br>(0.730) |
| day 35 | −0.129<br>(0.143) | 0.043<br>(0.122) | −0.488<br>(0.628) |
| day 36 | −0.049<br>(0.150) | 0.104<br>(0.110) | −0.329<br>(0.737) |
| day 37 | −0.137<br>(0.122) | 0.024<br>(0.112) | −0.676<br>(0.719) |
| day 38 | −0.205*<br>(0.108) | −0.007<br>(0.104) | −0.314<br>(0.600) |
| day 39 | −0.132<br>(0.093) | 0.060<br>(0.111) | −0.319<br>(0.579) |
| day 40 | −0.140<br>(0.091) | 0.043<br>(0.119) | −0.574<br>(0.640) |
| Constant | 30.186***<br>(8.618) | 25.208***<br>(7.743) | 220.637***<br>(47.945) |
| Observations | 3,164 | 3,164 | 3,164 |
| R<sup>2</sup> | 0.619 | 0.660 | 0.774 |
| Adjusted R<sup>2</sup> | 0.609 | 0.651 | 0.767 |
| Residual Std. Error (df = 3078) | 0.624 | 0.589 | 3.065 |

*Note:*
\*p<0.1; \*\*p<0.05; \*\*\*p<0.01

**Table S10:**
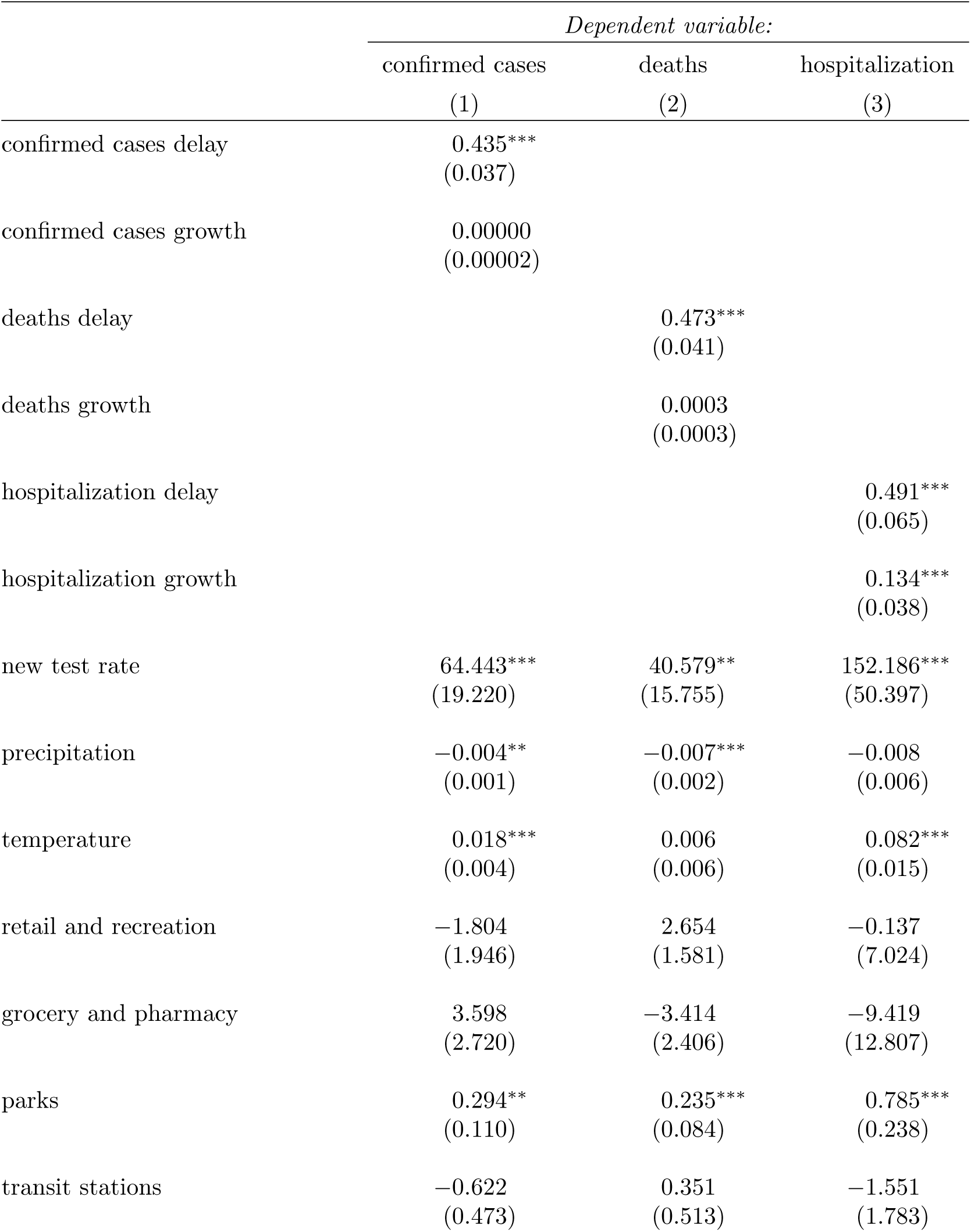
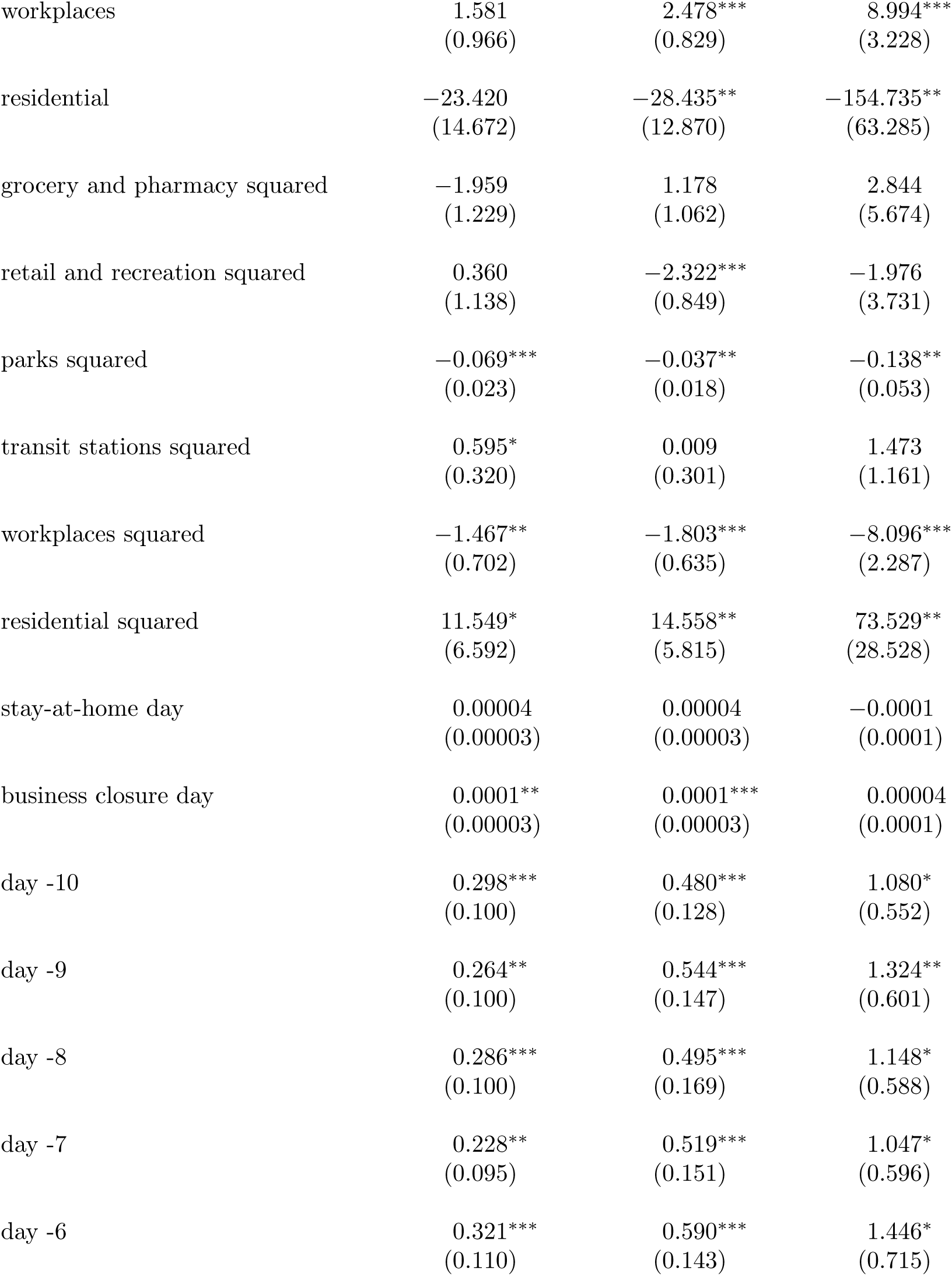
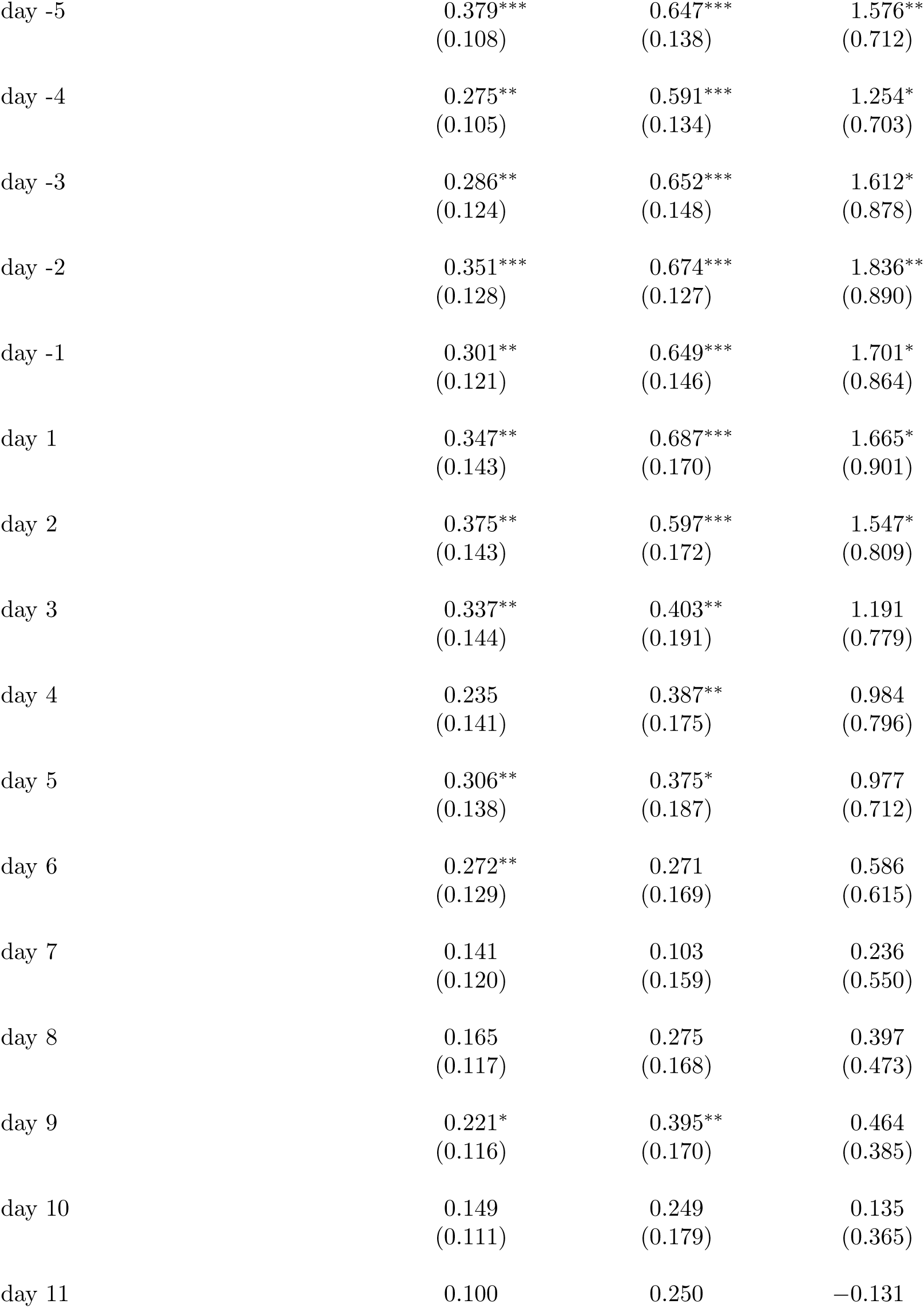
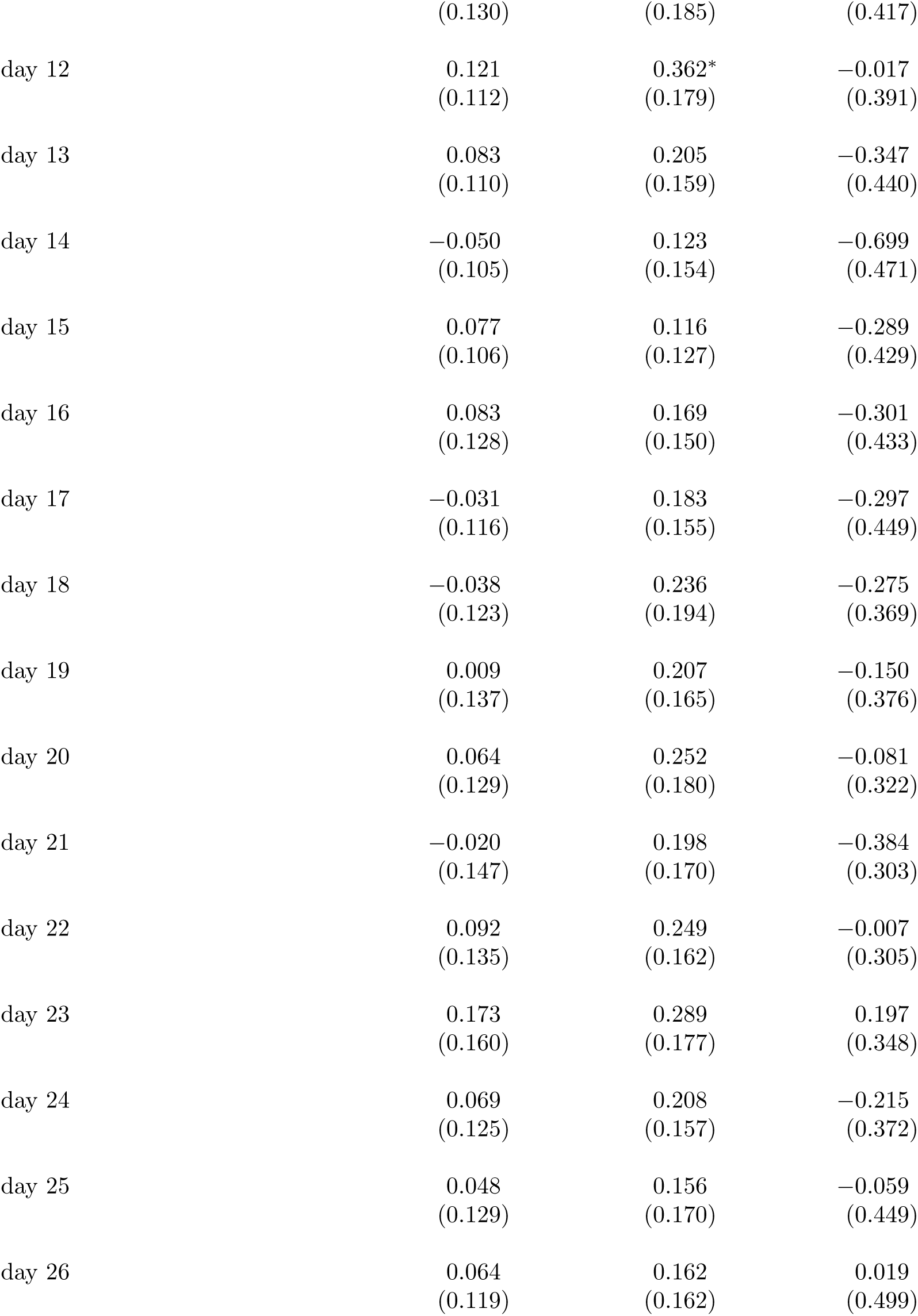
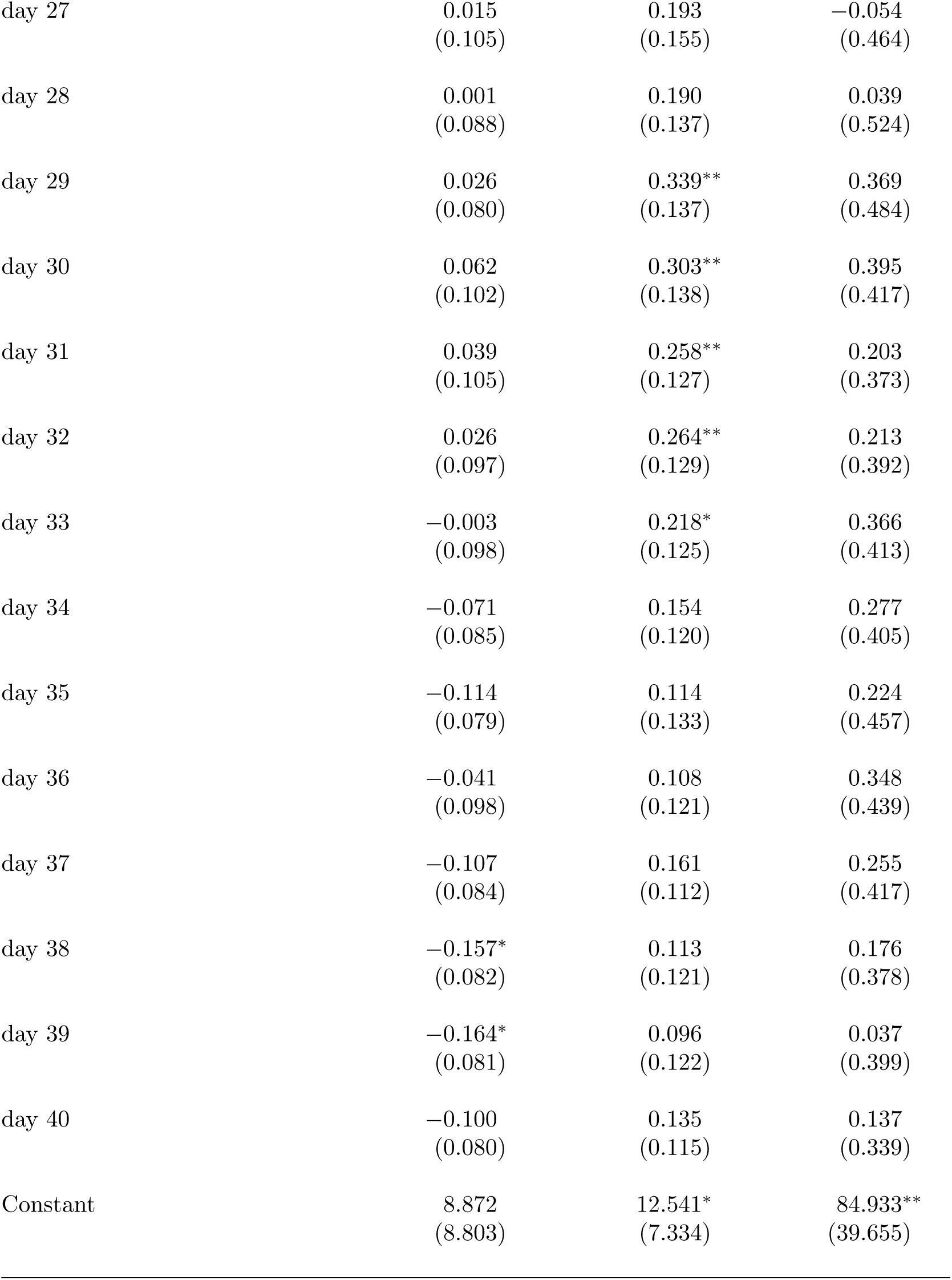
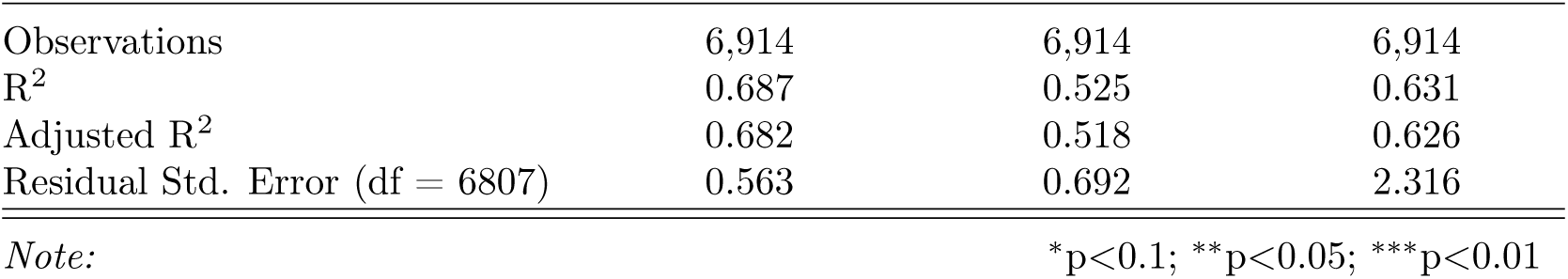
Event Study regression summary statistics of fig. 3 (for later states only, i.e. all states except NY, NJ, MA, CT, MI, DC, RI, IL, WA, PA, GA, VT, MD, FL, LA).

|  | *Dependent variable:* |  |  |
| --- | --- | --- | --- |
|  | confirmed cases | deaths | hospitalization |
|  | (1) | (2) | (3) |
| confirmed cases delay | 0.435***<br>(0.037) |  |  |
| confirmed cases growth | 0.00000<br>(0.00002) |  |  |
| deaths delay |  | 0.473***<br>(0.041) |  |
| deaths growth |  | 0.0003<br>(0.0003) |  |
| hospitalization delay |  |  | 0.491***<br>(0.065) |
| hospitalization growth |  |  | 0.134***<br>(0.038) |
| new test rate | 64.443***<br>(19.220) | 40.579**<br>(15.755) | 152.186***<br>(50.397) |
| precipitation | -0.004**<br>(0.001) | -0.007***<br>(0.002) | -0.008<br>(0.006) |
| temperature | 0.018***<br>(0.004) | 0.006<br>(0.006) | 0.082***<br>(0.015) |
| retail and recreation | -1.804<br>(1.946) | 2.654<br>(1.581) | -0.137<br>(7.024) |
| grocery and pharmacy | 3.598<br>(2.720) | -3.414<br>(2.406) | -9.419<br>(12.807) |
| parks | 0.294**<br>(0.110) | 0.235***<br>(0.084) | 0.785***<br>(0.238) |
| transit stations | -0.622<br>(0.473) | 0.351<br>(0.513) | -1.551<br>(1.783) |
| workplaces | 1.581<br>(0.966) | 2.478***<br>(0.829) | 8.994***<br>(3.228) |
| residential | -23.420<br>(14.672) | -28.435**<br>(12.870) | -154.735**<br>(63.285) |
| grocery and pharmacy squared | -1.959<br>(1.229) | 1.178<br>(1.062) | 2.844<br>(5.674) |
| retail and recreation squared | 0.360<br>(1.138) | -2.322***<br>(0.849) | -1.976<br>(3.731) |
| parks squared | -0.069***<br>(0.023) | -0.037**<br>(0.018) | -0.138**<br>(0.053) |
| transit stations squared | 0.595*<br>(0.320) | 0.009<br>(0.301) | 1.473<br>(1.161) |
| workplaces squared | -1.467**<br>(0.702) | -1.803***<br>(0.635) | -8.096***<br>(2.287) |
| residential squared | 11.549*<br>(6.592) | 14.558**<br>(5.815) | 73.529**<br>(28.528) |
| stay-at-home day | 0.00004<br>(0.00003) | 0.00004<br>(0.00003) | -0.0001<br>(0.0001) |
| business closure day | 0.0001**<br>(0.00003) | 0.0001***<br>(0.00003) | 0.00004<br>(0.0001) |
| day -10 | 0.298***<br>(0.100) | 0.480***<br>(0.128) | 1.080*<br>(0.552) |
| day -9 | 0.264**<br>(0.100) | 0.544***<br>(0.147) | 1.324**<br>(0.601) |
| day -8 | 0.286***<br>(0.100) | 0.495***<br>(0.169) | 1.148*<br>(0.588) |
| day -7 | 0.228**<br>(0.095) | 0.519***<br>(0.151) | 1.047*<br>(0.596) |
| day -6 | 0.321***<br>(0.110) | 0.590***<br>(0.143) | 1.446*<br>(0.715) |
| day -5 | 0.379***<br>(0.108) | 0.647***<br>(0.138) | 1.576**<br>(0.712) |
| day -4 | 0.275**<br>(0.105) | 0.591***<br>(0.134) | 1.254*<br>(0.703) |
| day -3 | 0.286**<br>(0.124) | 0.652***<br>(0.148) | 1.612*<br>(0.878) |
| day -2 | 0.351***<br>(0.128) | 0.674***<br>(0.127) | 1.836**<br>(0.890) |
| day -1 | 0.301**<br>(0.121) | 0.649***<br>(0.146) | 1.701*<br>(0.864) |
| day 1 | 0.347**<br>(0.143) | 0.687***<br>(0.170) | 1.665*<br>(0.901) |
| day 2 | 0.375**<br>(0.143) | 0.597***<br>(0.172) | 1.547*<br>(0.809) |
| day 3 | 0.337**<br>(0.144) | 0.403**<br>(0.191) | 1.191<br>(0.779) |
| day 4 | 0.235<br>(0.141) | 0.387**<br>(0.175) | 0.984<br>(0.796) |
| day 5 | 0.306**<br>(0.138) | 0.375*<br>(0.187) | 0.977<br>(0.712) |
| day 6 | 0.272**<br>(0.129) | 0.271<br>(0.169) | 0.586<br>(0.615) |
| day 7 | 0.141<br>(0.120) | 0.103<br>(0.159) | 0.236<br>(0.550) |
| day 8 | 0.165<br>(0.117) | 0.275<br>(0.168) | 0.397<br>(0.473) |
| day 9 | 0.221*<br>(0.116) | 0.395**<br>(0.170) | 0.464<br>(0.385) |
| day 10 | 0.149<br>(0.111) | 0.249<br>(0.179) | 0.135<br>(0.365) |
| day 11 | 0.100 | 0.250 | −0.131 |
|  | (0.130) | (0.185) | (0.417) |
| day 12 | 0.121<br>(0.112) | 0.362*<br>(0.179) | −0.017<br>(0.391) |
| day 13 | 0.083<br>(0.110) | 0.205<br>(0.159) | −0.347<br>(0.440) |
| day 14 | −0.050<br>(0.105) | 0.123<br>(0.154) | −0.699<br>(0.471) |
| day 15 | 0.077<br>(0.106) | 0.116<br>(0.127) | −0.289<br>(0.429) |
| day 16 | 0.083<br>(0.128) | 0.169<br>(0.150) | −0.301<br>(0.433) |
| day 17 | −0.031<br>(0.116) | 0.183<br>(0.155) | −0.297<br>(0.449) |
| day 18 | −0.038<br>(0.123) | 0.236<br>(0.194) | −0.275<br>(0.369) |
| day 19 | 0.009<br>(0.137) | 0.207<br>(0.165) | −0.150<br>(0.376) |
| day 20 | 0.064<br>(0.129) | 0.252<br>(0.180) | −0.081<br>(0.322) |
| day 21 | −0.020<br>(0.147) | 0.198<br>(0.170) | −0.384<br>(0.303) |
| day 22 | 0.092<br>(0.135) | 0.249<br>(0.162) | −0.007<br>(0.305) |
| day 23 | 0.173<br>(0.160) | 0.289<br>(0.177) | 0.197<br>(0.348) |
| day 24 | 0.069<br>(0.125) | 0.208<br>(0.157) | −0.215<br>(0.372) |
| day 25 | 0.048<br>(0.129) | 0.156<br>(0.170) | −0.059<br>(0.449) |
| day 26 | 0.064<br>(0.119) | 0.162<br>(0.162) | 0.019<br>(0.499) |
| day 27 | 0.015<br>(0.105) | 0.193<br>(0.155) | −0.054<br>(0.464) |
| day 28 | 0.001<br>(0.088) | 0.190<br>(0.137) | 0.039<br>(0.524) |
| day 29 | 0.026<br>(0.080) | 0.339**<br>(0.137) | 0.369<br>(0.484) |
| day 30 | 0.062<br>(0.102) | 0.303**<br>(0.138) | 0.395<br>(0.417) |
| day 31 | 0.039<br>(0.105) | 0.258**<br>(0.127) | 0.203<br>(0.373) |
| day 32 | 0.026<br>(0.097) | 0.264**<br>(0.129) | 0.213<br>(0.392) |
| day 33 | −0.003<br>(0.098) | 0.218*<br>(0.125) | 0.366<br>(0.413) |
| day 34 | −0.071<br>(0.085) | 0.154<br>(0.120) | 0.277<br>(0.405) |
| day 35 | −0.114<br>(0.079) | 0.114<br>(0.133) | 0.224<br>(0.457) |
| day 36 | −0.041<br>(0.098) | 0.108<br>(0.121) | 0.348<br>(0.439) |
| day 37 | −0.107<br>(0.084) | 0.161<br>(0.112) | 0.255<br>(0.417) |
| day 38 | −0.157*<br>(0.082) | 0.113<br>(0.121) | 0.176<br>(0.378) |
| day 39 | −0.164*<br>(0.081) | 0.096<br>(0.122) | 0.037<br>(0.399) |
| day 40 | −0.100<br>(0.080) | 0.135<br>(0.115) | 0.137<br>(0.339) |
| Constant | 8.872<br>(8.803) | 12.541*<br>(7.334) | 84.933**<br>(39.655) |
| Observations | 6,914 | 6,914 | 6,914 |
| R<sup>2</sup> | 0.687 | 0.525 | 0.631 |
| Adjusted R<sup>2</sup> | 0.682 | 0.518 | 0.626 |
| Residual Std. Error (df = 6807) | 0.563 | 0.692 | 2.316 |
| *Note:* |  | *p<0.1; **p<0.05; ***p<0.01 |  |

**Table S11:**
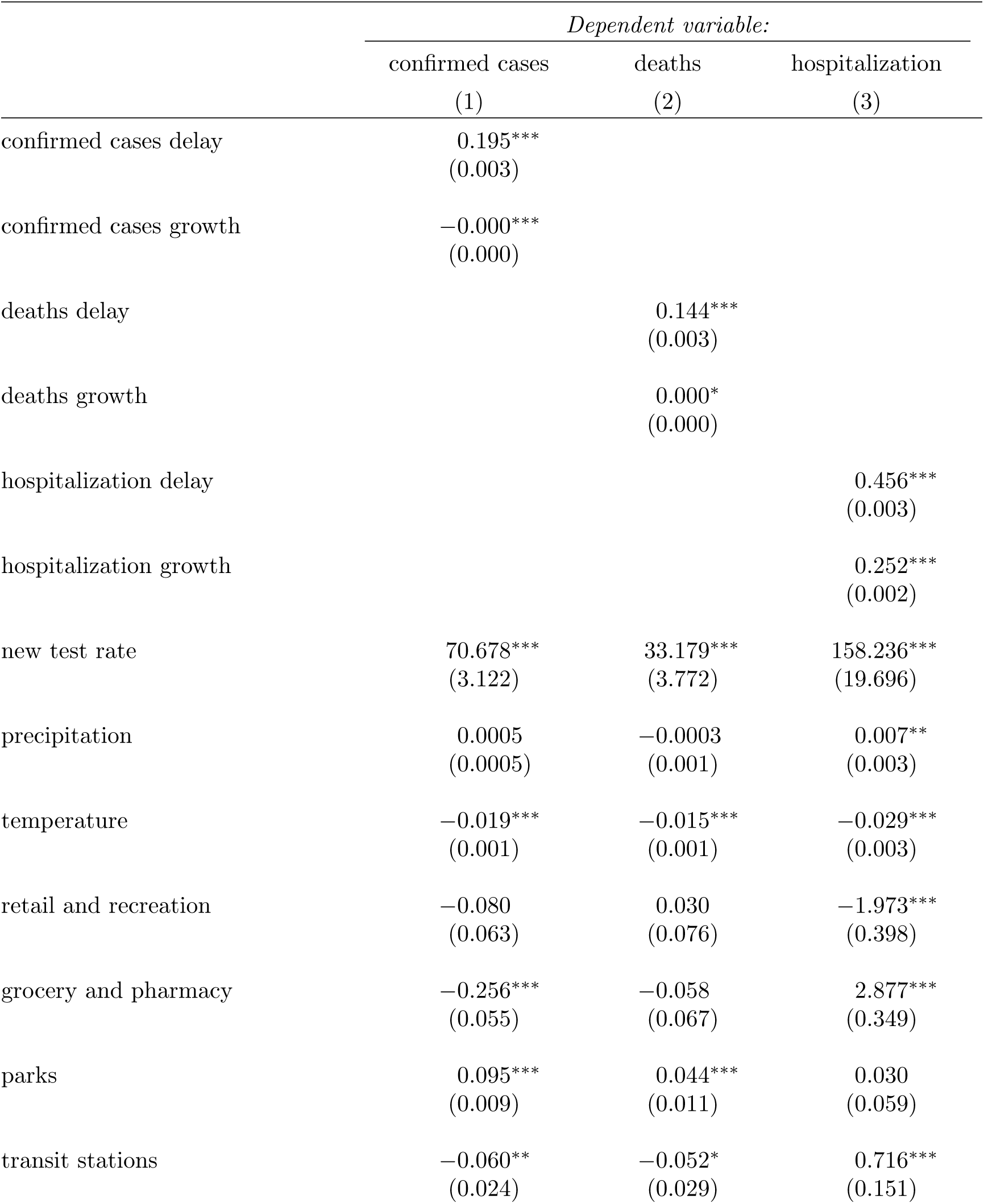
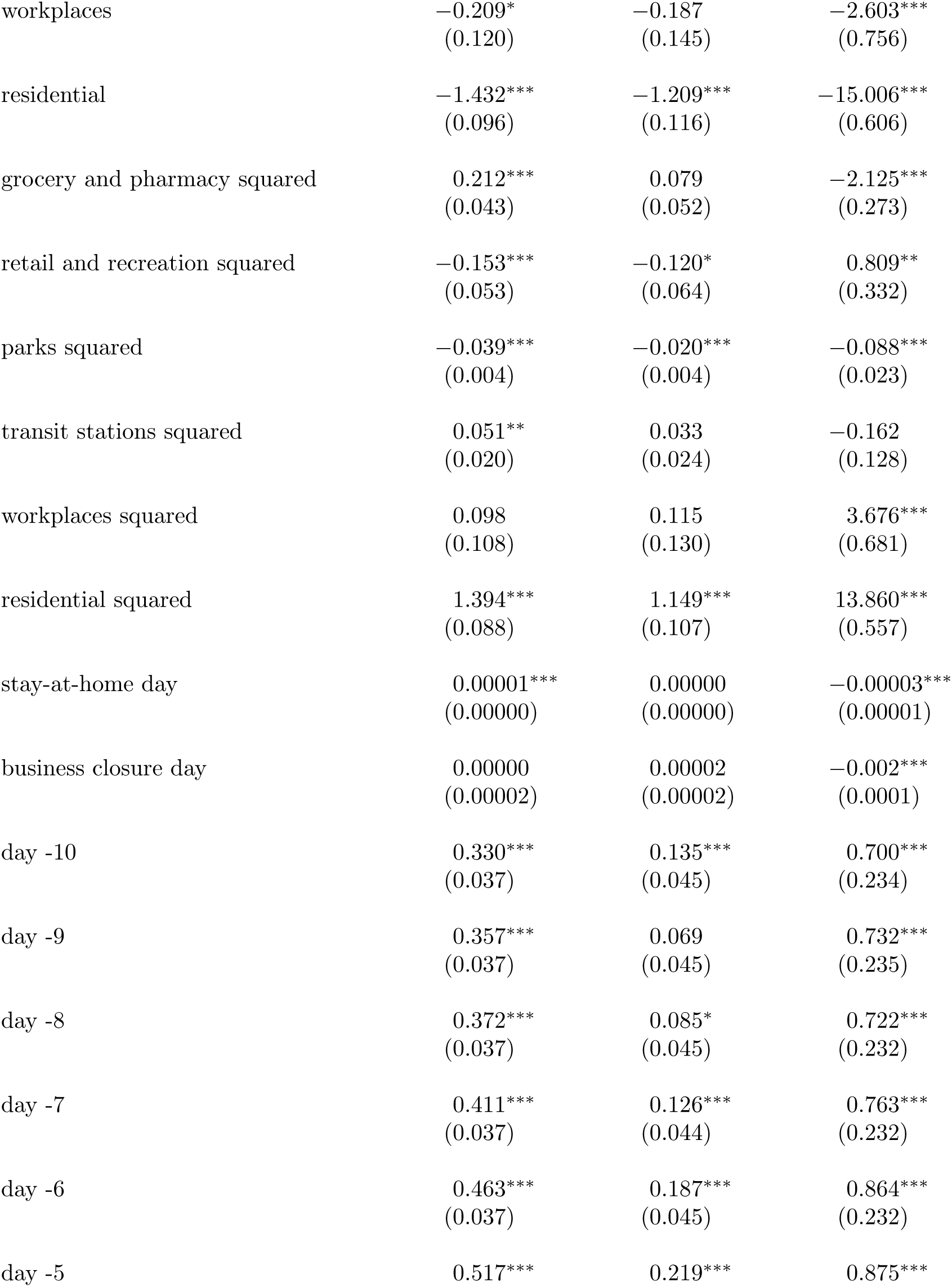
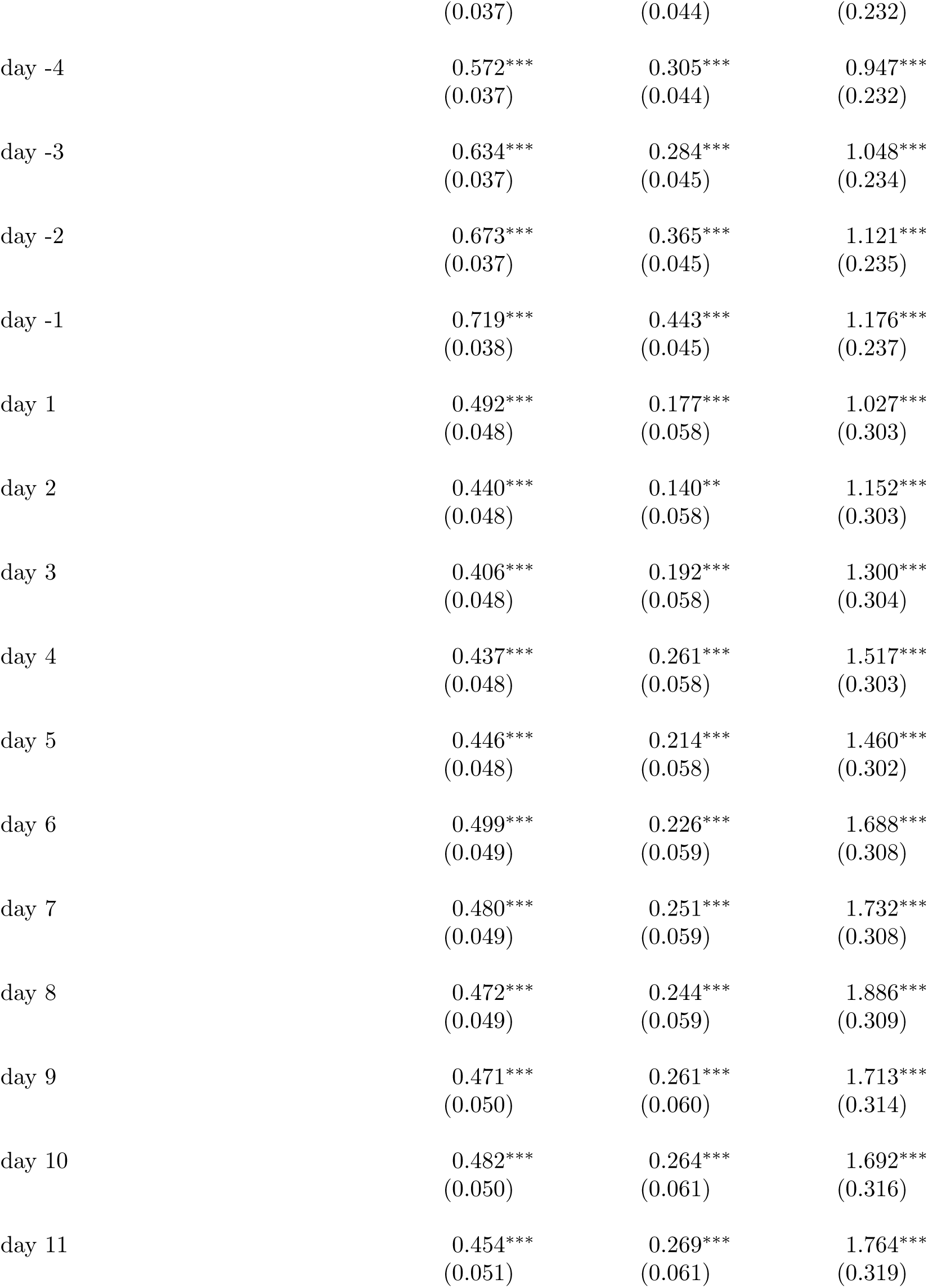
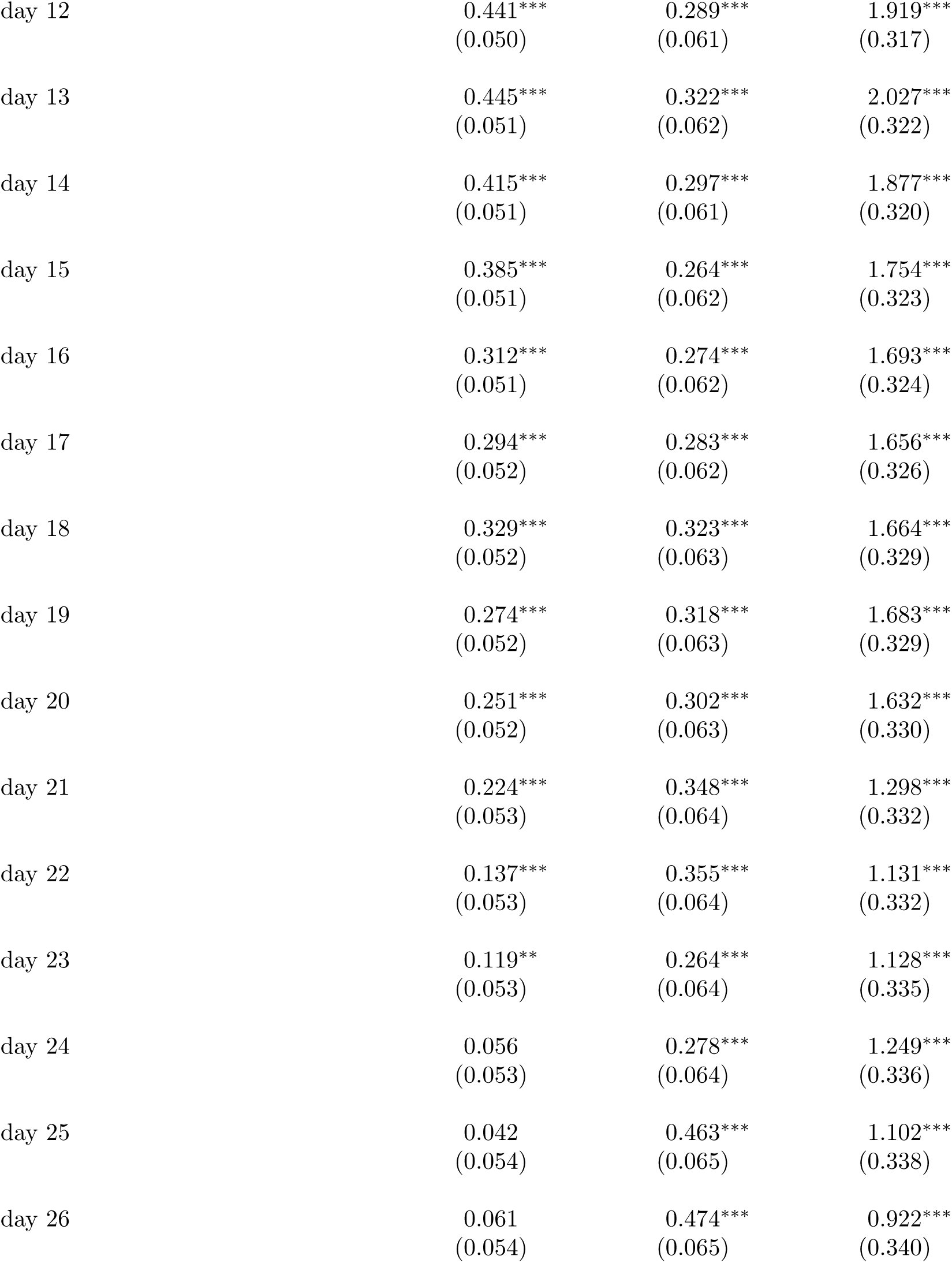
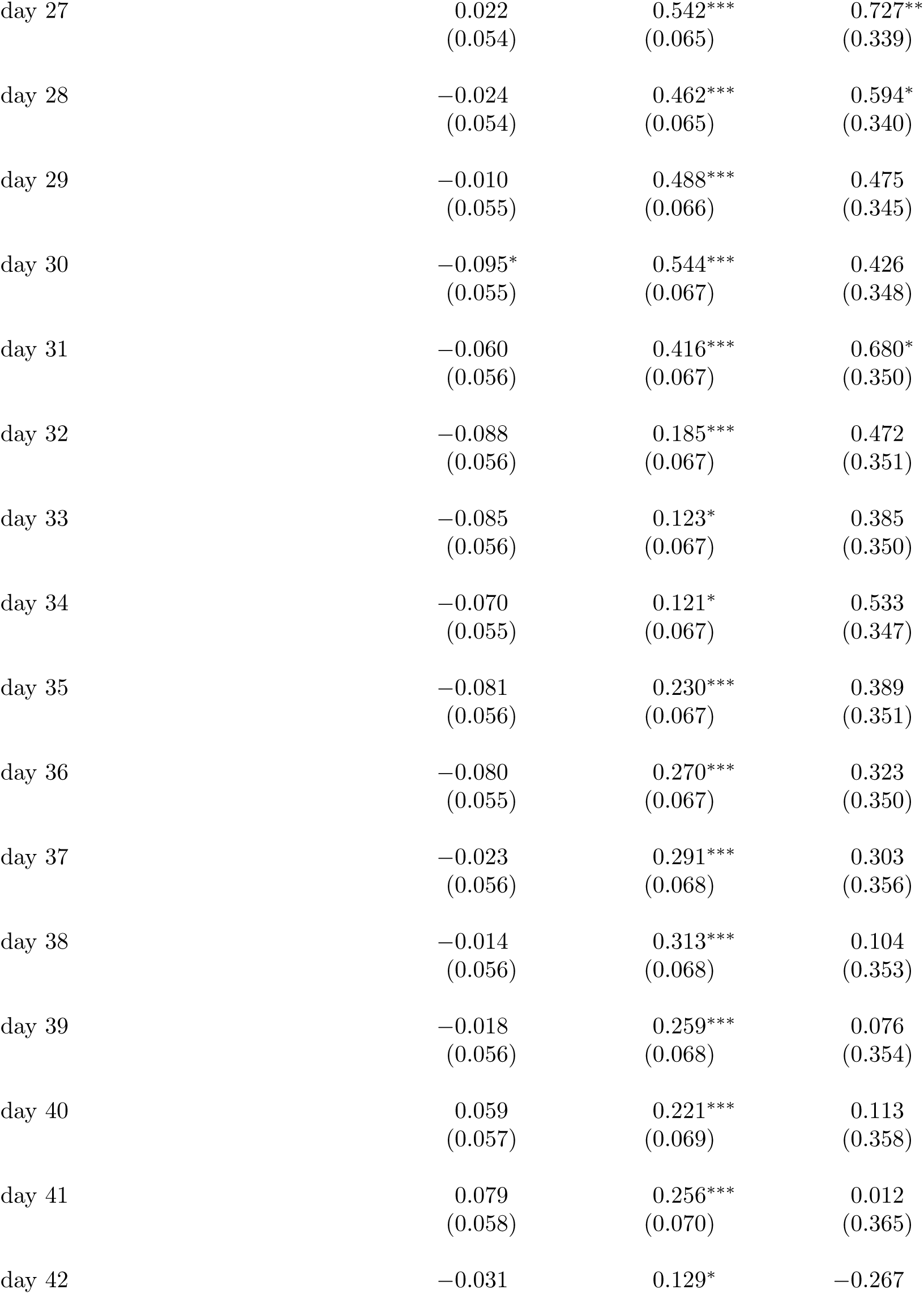
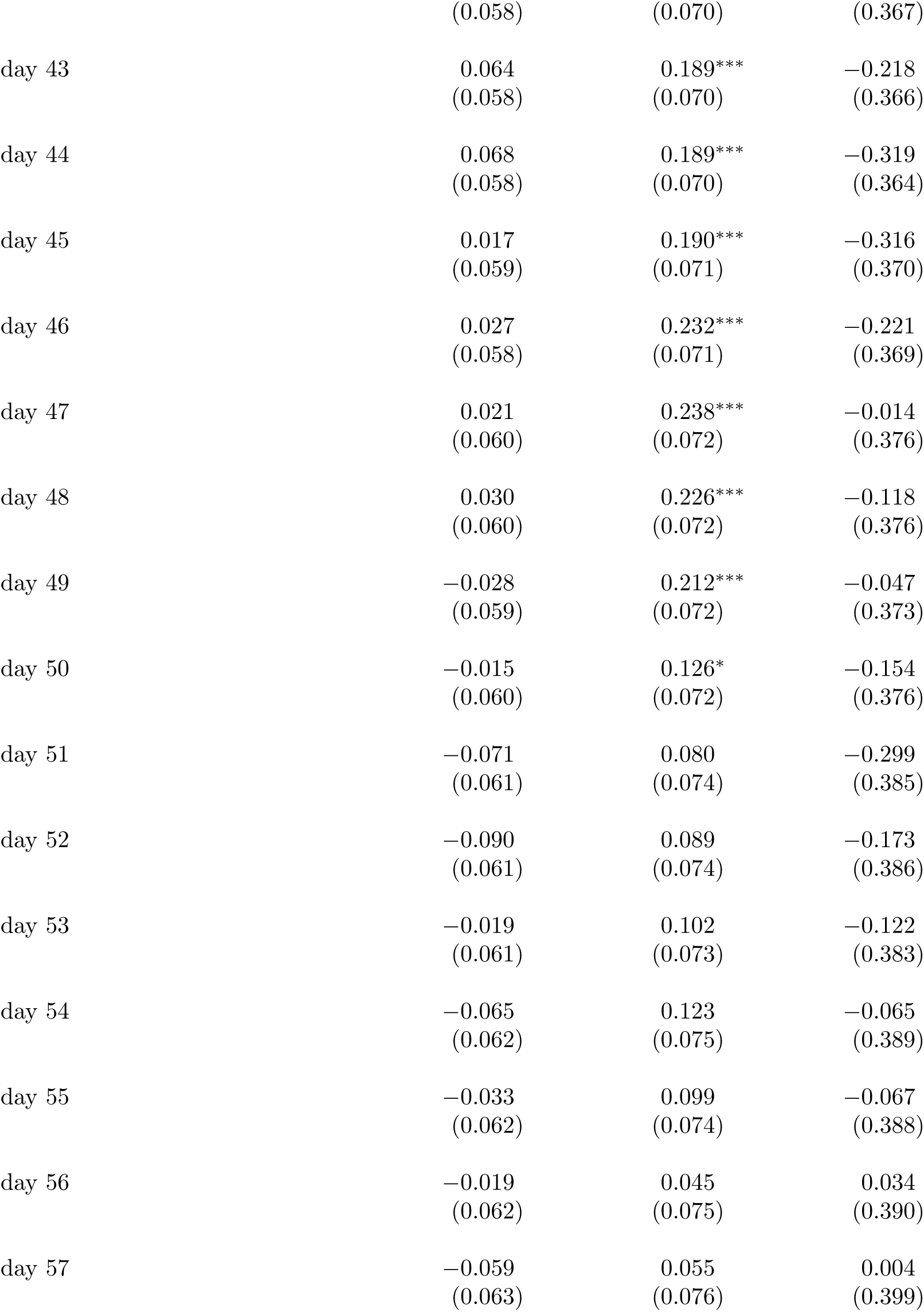
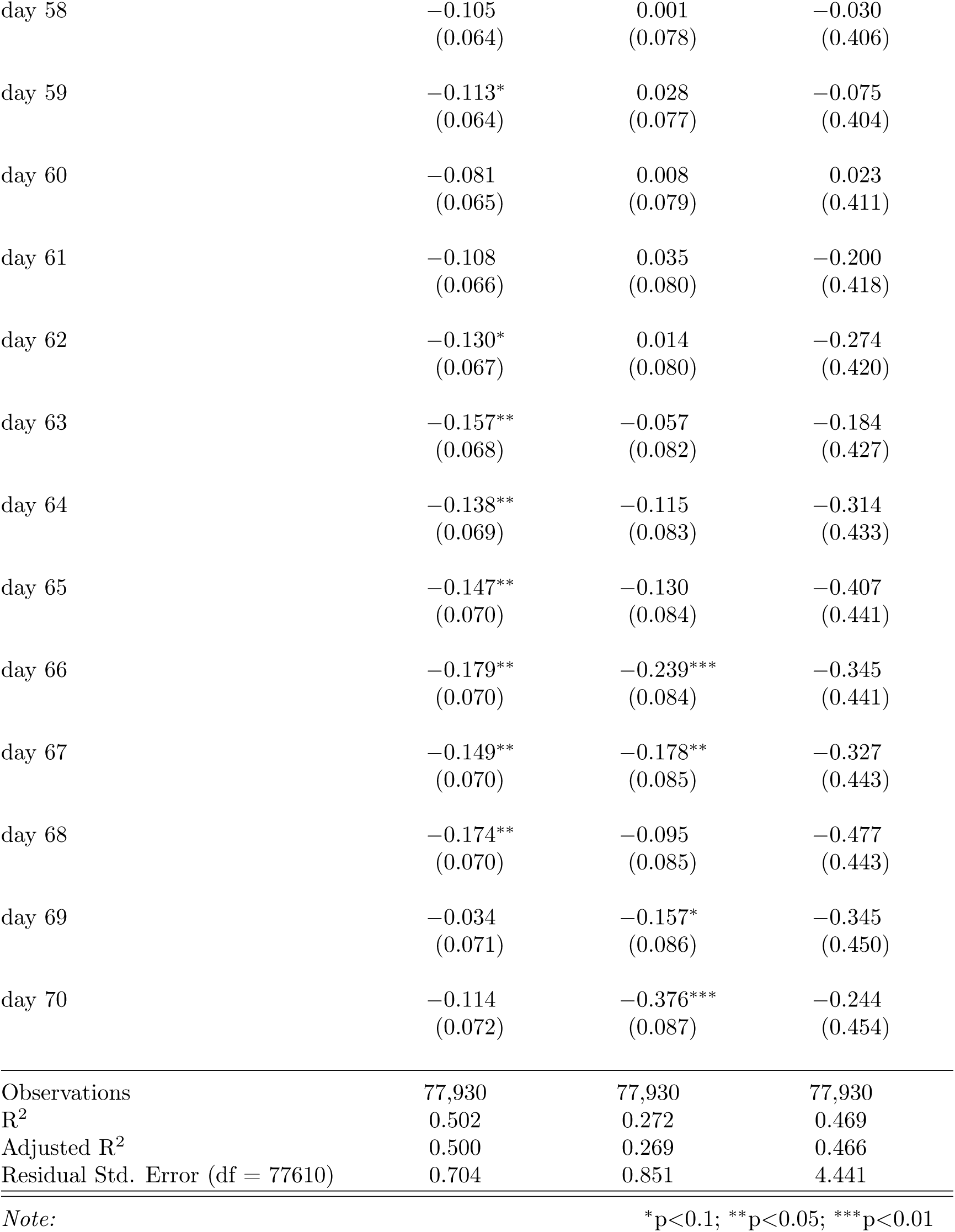
Event Study regression summary statistics for robustness check (fig. S4) where we focus on county-level mandates only.

|  | *Dependent variable:* |  |  |
| --- | --- | --- | --- |
|  | confirmed cases | deaths | hospitalization |
|  | (1) | (2) | (3) |
| confirmed cases delay | 0.195***<br>(0.003) |  |  |
| confirmed cases growth | -0.000***<br>(0.000) |  |  |
| deaths delay |  | 0.144***<br>(0.003) |  |
| deaths growth |  | 0.000*<br>(0.000) |  |
| hospitalization delay |  |  | 0.456***<br>(0.003) |
| hospitalization growth |  |  | 0.252***<br>(0.002) |
| new test rate | 70.678***<br>(3.122) | 33.179***<br>(3.772) | 158.236***<br>(19.696) |
| precipitation | 0.0005<br>(0.0005) | -0.0003<br>(0.001) | 0.007**<br>(0.003) |
| temperature | -0.019***<br>(0.001) | -0.015***<br>(0.001) | -0.029***<br>(0.003) |
| retail and recreation | -0.080<br>(0.063) | 0.030<br>(0.076) | -1.973***<br>(0.398) |
| grocery and pharmacy | -0.256***<br>(0.055) | -0.058<br>(0.067) | 2.877***<br>(0.349) |
| parks | 0.095***<br>(0.009) | 0.044***<br>(0.011) | 0.030<br>(0.059) |
| transit stations | -0.060**<br>(0.024) | -0.052*<br>(0.029) | 0.716***<br>(0.151) |
| workplaces | −0.209*<br>(0.120) | −0.187<br>(0.145) | −2.603***<br>(0.756) |
| residential | −1.432***<br>(0.096) | −1.209***<br>(0.116) | −15.006***<br>(0.606) |
| grocery and pharmacy squared | 0.212***<br>(0.043) | 0.079<br>(0.052) | −2.125***<br>(0.273) |
| retail and recreation squared | −0.153***<br>(0.053) | −0.120*<br>(0.064) | 0.809**<br>(0.332) |
| parks squared | −0.039***<br>(0.004) | −0.020***<br>(0.004) | −0.088***<br>(0.023) |
| transit stations squared | 0.051**<br>(0.020) | 0.033<br>(0.024) | −0.162<br>(0.128) |
| workplaces squared | 0.098<br>(0.108) | 0.115<br>(0.130) | 3.676***<br>(0.681) |
| residential squared | 1.394***<br>(0.088) | 1.149***<br>(0.107) | 13.860***<br>(0.557) |
| stay-at-home day | 0.00001***<br>(0.00000) | 0.00000<br>(0.00000) | −0.00003***<br>(0.00001) |
| business closure day | 0.00000<br>(0.00002) | 0.00002<br>(0.00002) | −0.002***<br>(0.0001) |
| day -10 | 0.330***<br>(0.037) | 0.135***<br>(0.045) | 0.700***<br>(0.234) |
| day -9 | 0.357***<br>(0.037) | 0.069<br>(0.045) | 0.732***<br>(0.235) |
| day -8 | 0.372***<br>(0.037) | 0.085*<br>(0.045) | 0.722***<br>(0.232) |
| day -7 | 0.411***<br>(0.037) | 0.126***<br>(0.044) | 0.763***<br>(0.232) |
| day -6 | 0.463***<br>(0.037) | 0.187***<br>(0.045) | 0.864***<br>(0.232) |
| day -5 | 0.517*** | 0.219*** | 0.875*** |

Working Paper
|  | (0.037) | (0.044) | (0.232) |
| --- | --- | --- | --- |
| day -4 | 0.572***<br>(0.037) | 0.305***<br>(0.044) | 0.947***<br>(0.232) |
| day -3 | 0.634***<br>(0.037) | 0.284***<br>(0.045) | 1.048***<br>(0.234) |
| day -2 | 0.673***<br>(0.037) | 0.365***<br>(0.045) | 1.121***<br>(0.235) |
| day -1 | 0.719***<br>(0.038) | 0.443***<br>(0.045) | 1.176***<br>(0.237) |
| day 1 | 0.492***<br>(0.048) | 0.177***<br>(0.058) | 1.027***<br>(0.303) |
| day 2 | 0.440***<br>(0.048) | 0.140**<br>(0.058) | 1.152***<br>(0.303) |
| day 3 | 0.406***<br>(0.048) | 0.192***<br>(0.058) | 1.300***<br>(0.304) |
| day 4 | 0.437***<br>(0.048) | 0.261***<br>(0.058) | 1.517***<br>(0.303) |
| day 5 | 0.446***<br>(0.048) | 0.214***<br>(0.058) | 1.460***<br>(0.302) |
| day 6 | 0.499***<br>(0.049) | 0.226***<br>(0.059) | 1.688***<br>(0.308) |
| day 7 | 0.480***<br>(0.049) | 0.251***<br>(0.059) | 1.732***<br>(0.308) |
| day 8 | 0.472***<br>(0.049) | 0.244***<br>(0.059) | 1.886***<br>(0.309) |
| day 9 | 0.471***<br>(0.050) | 0.261***<br>(0.060) | 1.713***<br>(0.314) |
| day 10 | 0.482***<br>(0.050) | 0.264***<br>(0.061) | 1.692***<br>(0.316) |
| day 11 | 0.454***<br>(0.051) | 0.269***<br>(0.061) | 1.764***<br>(0.319) |
| day 12 | 0.441***<br>(0.050) | 0.289***<br>(0.061) | 1.919***<br>(0.317) |
| day 13 | 0.445***<br>(0.051) | 0.322***<br>(0.062) | 2.027***<br>(0.322) |
| day 14 | 0.415***<br>(0.051) | 0.297***<br>(0.061) | 1.877***<br>(0.320) |
| day 15 | 0.385***<br>(0.051) | 0.264***<br>(0.062) | 1.754***<br>(0.323) |
| day 16 | 0.312***<br>(0.051) | 0.274***<br>(0.062) | 1.693***<br>(0.324) |
| day 17 | 0.294***<br>(0.052) | 0.283***<br>(0.062) | 1.656***<br>(0.326) |
| day 18 | 0.329***<br>(0.052) | 0.323***<br>(0.063) | 1.664***<br>(0.329) |
| day 19 | 0.274***<br>(0.052) | 0.318***<br>(0.063) | 1.683***<br>(0.329) |
| day 20 | 0.251***<br>(0.052) | 0.302***<br>(0.063) | 1.632***<br>(0.330) |
| day 21 | 0.224***<br>(0.053) | 0.348***<br>(0.064) | 1.298***<br>(0.332) |
| day 22 | 0.137***<br>(0.053) | 0.355***<br>(0.064) | 1.131***<br>(0.332) |
| day 23 | 0.119**<br>(0.053) | 0.264***<br>(0.064) | 1.128***<br>(0.335) |
| day 24 | 0.056<br>(0.053) | 0.278***<br>(0.064) | 1.249***<br>(0.336) |
| day 25 | 0.042<br>(0.054) | 0.463***<br>(0.065) | 1.102***<br>(0.338) |
| day 26 | 0.061<br>(0.054) | 0.474***<br>(0.065) | 0.922***<br>(0.340) |
| day 27 | 0.022<br>(0.054) | 0.542***<br>(0.065) | 0.727**<br>(0.339) |
| day 28 | −0.024<br>(0.054) | 0.462***<br>(0.065) | 0.594*<br>(0.340) |
| day 29 | −0.010<br>(0.055) | 0.488***<br>(0.066) | 0.475<br>(0.345) |
| day 30 | −0.095*<br>(0.055) | 0.544***<br>(0.067) | 0.426<br>(0.348) |
| day 31 | −0.060<br>(0.056) | 0.416***<br>(0.067) | 0.680*<br>(0.350) |
| day 32 | −0.088<br>(0.056) | 0.185***<br>(0.067) | 0.472<br>(0.351) |
| day 33 | −0.085<br>(0.056) | 0.123*<br>(0.067) | 0.385<br>(0.350) |
| day 34 | −0.070<br>(0.055) | 0.121*<br>(0.067) | 0.533<br>(0.347) |
| day 35 | −0.081<br>(0.056) | 0.230***<br>(0.067) | 0.389<br>(0.351) |
| day 36 | −0.080<br>(0.055) | 0.270***<br>(0.067) | 0.323<br>(0.350) |
| day 37 | −0.023<br>(0.056) | 0.291***<br>(0.068) | 0.303<br>(0.356) |
| day 38 | −0.014<br>(0.056) | 0.313***<br>(0.068) | 0.104<br>(0.353) |
| day 39 | −0.018<br>(0.056) | 0.259***<br>(0.068) | 0.076<br>(0.354) |
| day 40 | 0.059<br>(0.057) | 0.221***<br>(0.069) | 0.113<br>(0.358) |
| day 41 | 0.079<br>(0.058) | 0.256***<br>(0.070) | 0.012<br>(0.365) |
| day 42 | −0.031 | 0.129* | −0.267 |

Working Paper
|  | (0.058) | (0.070) | (0.367) |
| --- | --- | --- | --- |
| day 43 | 0.064<br>(0.058) | 0.189***<br>(0.070) | -0.218<br>(0.366) |
| day 44 | 0.068<br>(0.058) | 0.189***<br>(0.070) | -0.319<br>(0.364) |
| day 45 | 0.017<br>(0.059) | 0.190***<br>(0.071) | -0.316<br>(0.370) |
| day 46 | 0.027<br>(0.058) | 0.232***<br>(0.071) | -0.221<br>(0.369) |
| day 47 | 0.021<br>(0.060) | 0.238***<br>(0.072) | -0.014<br>(0.376) |
| day 48 | 0.030<br>(0.060) | 0.226***<br>(0.072) | -0.118<br>(0.376) |
| day 49 | -0.028<br>(0.059) | 0.212***<br>(0.072) | -0.047<br>(0.373) |
| day 50 | -0.015<br>(0.060) | 0.126*<br>(0.072) | -0.154<br>(0.376) |
| day 51 | -0.071<br>(0.061) | 0.080<br>(0.074) | -0.299<br>(0.385) |
| day 52 | -0.090<br>(0.061) | 0.089<br>(0.074) | -0.173<br>(0.386) |
| day 53 | -0.019<br>(0.061) | 0.102<br>(0.073) | -0.122<br>(0.383) |
| day 54 | -0.065<br>(0.062) | 0.123<br>(0.075) | -0.065<br>(0.389) |
| day 55 | -0.033<br>(0.062) | 0.099<br>(0.074) | -0.067<br>(0.388) |
| day 56 | -0.019<br>(0.062) | 0.045<br>(0.075) | 0.034<br>(0.390) |
| day 57 | -0.059<br>(0.063) | 0.055<br>(0.076) | 0.004<br>(0.399) |
| day 58 | −0.105<br>(0.064) | 0.001<br>(0.078) | −0.030<br>(0.406) |
| day 59 | −0.113*<br>(0.064) | 0.028<br>(0.077) | −0.075<br>(0.404) |
| day 60 | −0.081<br>(0.065) | 0.008<br>(0.079) | 0.023<br>(0.411) |
| day 61 | −0.108<br>(0.066) | 0.035<br>(0.080) | −0.200<br>(0.418) |
| day 62 | −0.130*<br>(0.067) | 0.014<br>(0.080) | −0.274<br>(0.420) |
| day 63 | −0.157**<br>(0.068) | −0.057<br>(0.082) | −0.184<br>(0.427) |
| day 64 | −0.138**<br>(0.069) | −0.115<br>(0.083) | −0.314<br>(0.433) |
| day 65 | −0.147**<br>(0.070) | −0.130<br>(0.084) | −0.407<br>(0.441) |
| day 66 | −0.179**<br>(0.070) | −0.239***<br>(0.084) | −0.345<br>(0.441) |
| day 67 | −0.149**<br>(0.070) | −0.178**<br>(0.085) | −0.327<br>(0.443) |
| day 68 | −0.174**<br>(0.070) | −0.095<br>(0.085) | −0.477<br>(0.443) |
| day 69 | −0.034<br>(0.071) | −0.157*<br>(0.086) | −0.345<br>(0.450) |
| day 70 | −0.114<br>(0.072) | −0.376***<br>(0.087) | −0.244<br>(0.454) |
| Observations | 77,930 | 77,930 | 77,930 |
| R<sup>2</sup> | 0.502 | 0.272 | 0.469 |
| Adjusted R<sup>2</sup> | 0.500 | 0.269 | 0.466 |
| Residual Std. Error (df = 77610) | 0.704 | 0.851 | 4.441 |

*Note:*
\*p<0.1; \*\*p<0.05; \*\*\*p<0.01

**Table S12:**
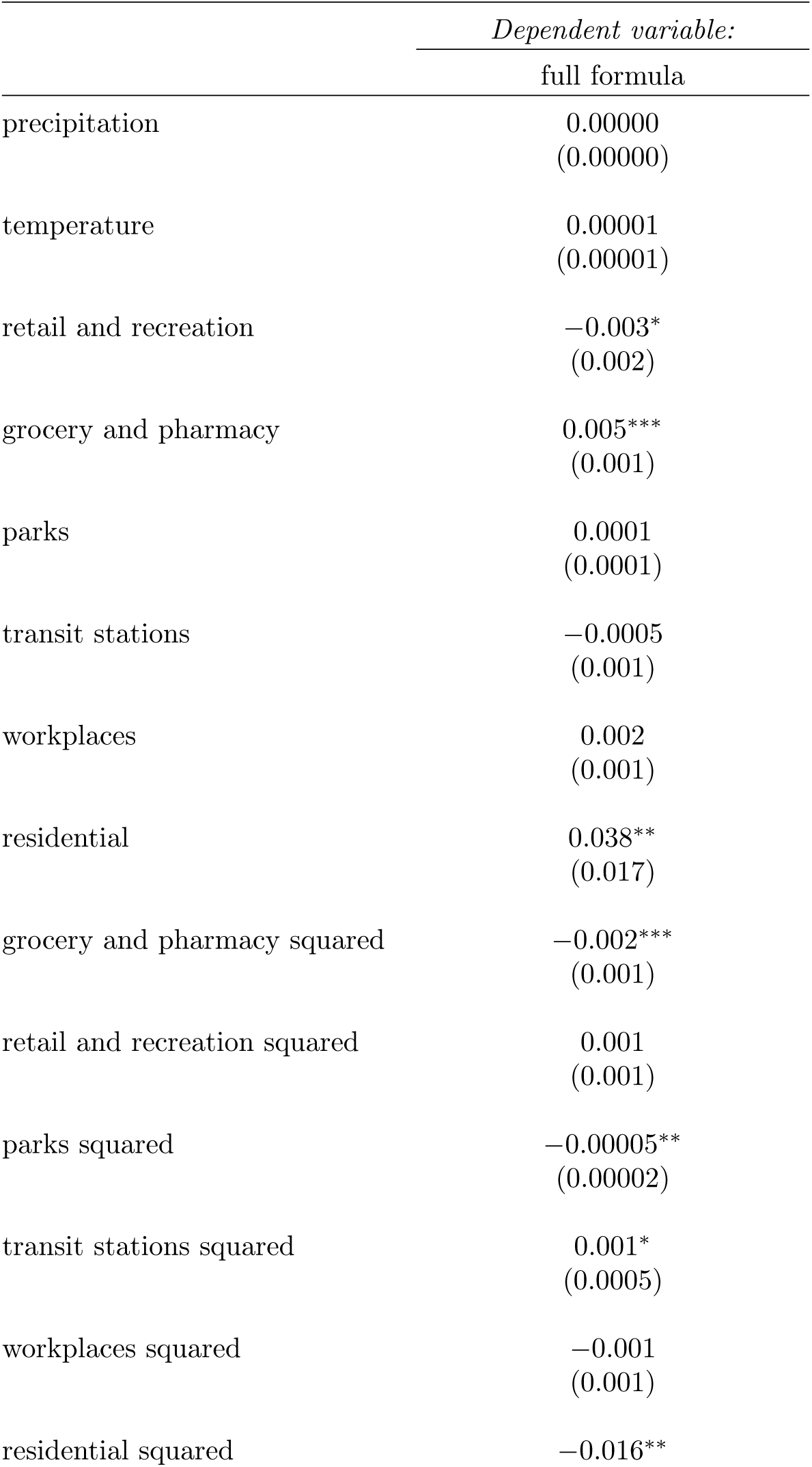
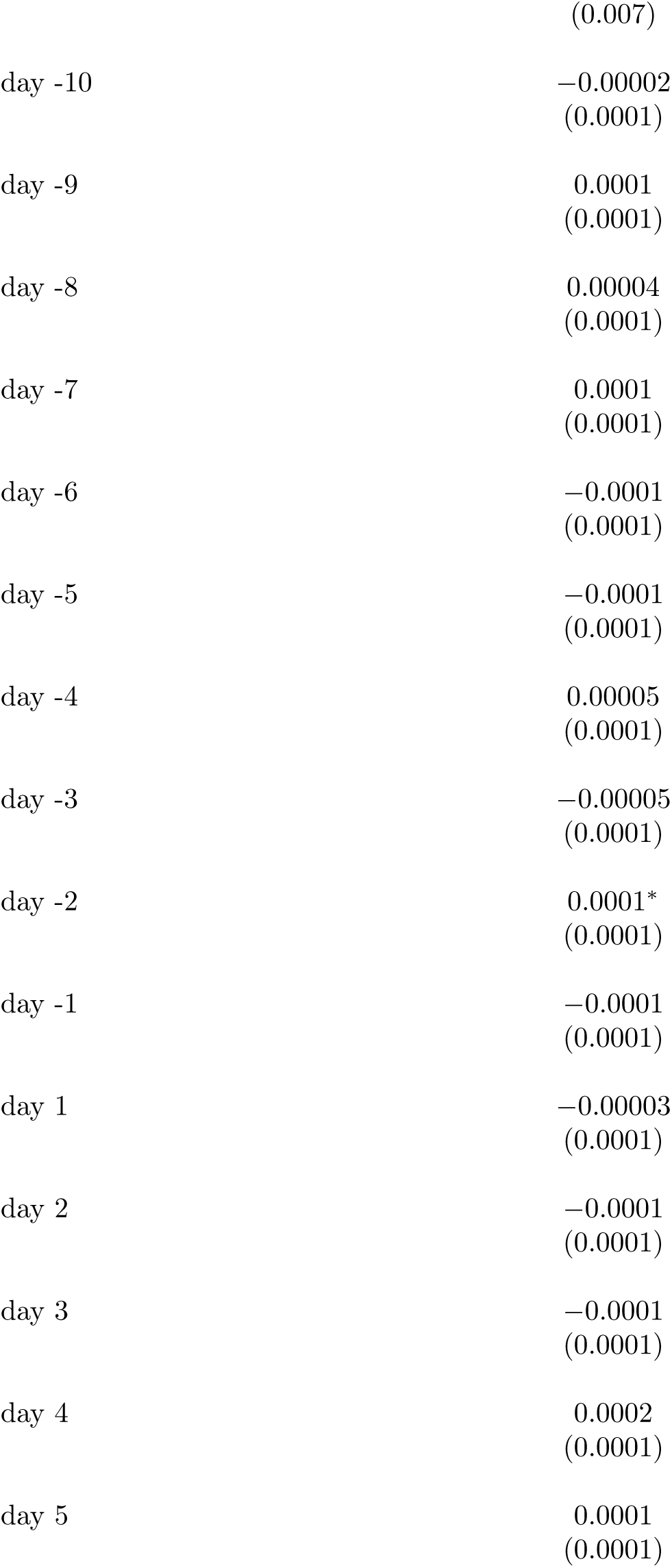
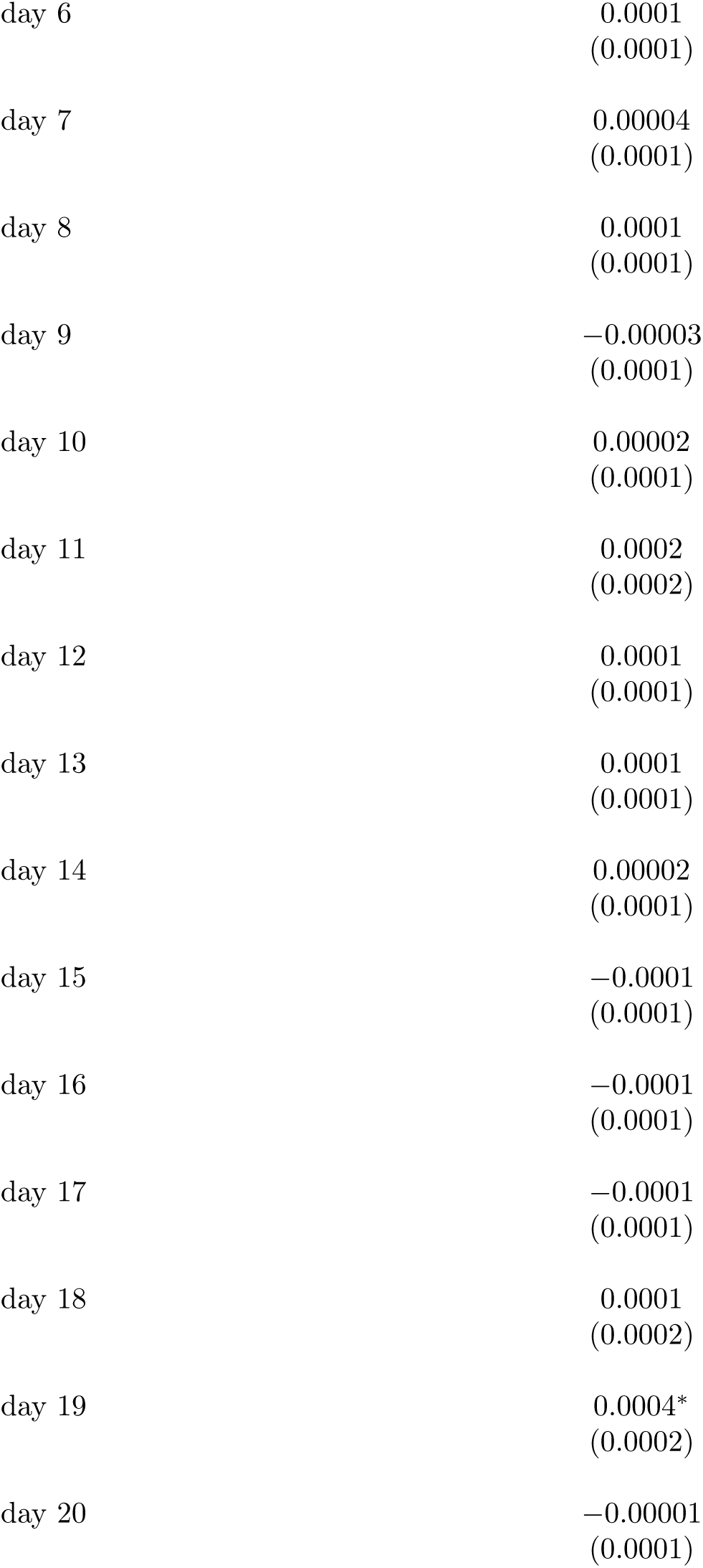
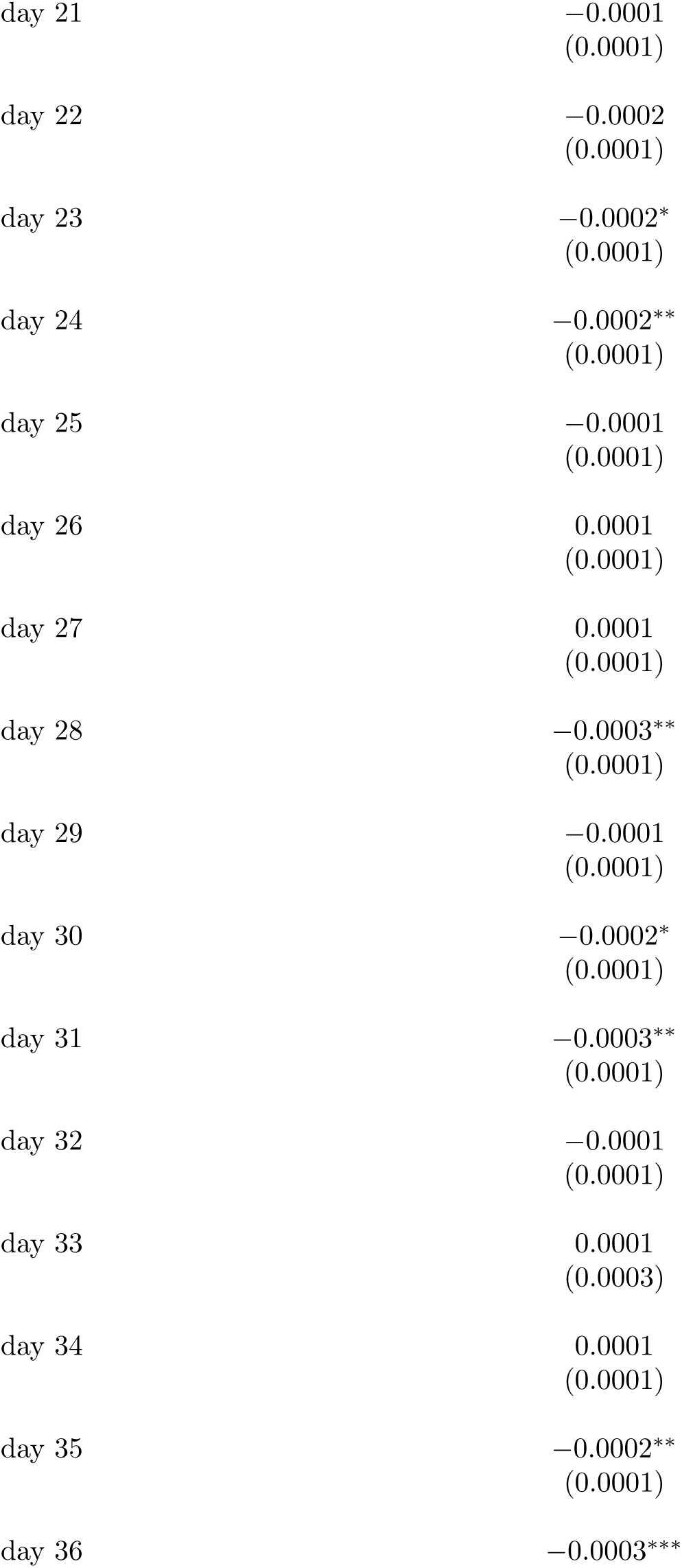
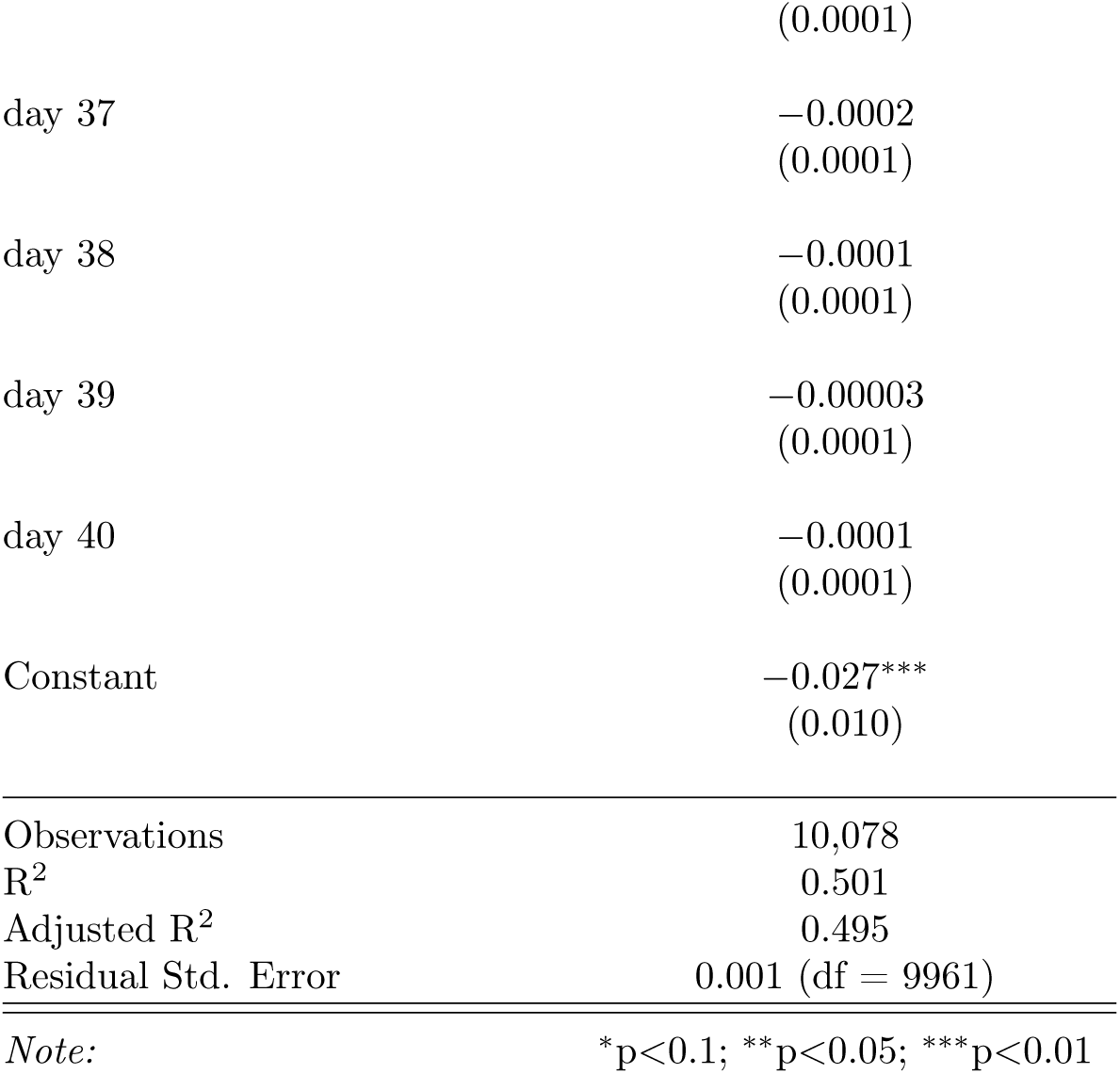
Event Study regression summary statistics of testing rate event study shown in fig. S2.

|  | *Dependent variable:* |
| --- | --- |
|  | full formula |
| precipitation | 0.00000<br>(0.00000) |
| temperature | 0.00001<br>(0.00001) |
| retail and recreation | −0.003*<br>(0.002) |
| grocery and pharmacy | 0.005***<br>(0.001) |
| parks | 0.0001<br>(0.0001) |
| transit stations | −0.0005<br>(0.001) |
| workplaces | 0.002<br>(0.001) |
| residential | 0.038**<br>(0.017) |
| grocery and pharmacy squared | −0.002***<br>(0.001) |
| retail and recreation squared | 0.001<br>(0.001) |
| parks squared | −0.00005**<br>(0.00002) |
| transit stations squared | 0.001*<br>(0.0005) |
| workplaces squared | −0.001<br>(0.001) |
| residential squared | −0.016** |
|  | (0.007) |
| day -10 | −0.00002<br>(0.0001) |
| day -9 | 0.0001<br>(0.0001) |
| day -8 | 0.00004<br>(0.0001) |
| day -7 | 0.0001<br>(0.0001) |
| day -6 | −0.0001<br>(0.0001) |
| day -5 | −0.0001<br>(0.0001) |
| day -4 | 0.00005<br>(0.0001) |
| day -3 | −0.00005<br>(0.0001) |
| day -2 | 0.0001*<br>(0.0001) |
| day -1 | −0.0001<br>(0.0001) |
| day 1 | −0.00003<br>(0.0001) |
| day 2 | −0.0001<br>(0.0001) |
| day 3 | −0.0001<br>(0.0001) |
| day 4 | 0.0002<br>(0.0001) |
| day 5 | 0.0001<br>(0.0001) |
| day 6 | 0.0001<br>(0.0001) |
| day 7 | 0.00004<br>(0.0001) |
| day 8 | 0.0001<br>(0.0001) |
| day 9 | −0.00003<br>(0.0001) |
| day 10 | 0.00002<br>(0.0001) |
| day 11 | 0.0002<br>(0.0002) |
| day 12 | 0.0001<br>(0.0001) |
| day 13 | 0.0001<br>(0.0001) |
| day 14 | 0.00002<br>(0.0001) |
| day 15 | −0.0001<br>(0.0001) |
| day 16 | −0.0001<br>(0.0001) |
| day 17 | −0.0001<br>(0.0001) |
| day 18 | 0.0001<br>(0.0002) |
| day 19 | 0.0004*<br>(0.0002) |
| day 20 | −0.00001<br>(0.0001) |
| day 21 | −0.0001<br>(0.0001) |
| day 22 | −0.0002<br>(0.0001) |
| day 23 | −0.0002*<br>(0.0001) |
| day 24 | −0.0002**<br>(0.0001) |
| day 25 | −0.0001<br>(0.0001) |
| day 26 | 0.0001<br>(0.0001) |
| day 27 | 0.0001<br>(0.0001) |
| day 28 | −0.0003**<br>(0.0001) |
| day 29 | −0.0001<br>(0.0001) |
| day 30 | −0.0002*<br>(0.0001) |
| day 31 | −0.0003**<br>(0.0001) |
| day 32 | −0.0001<br>(0.0001) |
| day 33 | 0.0001<br>(0.0003) |
| day 34 | 0.0001<br>(0.0001) |
| day 35 | −0.0002**<br>(0.0001) |
| day 36 | −0.0003*** |
|  | (0.0001) |
| day 37 | −0.0002<br>(0.0001) |
| day 38 | −0.0001<br>(0.0001) |
| day 39 | −0.00003<br>(0.0001) |
| day 40 | −0.0001<br>(0.0001) |
| Constant | −0.027***<br>(0.010) |
| Observations | 10,078 |
| R<sup>2</sup> | 0.501 |
| Adjusted R<sup>2</sup> | 0.495 |
| Residual Std. Error | 0.001 (df = 9961) |
| *Note:* | *p<0.1; **p<0.05; ***p<0.01 |

**Table S13:**
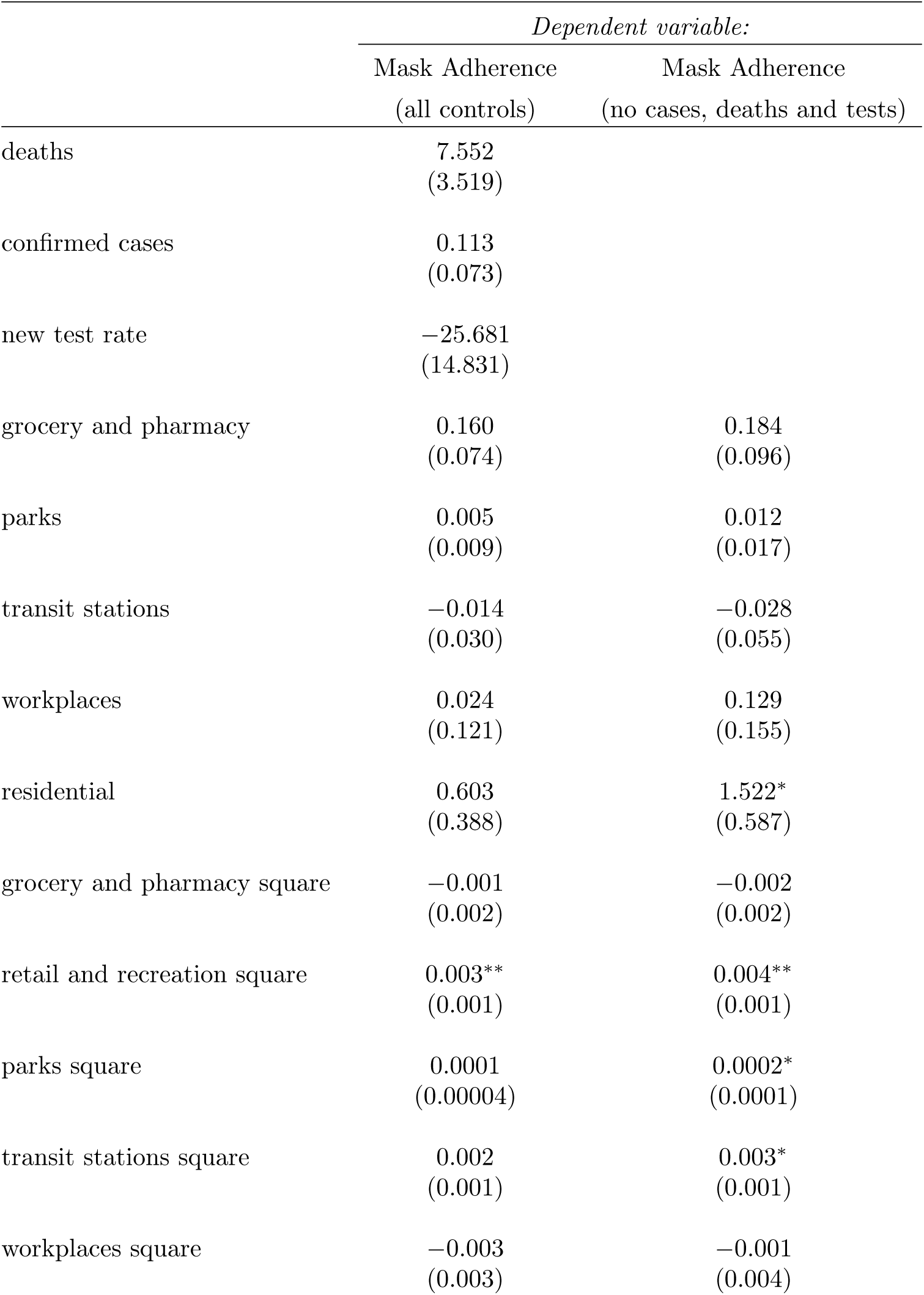
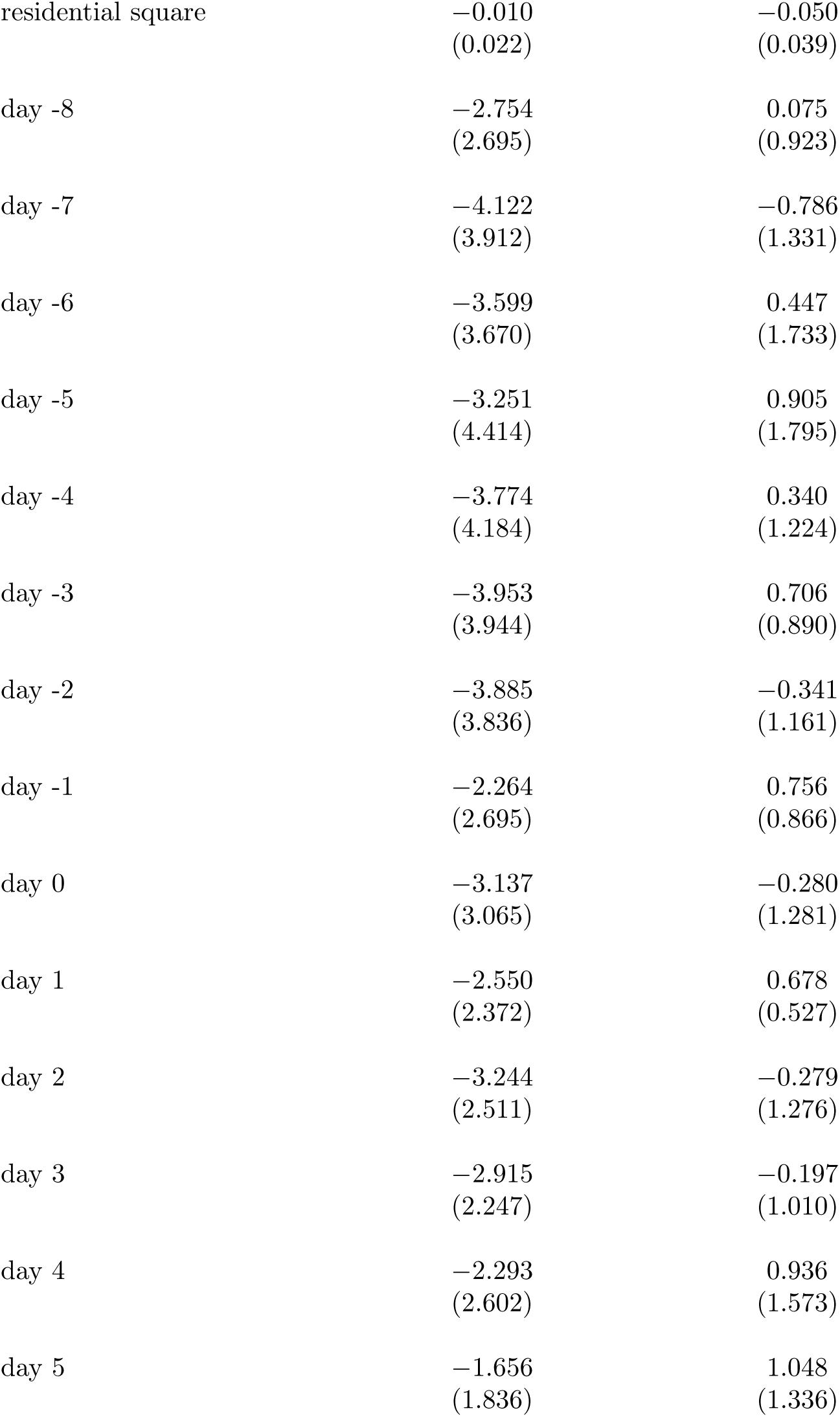
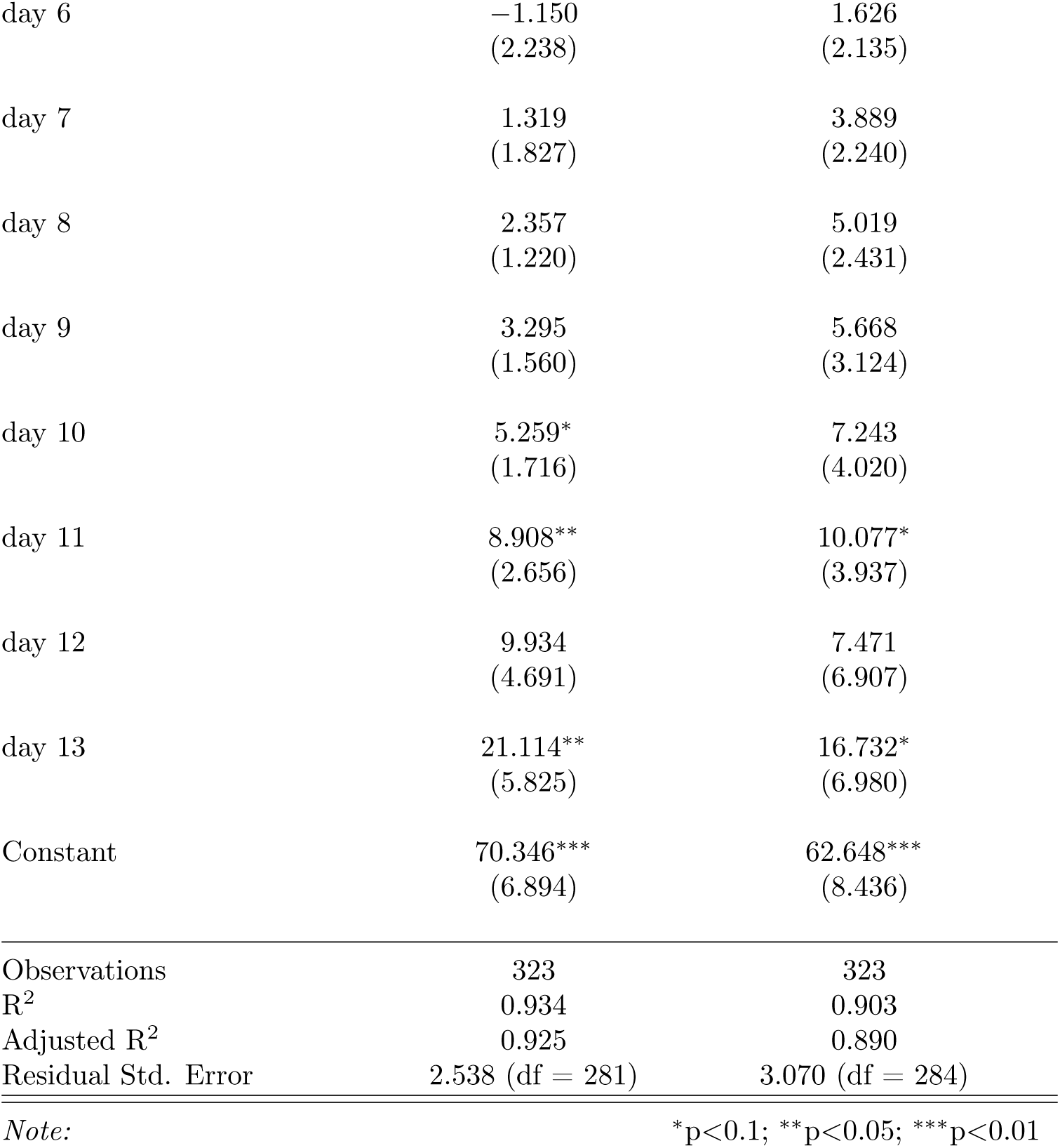
Event Study regression summary statistics for robustness check comparing controlling for cases, deaths and testing rate (fig. S7).

|  | *Dependent variable:* |  |
| --- | --- | --- |
|  | Mask Adherence | Mask Adherence |
|  | (all controls) | (no cases, deaths and tests) |
| deaths | 7.552<br>(3.519) |  |
| confirmed cases | 0.113<br>(0.073) |  |
| new test rate | −25.681<br>(14.831) |  |
| grocery and pharmacy | 0.160<br>(0.074) | 0.184<br>(0.096) |
| parks | 0.005<br>(0.009) | 0.012<br>(0.017) |
| transit stations | −0.014<br>(0.030) | −0.028<br>(0.055) |
| workplaces | 0.024<br>(0.121) | 0.129<br>(0.155) |
| residential | 0.603<br>(0.388) | 1.522*<br>(0.587) |
| grocery and pharmacy square | −0.001<br>(0.002) | −0.002<br>(0.002) |
| retail and recreation square | 0.003**<br>(0.001) | 0.004**<br>(0.001) |
| parks square | 0.0001<br>(0.00004) | 0.0002*<br>(0.0001) |
| transit stations square | 0.002<br>(0.001) | 0.003*<br>(0.001) |
| workplaces square | −0.003<br>(0.003) | −0.001<br>(0.004) |
| residential square | −0.010<br>(0.022) | −0.050<br>(0.039) |
| day -8 | −2.754<br>(2.695) | 0.075<br>(0.923) |
| day -7 | −4.122<br>(3.912) | −0.786<br>(1.331) |
| day -6 | −3.599<br>(3.670) | 0.447<br>(1.733) |
| day -5 | −3.251<br>(4.414) | 0.905<br>(1.795) |
| day -4 | −3.774<br>(4.184) | 0.340<br>(1.224) |
| day -3 | −3.953<br>(3.944) | 0.706<br>(0.890) |
| day -2 | −3.885<br>(3.836) | −0.341<br>(1.161) |
| day -1 | −2.264<br>(2.695) | 0.756<br>(0.866) |
| day 0 | −3.137<br>(3.065) | −0.280<br>(1.281) |
| day 1 | −2.550<br>(2.372) | 0.678<br>(0.527) |
| day 2 | −3.244<br>(2.511) | −0.279<br>(1.276) |
| day 3 | −2.915<br>(2.247) | −0.197<br>(1.010) |
| day 4 | −2.293<br>(2.602) | 0.936<br>(1.573) |
| day 5 | −1.656<br>(1.836) | 1.048<br>(1.336) |
| day 6 | -1.150<br>(2.238) | 1.626<br>(2.135) |
| day 7 | 1.319<br>(1.827) | 3.889<br>(2.240) |
| day 8 | 2.357<br>(1.220) | 5.019<br>(2.431) |
| day 9 | 3.295<br>(1.560) | 5.668<br>(3.124) |
| day 10 | 5.259*<br>(1.716) | 7.243<br>(4.020) |
| day 11 | 8.908**<br>(2.656) | 10.077*<br>(3.937) |
| day 12 | 9.934<br>(4.691) | 7.471<br>(6.907) |
| day 13 | 21.114**<br>(5.825) | 16.732*<br>(6.980) |
| Constant | 70.346***<br>(6.894) | 62.648***<br>(8.436) |
| Observations | 323 | 323 |
| R<sup>2</sup> | 0.934 | 0.903 |
| Adjusted R<sup>2</sup> | 0.925 | 0.890 |
| Residual Std. Error | 2.538 (df = 281) | 3.070 (df = 284) |

*Note:*
\*p<0.1; \*\*p<0.05; \*\*\*p<0.01

**Table S14:** Regression statistics of mask adherence on confirmed new cases (1) and deaths (2)

|  | *Dependent variable:* |  |
| --- | --- | --- |
|  | confirmed cases | deaths |
|  | (1) | (2) |
| compliance | −1.675***<br>(0.064) | −0.017***<br>(0.001) |
| new test rate | 1,528.504***<br>(108.597) | 14.615***<br>(1.225) |
| precipitation | −0.419***<br>(0.056) | −0.003***<br>(0.001) |
| temperature | −1.060***<br>(0.058) | −0.004***<br>(0.001) |
| grocery and pharmacy | 0.453***<br>(0.066) | 0.002***<br>(0.001) |
| parks | −0.170***<br>(0.016) | −0.002***<br>(0.0002) |
| transit stations | −0.237***<br>(0.043) | −0.005***<br>(0.0005) |
| workplaces | 2.261***<br>(0.158) | 0.015***<br>(0.002) |
| residential | 5.755***<br>(0.420) | 0.021***<br>(0.005) |
| grocery and pharmacy square | 0.008***<br>(0.002) | 0.0002***<br>(0.00003) |
| retail and recreation square | 0.0002<br>(0.001) | −0.00002<br>(0.00002) |
| parks square | 0.0004***<br>(0.0001) | 0.00000***<br>(0.00000) |
| transit stations square | −0.013***<br>(0.001) | −0.0002***<br>(0.00001) |
| workplaces square | 0.017***<br>(0.003) | 0.0001***<br>(0.00003) |
| residential square | −0.012<br>(0.025) | 0.001***<br>(0.0003) |
| Constant | 191.442***<br>(5.384) | 1.874***<br>(0.061) |
| Observations | 3,841 | 3,841 |
| R<sup>2</sup> | 0.502 | 0.343 |
| *Note:* | *p<0.1; **p<0.05; ***p<0.01 |  |

**Table S15:** Multi-linear survey regression statistics of community mask adherence and community mask attitudes on COVID-19 Deaths and Cases in 68 countries over all waves.

|  | *Dependent variable (per 100K):* |  |  |  |
| --- | --- | --- | --- | --- |
|  | Cases | Deaths | Cases | Deaths |
|  | (1) | (2) | (3) | (4) |
| Community Mask Attitudes | −0.530***<br>(0.055) | −0.035***<br>(0.002) |  |  |
| Community Mask Adherence |  |  | −0.497***<br>(0.106) | −0.042***<br>(0.002) |
| population density | −0.577***<br>(0.009) | −0.021***<br>(0.0003) | −0.574***<br>(0.013) | −0.021***<br>(0.0004) |
| human development index | 309.005***<br>(16.045) | 34.078***<br>(0.427) | 238.798***<br>(24.962) | 33.000***<br>(0.618) |
| new tests smoothed per thousand | 453.869***<br>(3.267) | 1.206***<br>(0.050) | 460.777***<br>(6.254) | 1.256***<br>(0.073) |
| retail and recreation | −4.134***<br>(0.178) | −0.154***<br>(0.005) | −4.653***<br>(0.259) | −0.162***<br>(0.008) |
| grocery and pharmacy | 3.294***<br>(0.148) | 0.033***<br>(0.004) | 2.966***<br>(0.212) | 0.033***<br>(0.006) |
| transit stations | −2.170***<br>(0.129) | 0.004<br>(0.004) | −1.877***<br>(0.202) | 0.007<br>(0.005) |
| workplaces | 13.075***<br>(0.203) | 0.186***<br>(0.005) | 12.518***<br>(0.315) | 0.170***<br>(0.007) |
| residential | 32.767***<br>(0.491) | 0.862***<br>(0.012) | 30.551***<br>(0.754) | 0.814***<br>(0.017) |
| Constant | −65.551***<br>(12.087) | −14.552***<br>(0.309) | −18.362<br>(19.673) | −13.298***<br>(0.437) |
| Observations | 216,663 | 216,663 | 97,802 | 97,802 |
| *Note:* |  |  | *p<0.1; **p<0.05; ***p<0.01 |  |

**Table S16:** Multi-linear survey regression statistics of community mask adherence on COVID-19 Cases in 68 countries per wave.

|  | *Dependent variable:* |  |  |  |  |  |  |  |
| --- | --- | --- | --- | --- | --- | --- | --- | --- |
|  | new cases |  |  |  |  |  |  |  |
|  | (1) | (2) | (3) | (4) | (5) | (6) | (7) | (8) |
| Community Mask Adherence | −0.014<br>(0.014) | −0.051***<br>(0.016) | −0.104***<br>(0.018) | −0.137***<br>(0.017) | −0.080***<br>(0.015) | −0.103***<br>(0.018) | 0.012*<br>(0.007) | 0.008<br>(0.033) |
| population density | −0.053***<br>(0.003) | −0.056***<br>(0.003) | −0.057***<br>(0.003) | −0.091***<br>(0.004) | −0.067***<br>(0.003) | −0.071***<br>(0.005) | −0.033***<br>(0.001) | −0.017***<br>(0.001) |
| human development index | 67.153***<br>(4.875) | 13.143***<br>(3.502) | 47.721***<br>(4.136) | 75.832***<br>(4.204) | 160.982***<br>(5.244) | 198.787***<br>(8.666) | −4.387<br>(3.002) | −1.285<br>(4.790) |
| new tests smoothed per thousand | 63.066***<br>(0.654) | 66.997***<br>(0.426) | 53.548***<br>(0.504) | 40.492***<br>(0.529) | 14.530***<br>(0.677) | 24.274***<br>(0.719) | 39.538***<br>(0.316) | 59.118***<br>(1.353) |
| retail and recreation | 0.545***<br>(0.035) | 0.234***<br>(0.029) | 0.064*<br>(0.034) | −0.394***<br>(0.059) | −2.577***<br>(0.057) | −1.464***<br>(0.096) | −1.403***<br>(0.040) | −1.913***<br>(0.129) |
| grocery and pharmacy | 0.217***<br>(0.026) | 0.323***<br>(0.031) | 0.818***<br>(0.038) | 1.447***<br>(0.059) | 3.237***<br>(0.076) | 1.188***<br>(0.108) | 0.212***<br>(0.036) | −0.347***<br>(0.038) |
| transit stations | −0.647***<br>(0.035) | −1.124***<br>(0.034) | −0.409***<br>(0.025) | −0.567***<br>(0.035) | 0.312***<br>(0.036) | 0.499***<br>(0.079) | 0.730***<br>(0.042) | 1.050***<br>(0.050) |
| workplaces | 2.289***<br>(0.060) | 1.316***<br>(0.052) | 1.759***<br>(0.052) | 1.874***<br>(0.048) | −0.471***<br>(0.063) | 2.200***<br>(0.061) | 0.326***<br>(0.037) | 0.175***<br>(0.052) |
| residential | 6.463***<br>(0.182) | 3.157***<br>(0.146) | 6.427***<br>(0.172) | 7.122***<br>(0.157) | 2.467***<br>(0.198) | 5.959***<br>(0.213) | 1.120***<br>(0.048) | −0.205<br>(0.263) |
| Constant | −55.461***<br>(3.459) | −8.636***<br>(2.518) | −24.440***<br>(3.259) | −46.732***<br>(3.060) | −122.869***<br>(3.754) | −122.170***<br>(6.643) | 8.818***<br>(2.351) | 0.589<br>(4.311) |
| Observations | 25,666 | 31,614 | 28,066 | 22,971 | 21,006 | 7,802 | 18,554 | 17,658 |

Note:
\*p<0.1; \*\*p<0.05; \*\*\*p<0.01

**Table S17:** Multi-linear survey regression statistics of community mask attitudes on COVID-19 Cases in 68 countries per wave.

|  | *Dependent variable:* |  |  |  |  |  |  |  |
| --- | --- | --- | --- | --- | --- | --- | --- | --- |
|  | new cases |  |  |  |  |  |  |  |
|  | (1) | (2) | (3) | (4) | (5) | (6) | (7) | (8) |
| Community Mask Attitudes | −0.043***<br>(0.016) | −0.046***<br>(0.015) | −0.105***<br>(0.016) | −0.103***<br>(0.016) | −0.082***<br>(0.013) | −0.077***<br>(0.013) | −0.006<br>(0.004) | −0.001<br>(0.014) |
| population density | −0.048***<br>(0.002) | −0.058***<br>(0.003) | −0.055***<br>(0.003) | −0.087***<br>(0.004) | −0.068***<br>(0.003) | −0.070***<br>(0.003) | −0.031***<br>(0.001) | −0.016***<br>(0.001) |
| human development index | 70.177***<br>(4.932) | 15.275***<br>(3.486) | 46.990***<br>(4.171) | 68.568***<br>(4.407) | 170.880***<br>(5.192) | 200.450***<br>(5.549) | −1.349<br>(1.852) | 0.290<br>(3.125) |
| new tests smoothed per thousand | 62.866***<br>(0.662) | 66.708***<br>(0.440) | 53.623***<br>(0.487) | 39.920***<br>(0.578) | 13.285***<br>(0.705) | 23.600***<br>(0.473) | 39.678***<br>(0.211) | 58.457***<br>(0.709) |
| retail and recreation | 0.498***<br>(0.036) | 0.250***<br>(0.031) | 0.069*<br>(0.036) | −0.401***<br>(0.061) | −2.588***<br>(0.058) | −1.487***<br>(0.060) | −1.375***<br>(0.026) | −1.755***<br>(0.072) |
| grocery and pharmacy | 0.196***<br>(0.027) | 0.298***<br>(0.032) | 0.812***<br>(0.038) | 1.470***<br>(0.067) | 3.328***<br>(0.073) | 1.202***<br>(0.065) | 0.266***<br>(0.025) | −0.363***<br>(0.022) |
| transit stations | −0.668***<br>(0.034) | −1.152***<br>(0.035) | −0.375***<br>(0.026) | −0.604***<br>(0.035) | 0.331***<br>(0.036) | 0.513***<br>(0.045) | 0.677***<br>(0.028) | 0.970***<br>(0.032) |
| workplaces | 2.263***<br>(0.060) | 1.405***<br>(0.054) | 1.712***<br>(0.053) | 1.793***<br>(0.051) | −0.378***<br>(0.065) | 2.124***<br>(0.037) | 0.308***<br>(0.025) | 0.150***<br>(0.034) |
| residential | 6.326***<br>(0.182) | 3.258***<br>(0.148) | 6.429***<br>(0.173) | 6.968***<br>(0.166) | 2.763***<br>(0.200) | 5.847***<br>(0.125) | 1.082***<br>(0.032) | −0.084<br>(0.147) |
| Constant | −57.361***<br>(3.533) | −8.805***<br>(2.670) | −24.143***<br>(3.361) | −44.760***<br>(3.215) | −128.983***<br>(3.847) | −125.463***<br>(4.293) | 6.501***<br>(1.508) | 0.781<br>(2.592) |
| Observations | 25,359 | 31,058 | 27,689 | 22,663 | 20,804 | 15,721 | 37,708 | 35,650 |

Note:
\*p<0.1; \*\*p<0.05; \*\*\*p<0.01

**Table S18:** Multi-linear survey regression statistics of community mask adherence on COVID-19 cases in 68 countries per wave.

|  | *Dependent variable:* |  |  |  |  |  |  |  |
| --- | --- | --- | --- | --- | --- | --- | --- | --- |
|  | new deaths |  |  |  |  |  |  |  |
|  | (1) | (2) | (3) | (4) | (5) | (6) | (7) | (8) |
| Community Mask Adherence | −0.003***<br>(0.0005) | −0.005***<br>(0.0005) | −0.006***<br>(0.001) | −0.006***<br>(0.001) | −0.004***<br>(0.0004) | −0.005***<br>(0.001) | −0.002***<br>(0.0003) | −0.003***<br>(0.0003) |
| population density | −0.002***<br>(0.0001) | −0.002***<br>(0.0001) | −0.003***<br>(0.0001) | −0.003***<br>(0.0001) | −0.002***<br>(0.0001) | −0.002***<br>(0.0002) | −0.002***<br>(0.0001) | −0.001***<br>(0.0001) |
| human development index | 4.854***<br>(0.141) | 3.953***<br>(0.116) | 4.324***<br>(0.131) | 4.407***<br>(0.144) | 5.486***<br>(0.156) | 6.819***<br>(0.318) | 2.659***<br>(0.101) | 4.644***<br>(0.157) |
| new tests smoothed per thousand | 0.173***<br>(0.024) | 0.527***<br>(0.016) | 0.473***<br>(0.020) | 0.318***<br>(0.020) | −0.310***<br>(0.018) | −0.164***<br>(0.031) | 0.075***<br>(0.012) | −0.066***<br>(0.017) |
| retail and recreation | −0.003**<br>(0.001) | −0.001<br>(0.001) | −0.010***<br>(0.001) | −0.017***<br>(0.002) | −0.052***<br>(0.001) | −0.042***<br>(0.003) | −0.043***<br>(0.001) | −0.086***<br>(0.002) |
| grocery and pharmacy | −0.006***<br>(0.001) | −0.004***<br>(0.001) | 0.004***<br>(0.001) | 0.023***<br>(0.002) | 0.043***<br>(0.002) | 0.020***<br>(0.003) | 0.005***<br>(0.001) | 0.004***<br>(0.001) |
| transit stations | −0.007***<br>(0.001) | −0.020***<br>(0.001) | −0.007***<br>(0.001) | −0.015***<br>(0.001) | 0.004***<br>(0.001) | 0.014***<br>(0.002) | 0.027***<br>(0.001) | 0.060***<br>(0.002) |
| workplaces | 0.040***<br>(0.002) | 0.029***<br>(0.001) | 0.032***<br>(0.001) | 0.034***<br>(0.001) | −0.016***<br>(0.001) | 0.023***<br>(0.002) | 0.002*<br>(0.001) | −0.021***<br>(0.002) |
| residential | 0.142***<br>(0.005) | 0.099***<br>(0.004) | 0.140***<br>(0.005) | 0.155***<br>(0.004) | 0.052***<br>(0.004) | 0.121***<br>(0.006) | 0.038***<br>(0.002) | 0.015***<br>(0.003) |
| Constant | −2.919***<br>(0.100) | −2.121***<br>(0.080) | −2.144***<br>(0.091) | −2.526***<br>(0.103) | −3.428***<br>(0.110) | −3.845***<br>(0.231) | −1.123***<br>(0.075) | −2.618***<br>(0.112) |
| Observations | 25,666 | 31,614 | 28,066 | 22,971 | 21,006 | 7,802 | 18,554 | 17,658 |

Note:
\*p<0.1; \*\*p<0.05; \*\*\*p<0.01

**Table S19:** Multi-linear survey regression statistics of community mask attitudes on COVID-19 deaths in 68 countries per wave.

|  | *Dependent variable:* |  |  |  |  |  |  |  |
| --- | --- | --- | --- | --- | --- | --- | --- | --- |
|  | new deaths |  |  |  |  |  |  |  |
|  | (1) | (2) | (3) | (4) | (5) | (6) | (7) | (8) |
| Community Mask Attitudes | −0.004***<br>(0.001) | −0.003***<br>(0.0005) | −0.005***<br>(0.001) | −0.005***<br>(0.001) | −0.004***<br>(0.0004) | −0.004***<br>(0.001) | −0.002***<br>(0.0002) | −0.003***<br>(0.0002) |
| population density | −0.002***<br>(0.0001) | −0.002***<br>(0.0001) | −0.002***<br>(0.0001) | −0.003***<br>(0.0001) | −0.002***<br>(0.0001) | −0.002***<br>(0.0001) | −0.002***<br>(0.00004) | −0.001***<br>(0.00003) |
| human development index | 5.058***<br>(0.144) | 3.885***<br>(0.116) | 4.076***<br>(0.126) | 4.168***<br>(0.154) | 5.635***<br>(0.150) | 6.819***<br>(0.203) | 2.655***<br>(0.063) | 4.494***<br>(0.098) |
| new tests smoothed per thousand | 0.146***<br>(0.026) | 0.524***<br>(0.016) | 0.504***<br>(0.019) | 0.293***<br>(0.021) | −0.351***<br>(0.018) | −0.188***<br>(0.019) | 0.082***<br>(0.008) | −0.071***<br>(0.011) |
| retail and recreation | −0.006***<br>(0.001) | 0.001<br>(0.001) | −0.010***<br>(0.001) | −0.016***<br>(0.002) | −0.053***<br>(0.001) | −0.045***<br>(0.002) | −0.042***<br>(0.001) | −0.086***<br>(0.001) |
| grocery and pharmacy | −0.008***<br>(0.001) | −0.005***<br>(0.001) | 0.005***<br>(0.001) | 0.024***<br>(0.002) | 0.045***<br>(0.002) | 0.019***<br>(0.002) | 0.006***<br>(0.001) | 0.004***<br>(0.001) |
| transit stations | −0.007***<br>(0.001) | −0.019***<br>(0.001) | −0.006***<br>(0.001) | −0.017***<br>(0.001) | 0.005***<br>(0.001) | 0.017***<br>(0.001) | 0.026***<br>(0.001) | 0.060***<br>(0.001) |
| workplaces | 0.037***<br>(0.002) | 0.031***<br>(0.001) | 0.032***<br>(0.001) | 0.032***<br>(0.001) | −0.015***<br>(0.001) | 0.021***<br>(0.001) | 0.003***<br>(0.001) | −0.023***<br>(0.001) |
| residential | 0.136***<br>(0.005) | 0.107***<br>(0.004) | 0.135***<br>(0.005) | 0.152***<br>(0.005) | 0.056***<br>(0.004) | 0.118***<br>(0.004) | 0.039***<br>(0.002) | 0.013***<br>(0.002) |
| Constant | −3.078***<br>(0.102) | −2.121***<br>(0.084) | −2.033***<br>(0.091) | −2.450***<br>(0.111) | −3.553***<br>(0.111) | −3.922***<br>(0.148) | −1.167***<br>(0.048) | −2.539***<br>(0.070) |
| Observations | 25,359 | 31,058 | 27,689 | 22,663 | 20,804 | 15,721 | 37,708 | 35,650 |

Note:
\*p<0.1; \*\*p<0.05; \*\*\*p<0.01

## References

[1] Dong E, Du H, Gardner L. An interactive web-based dashboard to track COVID-19 in real time. The Lancet infectious diseases. 2020;20(5):533–534.

[2] Howard J, Huang A, Li Z, Tufekci Z, Zdimal V, van der Westhuizen HM, et al. Face masks against COVID-19: an evidence review. 2020;.

[3] Liang M, Gao L, Cheng C, Zhou Q, Uy JP, Heiner K, et al. Efficacy of face mask in preventing respiratory virus transmission: a systematic review and meta-analysis. Travel Medicine and Infectious Disease. 2020; p. 101751.

[4] Chu DK, Akl EA, Duda S, Solo K, Yaacoub S, Schünemann HJ, et al. Physical distancing, face masks, and eye protection to prevent person-to-person transmission of SARS-CoV-2 and COVID-19: a systematic review and meta-analysis. The Lancet. 2020;.

[5] CDC. Scientific Brief: Community Use of Cloth Masks to Control the Spread of SARS-CoV-2. Science Briefs;.

[6] Leffler CT, Ing EB, Lykins JD, Hogan MC, McKeown CA, Grzybowski A. Association of country-wide coronavirus mortality with demographics, testing, lockdowns, and public wearing of masks. Update June 2, 2020. medRxiv. 2020;.

[7] Wang X, Ferro EG, Zhou G, Hashimoto D, Bhatt DL. Association between universal masking in a health care system and SARS-CoV-2 positivity among health care workers. JAMA. 2020;324(7):703–704.

[8] Lyu W, Wehby GL. Community Use Of Face Masks And COVID-19: Evidence From A Natural Experiment Of State Mandates In The US: Study examines impact on COVID-19 growth rates associated with state government mandates requiring face mask use in public. Health affairs. 2020;39(8):1419–1425.

[9] Van Dyke ME. Trends in County-Level COVID-19 Incidence in Counties With and Without a Mask Mandate— Kansas, June 1–August 23, 2020. MMWR Morbidity and Mortality Weekly Report. 2020;69.

[10] Mitze T, Kosfeld R, Rode J, Wälde K. Face Masks Considerably Reduce COVID-19 Cases in Germany. 2020;.

[11] Eikenberry SE, Mancuso M, Iboi E, Phan T, Eikenberry K, Kuang Y, et al. To mask or not to mask: Modeling the potential for face mask use by the general public to curtail the COVID-19 pandemic. Infectious Disease Modelling. 2020;.

[12] Li T, Liu Y, Li M, Qian X, Dai SY. Mask or no mask for COVID-19: A public health and market study. PloS one. 2020;15(8):e0237691.

[13] Fisman DN, Greer AL, Tuite AR. Bidirectional impact of imperfect mask use on reproduction number of COVID-19: A next generation matrix approach. Infectious Disease Modelling. 2020;5:405–408.

[14] Mittal R, Ni R, Seo JH. The flow physics of COVID-19. Journal of fluid Mechanics. 2020;894.

[15] Verma S, Dhanak M, Frankenfield J. Visualizing droplet dispersal for face shields and masks with exhalation valves. Physics of Fluids. 2020;32(9):091701.

[16] Dugdale CM, Walensky RP. Filtration efficiency, effectiveness, and availability of N95 face masks for COVID-19 prevention. JAMA Internal Medicine. 2020;.

[17] Haischer MH, Beilfuss R, Hart MR, Opielinski L, Wrucke D, Zirgaitis G, et al. Who is wearing a mask? Gender-, age-, and location-related differences during the COVID-19 pandemic. Plos one. 2020;15(10):e0240785.

[18] Nikolov P, Pape A, Tonguc O, Williams C. Predictors of Social Distancing and Mask-Wearing Behavior: Panel Survey in Seven US States. arXiv preprint arXiv:200913103. 2020;.

[19] Howard MC. Gender, face mask perceptions, and face mask wearing: Are men being dangerous during the COVID-19 pandemic? Personality and individual differences. 2020;170:110417.

[20] Bhasin T, Butcher C, Gordon E, Hallward M, LeFebvre R. Does Karen wear a mask? The gendering of COVID-19 masking rhetoric. International Journal of Sociology and Social Policy. 2020;.

[21] Chernozhukov V, Kasaha H, Schrimpf P. Causal impact of masks, policies, behavior on early COVID-19 pandemic in the US. Journal of Econometrics. 2021;.

[22] Joo H, Miller GF, Sunshine G, Gakh M, Pike J, Havers FP, et al. Decline in COVID-19 hospitalization growth rates associated with statewide mask mandates—10 states, March–October 2020. 2021;.

[23] Bundgaard H, Bundgaard JS, Raaschou-Pedersen DET, von Buchwald C, Todsen T, Norsk JB, et al. Effectiveness of adding a mask recommendation to other public health measures to prevent SARS-CoV-2 infection in Danish mask wearers: a randomized controlled trial. Annals of Internal Medicine. 2020;.

[24] Scerri M, Grech V. To wear or not to wear? Adherence to face mask use during the COVID-19 and Spanish influenza pandemics. Early human development. 2020; p. 105253.

[25] Collis A, Garimella K, Moehring A, Rahimian MA, Babalola S, Shattuck D, et al. Global survey on COVID-19 beliefs, behaviors, and norms. MIT Sloan School of Management Technical Report. 2020;.

[26] Bhattacharjee S, Bahl P, Chughtai AA, MacIntyre CR. Last-resort strategies during mask shortages: optimal design features of cloth masks and decontamination of disposable masks during the COVID-19 pandemic. BMJ open respiratory research. 2020;7(1):e000698.

[27] de Man P, van Straten B, van den Dobbelsteen J, van der Eijk A, Horeman T, Koeleman H. Sterilization of disposable face masks by means of standardized dry and steam sterilization processes; an alternative in the fight against mask shortages due to COVID-19. Journal of Hospital Infection. 2020;105(2):356–357.

[28] Wang M, Zhou M, Ji G, Ye L, Cheng Y, Feng Z, et al. Mask crisis during the COVID-19 outbreak. Eur Rev Med Pharmacol Sci. 2020;24(6):3397–9.

[29] Neal DT, Wood W, Quinn JM. Habits—A repeat performance. Current directions in psychological science. 2006;15(4):198–202.

[30] Riekert KA, Ockene JK, Pbert L. The handbook of health behavior change. Springer Publishing Company; 2013.

[31] Borusyak K, Jaravel X. Revisiting event study designs. Available at SSRN 2826228. 2017;.

[32] Chang S, Pierson E, Koh PW, Gerardin J, Redbird B, Grusky D, et al. Mobility network models of COVID-19 explain inequities and inform reopening. Nature. 2020; p. 1–6.

[33] Xie Y. The log-multiplicative layer effect model for comparing mobility tables. American sociological review. 1992; p. 380–395.

[34] Geiger D, Verma T, Pearl J. d-separation: From theorems to algorithms. In: Machine Intelligence and Pattern Recognition. vol. 10. Elsevier; 1990. p. 139–148.

[35] Wright AL, Chawla G, Chen L, Farmer A. Tracking Mask Mandates During the Covid-19 Pandemic. University of Chicago, Becker Friedman Institute for Economics Working Paper. 2020;(2020-104).

[36] Holtz D, Zhao M, Benzell SG, Cao CY, Rahimian MA, Yang J, et al. Interdependence and the Cost of Uncoordinated Responses to COVID-19. 2020;.

[37] Pearce N, Vandenbroucke JP, VanderWeele TJ, Greenland S. Accurate statistics on COVID-19 are essential for policy guidance and decisions; 2020.

[38] Grother PJ, Ngan ML, Hanaoka KK. Ongoing face recognition vendor test (frvt) part 2: Identification. 2018;.

[39] Chen J, Cremer JF, Zarei K, Segre AM, Polgreen PM. Using computer vision and depth sensing to measure healthcare worker-patient contacts and personal protective equipment adherence within hospital rooms. In: Open forum infectious diseases. vol. 3. Oxford University Press; 2016.

[40] Draughon GT, Sun P, Lynch JP. Implementation of a Computer Vision Framework for Tracking and Visualizing Face Mask Usage in Urban Environments. In: 2020 IEEE International Smart Cities Conference (ISC2). IEEE;. p. 1–8.

[41] Gibson KL, Smith JM. The emperor’s new masks: On demographic differences and disguises. In: Proceedings of the IEEE Conference on Computer Vision and Pattern Recognition Workshops; 2015. p. 57–64.

[42] Farrow DC, Brooks LC, Rumack A, Tibshirani RJ, Rosenfeld R. Delphi Epidata API. 2015;.

[43] Barkay N, Cobb C, Eilat R, Galili T, Haimovich D, LaRocca S, et al. Weights and Methodology Brief for the COVID-19 Symptom Survey by University of Maryland and Carnegie Mellon University, in Partnership with Facebook. arXiv preprint arXiv:200914675. 2020;.

[44] Betsch C, Korn L, Sprengholz P, Felgendreff L, Eitze S, Schmid P, et al. Social and behavioral consequences of mask policies during the COVID-19 pandemic. Proceedings of the National Academy of Sciences. 2020;117(36):21851–21853.

[45] Chetty R, Friedman J, Hendren N, Stepner M. The economic impacts of COVID-19: Evidence from a new public database built from private sector data. Opportunity Insights. 2020;.

[46] Fauci AS, Lane HC, Redfield RR. Covid-19—navigating the uncharted; 2020.

[47] Velavan TP, Meyer CG. The COVID-19 epidemic. Tropical medicine & international health. 2020;25(3):278.

[48] Flaxman S, Mishra S, Gandy A, Unwin HJT, Mellan TA, Coupland H, et al. Estimating the effects of non-pharmaceutical interventions on COVID-19 in Europe. Nature. 2020;584(7820):257–261.

[49] Ferguson N, Laydon D, Nedjati-Gilani G, Imai N, Ainslie K, Baguelin M, et al. Report 9: Impact of non-pharmaceutical interventions (NPIs) to reduce COVID19 mortality and healthcare demand. Imperial College London. 2020;10:77482.

[50] Lai S, Ruktanonchai NW, Zhou L, Prosper O, Luo W, Floyd JR, et al. Effect of non-pharmaceutical interventions for containing the COVID-19 outbreak in China. medRxiv. 2020;.

[51] Chen X, Qiu Z. Scenario analysis of non-pharmaceutical interventions on global COVID-19 transmissions. arXiv preprint arXiv:200404529. 2020;.

[52] Kasting ML, Head KJ, Hartsock JA, Sturm L, Zimet GD. Public perceptions of the effectiveness of recommended non-pharmaceutical intervention behaviors to mitigate the spread of SARS-CoV-2. PloS one. 2020;15(11):e0241662.

[53] Salanié F, Treich N. Public and private incentives for self-protection. The Geneva Risk and Insurance Review. 2020;45(2):104–113.

[54] Gray L, MacDonald C, Tassell-Matamua N, Stanley J, Kvalsvig A, Zhang J, et al. Wearing one for the team: views and attitudes to face covering in New Zealand/Aotearoa during COVID-19 Alert Level 4 lockdown. Journal of Primary Health Care. 2020;12(3):199–206.

[55] Pejó B, Biczók G. Corona Games: Masks, Social Distancing and Mechanism Design. In: Proceedings of the 1st ACM SIGSPATIAL International Workshop on Modeling and Understanding the Spread of COVID-19; 2020. p. 24–31.

[56] Pfattheicher S, Nockur L, Böhm R, Sassenrath C, Petersen MB. The emotional path to action: Empathy promotes physical distancing and wearing of face masks during the COVID-19 pandemic. Psychological Science. 2020;31(11):1363–1373.

[57] Tangherlini TR, Shahsavari S, Shahbazi B, Ebrahimzadeh E, Roychowdhury V. An automated pipeline for the discovery of conspiracy and conspiracy theory narrative frameworks: Bridgegate, Pizzagate and storytelling on the web. PloS one. 2020;15(6):e0233879.

[58] Weinstock D. A harm reduction approach to the ethical management of the COVID-19 pandemic. Public Health Ethics. 2020;13(2):166–175.

[59] Gray DM, Anyane-Yeboa A, Balzora S, Issaka RB, May FP. COVID-19 and the other pandemic: populations made vulnerable by systemic inequity. Nature Reviews Gastroenterology & Hepatology. 2020;17(9):520–522.

[60] Rosenstock IM, Strecher VJ, Becker MH. The health belief model and HIV risk behavior change. In: Preventing AIDS. Springer; 1994. p. 5–24.

[61] Fisher JD, Fisher WA. Theoretical approaches to individual-level change in HIV risk behavior. In: Handbook of HIV prevention. Springer; 2000. p. 3–55.

[62] Fisher JD, Fisher WA, Bryan AD, Misovich SJ. Information-motivation-behavioral skills model-based HIV risk behavior change intervention for inner-city high school youth. Health Psychology. 2002;21(2):177.

[63] Albarracín D, Gillette JC, Earl AN, Glasman LR, Durantini MR, Ho MH. A test of major assumptions about behavior change: a comprehensive look at the effects of passive and active HIV-prevention interventions since the beginning of the epidemic. Psychological bulletin. 2005;131(6):856.

[64] Raifman J, Nocka K, Jones D, Bor J, Lipson S, Jay J, et al. COVID-19 US state policy database. 2020;.

[65] Adolph C, Amano K, Bang-Jensen B, Fullman N, Wilkerson J. Pandemic politics: Timing state-level social distancing responses to COVID-19. medRxiv. 2020;.

[66] Keeley C, Jimenez J, Jackson H, Boudourakis L, Salway RJ, Cineas N, et al. Staffing Up For The Surge: Expanding The New York City Public Hospital Workforce During The COVID-19 Pandemic: Article describes how New York City’s public health care system rapidly expanded capacity across 11 acute-care hospitals and three new field hospitals to meet the challenges of the COVID-19 Pandemic. Health Affairs. 2020;39(8):1426–1430.

[67] Moghadas SM, Shoukat A, Fitzpatrick MC, Wells CR, Sah P, Pandey A, et al. Projecting hospital utilization during the COVID-19 outbreaks in the United States. Proceedings of the National Academy of Sciences. 2020;117(16):9122–9126.

[68] Cavallo JJ, Donoho DA, Forman HP. Hospital capacity and operations in the coronavirus disease 2019 (covid-19) pandemic—planning for the nth patient. In: JAMA Health Forum. vol. 1. American Medical Association; 2020. p. e200345–e200345.

[69] Weissman GE, Crane-Droesch A, Chivers C, Luong T, Hanish A, Levy MZ, et al. Locally informed simulation to predict hospital capacity needs during the COVID-19 pandemic. Annals of internal medicine. 2020;.

[70] Aktay A, Bavadekar S, Cossoul G, Davis J, Desfontaines D, Fabrikant A, et al. Google COVID-19 community mobility reports: Anonymization process description (version 1.0). arXiv preprint arXiv:200404145. 2020;.

[71] COVID Tracking Project at The Atlantic;.

[72] Ansari S, Hutchins C, Del Greco S. The NOAA Weather and Climate Toolkit. American Geophysical Union, Fall Meeting 2010, abstract id IN32A-06;.

[73] Max Roser EOO Hannah Ritchie, Hasell J. Coronavirus Pandemic (COVID-19). Our World in Data. 2020;.

[74] Kong E, Prinz D. Disentangling policy effects using proxy data: Which shutdown policies affected unemployment during the COVID-19 pandemic? Journal of Public Economics. 2020;189:104257.

[75] Gupta S, Nguyen TD, Rojas FL, Raman S, Lee B, Bento A, et al. Tracking public and private response to the covid-19 epidemic: Evidence from state and local government actions. National Bureau of Economic Research; 2020.

[76] Maneenop S, Kotcharin S. The impacts of COVID-19 on the global airline industry: An event study approach. Journal of air transport management. 2020;89:101920.

[77] Liu H, Manzoor A, Wang C, Zhang L, Manzoor Z. The COVID-19 outbreak and affected countries stock markets response. International Journal of Environmental Research and Public Health. 2020;17(8):2800.

[78] Alam MN, Alam MS, Chavali K. Stock market response during COVID-19 lockdown period in India: An event study. The Journal of Asian Finance, Economics, and Business. 2020;7(7):131–137.

[79] He P, Sun Y, Zhang Y, Li T. COVID–19’s impact on stock prices across different sectors—An event study based on the Chinese stock market. Emerging Markets Finance and Trade. 2020;56(10):2198–2212.

[80] GÖKER İEK, EREN BS, KARACA SS. The Impact of the COVID-19 (Coronavirus) on The Borsa Istanbul Sector Index Returns: An Event Study. Gaziantep Üniversitesi Sosyal Bilimler Dergisi. 2020;19(COVID-19 Special Issue):14–41.

[81] Lyu W, Wehby GL. Shelter-in-place orders reduced COVID-19 Mortality and reduced the rate of growth in hospitalizations: study examine effects of shelter-in-places orders on daily growth rates of COVID-19 deaths and hospitalizations using event study models. Health Affairs. 2020;39(9):1615–1623.

[82] Guan Wj, Ni Zy, Hu Y, Liang Wh, Ou Cq, He Jx, et al. Clinical characteristics of coronavirus disease 2019 in China. New England journal of medicine. 2020;382(18):1708–1720.

[83] To KKW, Tsang OTY, Leung WS, Tam AR, Wu TC, Lung DC, et al. Temporal profiles of viral load in posterior oropharyngeal saliva samples and serum antibody responses during infection by SARS-CoV-2: an observational cohort study. The Lancet Infectious Diseases. 2020;.

[84] Grasselli G, Zangrillo A, Zanella A, Antonelli M, Cabrini L, Castelli A, et al. Baseline characteristics and outcomes of 1591 patients infected with SARS-CoV-2 admitted to ICUs of the Lombardy Region, Italy. JAMA. 2020;323(16):1574–1581.

[85] Guo YR, Cao QD, Hong ZS, Tan YY, Chen SD, Jin HJ, et al. The origin, transmission and clinical therapies on coronavirus disease 2019 (COVID-19) outbreak–an update on the status. Military Medical Research. 2020;7(1):1–10.

[86] Bi Q, Wu Y, Mei S, Ye C, Zou X, Zhang Z, et al. Epidemiology and transmission of COVID-19 in 391 cases and 1286 of their close contacts in Shenzhen, China: a retrospective cohort study. The Lancet Infectious Diseases. 2020;.

[87] COVID-19 Pandemic Planning Scenarios;.

[88] Dietz K. The estimation of the basic reproduction number for infectious diseases. Statistical methods in medical research. 1993;2(1):23–41.

[89] Havers FP, Reed C, Lim T, Montgomery JM, Klena JD, Hall AJ, et al. Seroprevalence of antibodies to SARS-CoV-2 in 10 sites in the United States, March 23-May 12, 2020. JAMA internal medicine. 2020;.

[90] Angrist JD, Pischke JS. Mostly harmless econometrics: An empiricist’s companion. Princeton university press; 2008.

[91] Makridis C, Rothwell J. The Real Cost of Political Polarization: Evidence from the COVID-19 Pandemic. 2020;.

[92] Gaure S. OLS with multiple high dimensional category variables. Computational Statistics & Data Analysis. 2013;66:8–18.

